# Harnessing consumer wearable digital biomarkers for individualized recognition of postpartum depression using the *All of Us* Research Program dataset

**DOI:** 10.1101/2023.10.13.23296965

**Authors:** Eric Hurwitz, Zachary Butzin-Dozier, Hiral Master, Shawn T. O’Neil, Anita Walden, Michelle Holko, Rena C. Patel, Melissa A. Haendel

## Abstract

Postpartum depression (PPD), afflicting one in seven women, poses a major challenge in maternal health. Existing approaches to detect PPD heavily depend on in-person postpartum visits, leading to cases of the condition being overlooked and untreated. We explored the potential of consumer wearable-derived digital biomarkers for PPD recognition to address this gap. Our study demonstrated that intra-individual machine learning (ML) models developed using these digital biomarkers can discern between pre-pregnancy, pregnancy, postpartum without depression, and postpartum with depression time periods (i.e., PPD diagnosis). When evaluating variable importance, calories burned from the basal metabolic rate (calories BMR) emerged as the digital biomarker most predictive of PPD. To confirm the specificity of our method, we demonstrated that models developed in women without PPD could not accurately classify the PPD-equivalent phase. Prior depression history did not alter model efficacy for PPD recognition. Furthermore, the individualized models demonstrated superior performance compared to a conventional cohort-based model for the detection of PPD, underscoring the effectiveness of our individualized ML approach. This work establishes consumer wearables as a promising avenue for PPD identification. More importantly, it also emphasizes the utility of individualized ML model methodology, potentially transforming early disease detection strategies.

## Introduction

Postpartum depression (PPD) is the most common complication of childbirth, occurring in approximately one in seven women^1^. PPD can have several implications for women, manifesting in ways such as irritability, mood swings, fatigue, sleep and appetite disturbance, and thoughts of suicide^2^. Undetected PPD has also been shown to have financial implications for affected individuals, as it can lead to challenges in maintaining employment or reduced work performance^3^. Furthermore, PPD has been linked to an elevated risk of mood disorders in the child as well as paternal depression^4,5^.

Unfortunately, PPD remains significantly underdiagnosed and undertreated, as indicated by the strikingly low treatment rate of only 15%^6^. The current method of diagnosing PPD relies on screening instruments, such as the Edinburgh Postnatal Depression Score (EPDS), Center for Epidemiologic Studies of Depression instrument (CES-D), Patient Health Questionnaire (PHQ-9), and Postpartum Depression Screening Scale (PDSS), where the EPDS is the most commonly used instrument^7^. Often, women also need to undergo blood tests to assess thyroid function, as the symptoms of PPD frequently overlap with hyperthyroidism^7^. Due to the challenges in diagnosing PPD, traditional approaches using these screening tools contribute to inadequate screening of women and subsequent underdiagnosis^8,9^. Therefore, the advent of new technologies is greatly needed to enable adequate, and hopefully earlier, detection of PPD.

Digital health tools have been gaining traction in recent years due to the near-ubiquitous ownership of smartphones^10^. Leveraging data passively collected by wearables (i.e., digital biomarkers such as the average heart rate, total steps, and calories burned per day), coupled with machine learning (ML) algorithms, provides an opportunity to model the relationship between digital biomarkers and a particular disease for early recognition. Previous studies have demonstrated that ML algorithms using digital biomarkers from smartwatches can predict cardiovascular diseases, infection, diabetes, and mental health conditions^11–14^. For example, one study demonstrated that a wearable device could estimate the changes in the severity of patients with major depressive disorder (MDD), where their findings indicated that ML models exclusively utilizing digital biomarkers from wearables achieved moderate performance with correlation coefficients of 0.56 (95% CI: 0.39-0.73) and 0.54 (95% CI: 0.49, 0.59) in the time-split and user-split scenarios between model predictions and actual Hamilton Depression Rating Scale (HRDS) scores, respectively^15^. Another study recruited moderately depressed individuals for four weeks to develop individualized ML models based on digital biomarkers to predict mood. Their findings displayed a correlation between digital biomarkers and depression, as evidenced by high-performing models with a mean absolute error (MAE) of 0.77 ± 0.27 points on the 7-point Likert scale, which corresponds to a mean absolute percent error (MAPE) of 27.9 ± 10.3%^16^. Notably, both studies adopted an individualized (intra-individual) framework for developing ML models, corroborating its potential for depression detection. However, while these studies highlight a relationship between digital biomarkers and depression, they suffer from the following limitations: 1) they do not assess whether models are applicable in a cohort of postpartum women to detect PPD; and 2) they utilize data in the model that need active patient engagement with partnered mobile applications, where user retention is known to decrease over time with health-related apps^17^. Therefore, a method that provides continuous monitoring without the need for clinical encounters to enable early detection of mental health disorders, including PPD, is needed.

The *All of Us* Research Program (AoURP) is a comprehensive dataset that collects several health-related data, including surveys, electronic health records (EHRs), physical measurements, and wearable data from Fitbit devices, with an emphasis on patient populations that have been previously underrepresented in biomedical research^18^. Currently, the longitudinal Fitbit data from approximately 13,000 AoURP participants are made available to registered researchers on the *All of Us* Researcher Workbench, providing an opportunity to explore digital biomarkers in a diverse cohort of participants.

It is unknown whether digital biomarkers from consumer wearables can be used to detect PPD. Here, we combine several orthogonal approaches demonstrating that digital biomarkers can be employed for individualized classification of PPD with data passively collected from Fitbit using the AoURP (Figure 1). As such, our findings uncover a novel method for recognizing PPD and serve as a framework that can be leveraged to facilitate early PPD detection.

**Figure 1:**
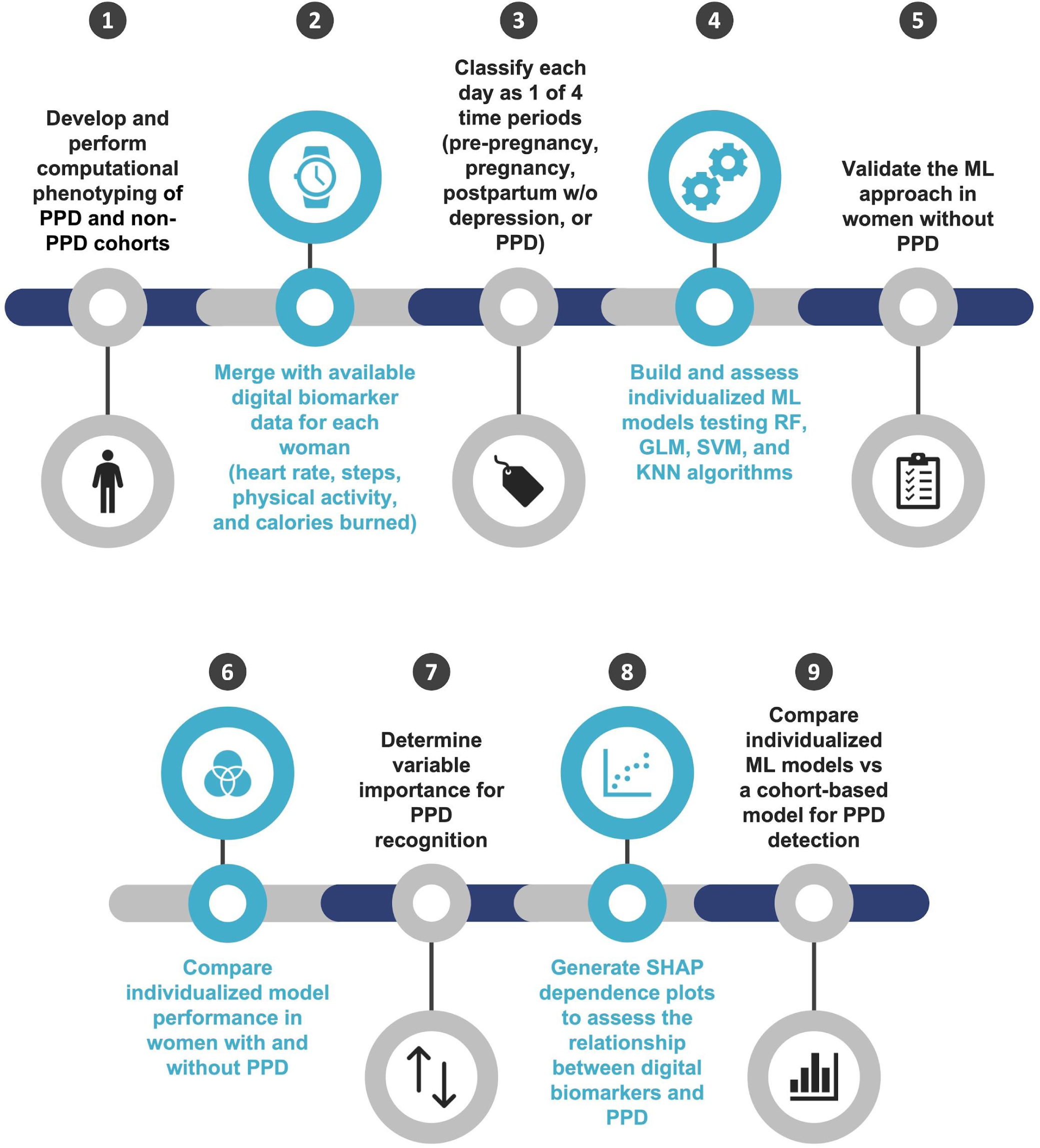
An overview of the analysis workflow to evaluate the potential for digital biomarkers in postpartum depression (PPD) recognition. *RF = random forest, GLM = generalized linear models, SVM = support vector machine, KNN = k-nearest neighbors

## Results

### Descriptive statistics

Through computational phenotyping in the AoURP, a patient cohort of women who gave birth with PPD (n = <20) and without PPD (n = 39) provided valid Fitbit data (Figure 2). The median age was 35.60 (interquartile range (IQR) 4.53) years, compared to those in the non-PPD cohort, which was 33.60 (IQR 4.85). The median and IQR were calculated for each digital biomarker across all women in the PPD and non-PPD cohorts (Table 1). In both the PPD and non-PPD cohorts, we computed the median and IQR number of days with digital biomarker data during the pre-pregnancy, pregnancy, postpartum, and PPD (or PPD equivalent) time periods (additional details about the PPD-equivalent time period, a similar fourth time period for those without PPD, can be found in the Materials and Methods section) (Table 1). Briefly, digital biomarkers included in this analysis were daily average heart rate (HR), standard deviation HR, minimum HR, Q1 HR, median HR, Q3 HR, maximum HR, sum of steps, activity calories, calories BMR, calories out, fairly active minutes, lightly active minutes, marginal calories, sedentary minutes, and very active minutes (see descriptions in Supplementary Table 1).

**Figure 2:**
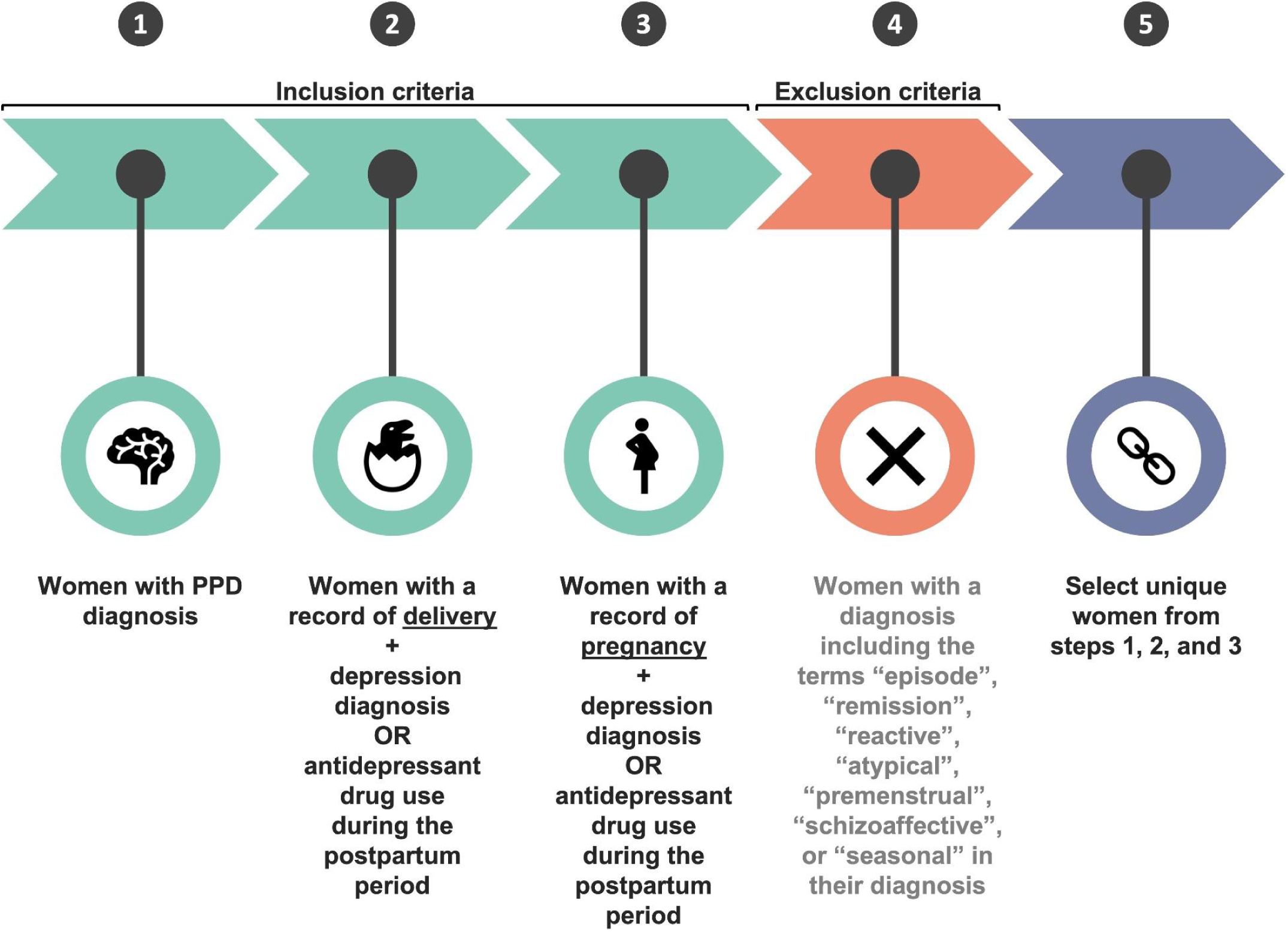
A schematic of PPD computational phenotyping.

**Table 1:** Descriptive statistics of the postpartum depression (PPD) and non-PPD patient cohorts in the AoURP.

| Sample size |  |
| --- | --- |
| PPD cohort | Non-PPD |
| <20 | 39 |

| Age (years) |  |  |  |
| --- | --- | --- | --- |
| PPD cohort |  | Non-PPD |  |
| Median | IQR | Median | IQR |
| 35.60 | 4.53 | 33.60 | 4.85 |

| Digital biomarkers |  |  |  |  |  |
| --- | --- | --- | --- | --- | --- |
|  | PPD |  | Non-PPD |  |  |
| Digital biomarker | Median | IQR | Median | IQR | Units |
| Average HR | 74.23 | 12.30 | 78.31 | 11.65 | bpm |
| SD HR | 12.18 | 3.47 | 12.70 | 4.39 | bpm |
| Minimum HR | 54.00 | 11.00 | 57.00 | 9.00 | bpm |
| Q1 HR | 64.00 | 12.00 | 68.00 | 12.00 | bpm |
| Median HR | 72.00 | 12.00 | 76.00 | 12.00 | bpm |
| Q3 HR | 81.00 | 14.00 | 85.00 | 14.00 | bpm |
| Maximum HR | 124.00 | 18.00 | 127.00 | 22.00 | bpm |
| Sum steps | 7567.50 | 5652.25 | 7352.00 | 5996.00 | steps |
| Activity calories | 989.00 | 520.25 | 964.00 | 591.00 | calories |
| Calories BMR | 1466.00 | 160.00 | 1390.00 | 156.00 | calories |
| Calories out | 2236.00 | 471.25 | 2180.00 | 540.50 | calories |
| Fairly active minutes | 9.00 | 24.00 | 8.00 | 23.00 | minutes |
| Lightly active minutes | 245.00 | 126.00 | 245.00 | 122.50 | minutes |
| Marginal calories | 501.00 | 316.00 | 489.00 | 358.00 | calories |
| Sedentary minutes | 646.00 | 178.00 | 710.00 | 273.50 | minutes |
| Very active minutes | 2.00 | 18.00 | 4.00 | 21.00 | minutes |

| Number of days in each time period across all women |  |  |  |  |
| --- | --- | --- | --- | --- |
|  | PPD cohort |  | Non-PPD cohort |  |
| Time period | Median | IQR | Median | IQR |
| Pre-pregnancy | 206.00 | 159.00 | 227.00 | 231.25 |
| Pregnancy | 258.00 | 38.00 | 221.00 | 140.50 |
| Postpartum | 42.00 | 62.50 | 72.00 | 36.00 |
| PPD | 42.50 | 4.50 | 29.00 | 16.50 |

\*HR = heart rate, SD = standard deviation, BMR = basal metabolic rate

### Digital biomarker comparison across time periods of pregnancy reveals altered profiles and heterogeneity between women

Because of the known heterogeneity in depressive symptoms, we hypothesized that variability in digital biomarkers may exist across individuals in the PPD cohort^19^. To probe this hypothesis, we conducted linear mixed-effects models for each digital biomarker in women with PPD, where we found the random effect of person ID was significant for all digital biomarkers, suggesting meaningful variability across individuals (Supplementary Table 2). These results coupled with a smaller cohort sample size prompted us to perform subsequent analyses using an intra-individual approach.

In women with PPD, we next sought to compare whether there was a difference in digital biomarkers across different time periods of pregnancy, including pre-pregnancy, pregnancy, postpartum, and PPD (where PPD represents both a time period and diagnosis). Therefore, an intra-individual interrupted time series analysis (ITSA) and Tukey Honest Significance Differences (HSD) tests were conducted for each digital biomarker. Because of the physiological changes associated with pregnancy, such as increases in blood and stroke volume, in addition to the behavioral fluctuations that occur during PPD, like a loss of energy and psychomotor retardation, we hypothesized that all digital biomarkers (those related to heart rate, steps, physical activity, and calories burned), would be altered across pre-pregnancy, pregnancy, postpartum, and PPD time periods^20–23^. ITSA results supported our hypothesis and demonstrated a significant difference in all digital biomarkers across time periods in the majority of women with PPD (Supplementary Table 3). Consistent with ITSA findings, Tukey HSD results showed that several digital biomarkers were significantly altered between PPD and other time periods (pre-pregnancy, pregnancy, and the postpartum period) (Supplementary Tables 4 and 5). We further observed various trends in digital biomarkers between pairs of time periods (i.e., PPD and pre-pregnancy, PPD and pregnancy, PPD and postpartum, etc.) (Supplementary Table 6).

### Individualized ML models effectively differentiate PPD from alternative time periods of pregnancy

Having seen that digital biomarkers were significantly altered across multiple time periods of pregnancy within women with PPD, we surmised that individualized multinomial ML models could accurately distinguish between our four time periods of pregnancy (pre-pregnancy, pregnancy, postpartum, or PPD) (Supplementary Tables 3-6). To probe this hypothesis, intra-individual ML models were generated using random forest (RF), generalized linear model (GLM), support vector machine (SVM), and K-nearest neighbor (KNN) to conclude which algorithm would yield the best-performing results. Models were assessed using a combination of the multiclass area under the Receiving Operator Characteristics curve (hereafter referred to as mAUC) and kappa, which are two frequently used metrics^24,25^. After averaging the mAUC for individual models within each algorithm, the results revealed that RF models performed the best, followed by GLM, SVM, then KNN with an average mAUC of 0.85, 0.82, 0.75, and 0.74, respectively (Figure 3A). Assessing models in a similar fashion using another metric, kappa, displayed concordant results in RF (0.80), GLM (0.74), SVM (0.72), and KNN (0.62) model performance, suggesting that the RF algorithm had the best performance and should be used going forward (Figure 3B).

**Figure 3:**
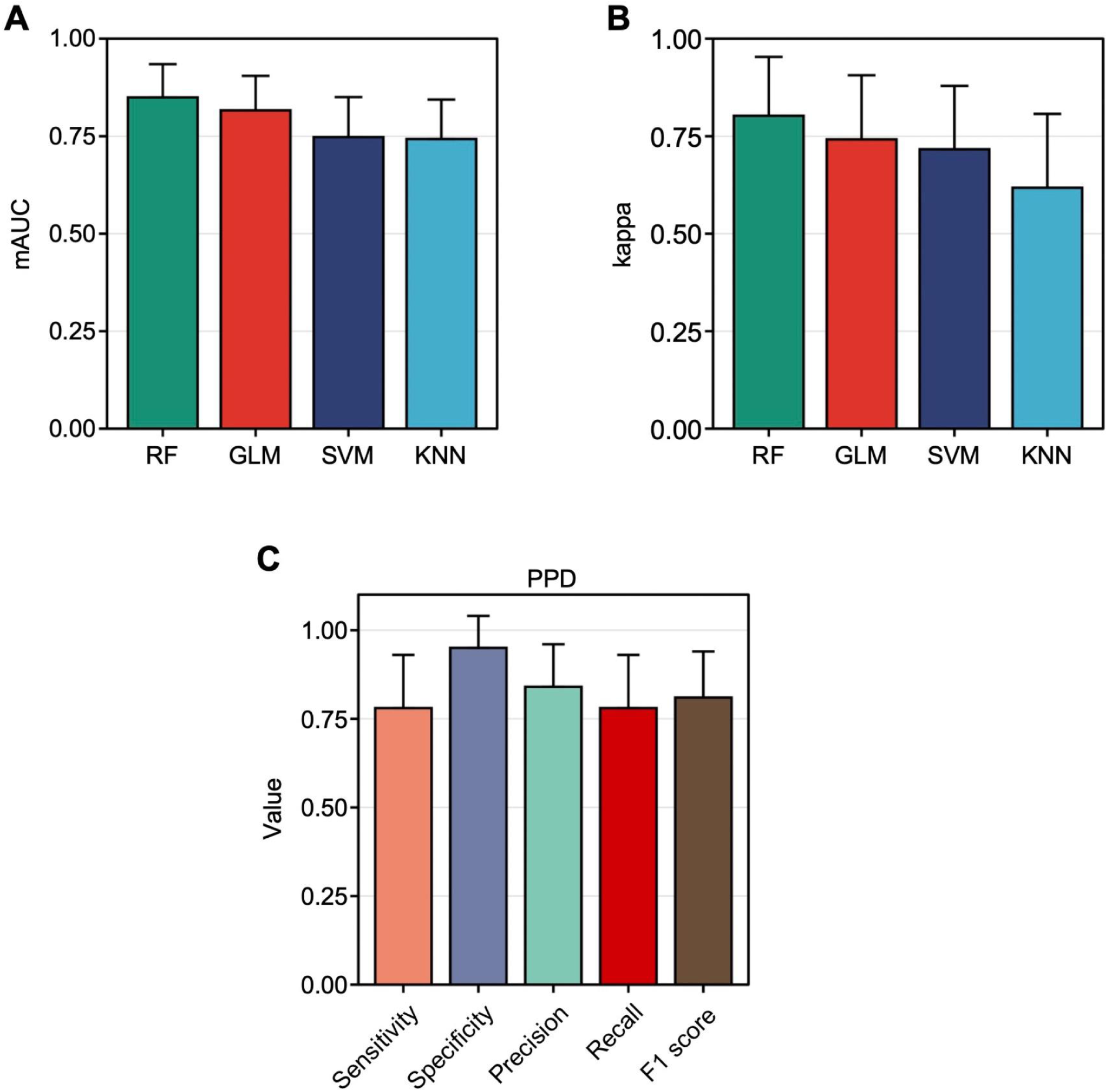
Individualized RF models exhibited the best performance for multinomial time period classification. **A:** The multiclass AUC (mAUC) across individual random forest (RF), generalized linear model (GLM), support vector machine (SVM), and K-nearest neighbor (KNN) models. **B:** The kappa value across individual RF, GLM, SVM, and KNN models. **C:** The sensitivity, specificity, precision, recall, and F1 score across individualized multinomial RF models for the PPD time period. The average mAUC and kappa values of individualized RF models were higher than that of GLM, SVM, and KNN models. Utilizing individualized RF models resulted in an average sensitivity, specificity, precision, recall, and F1 score with good performance for recognizing the PPD time period. Data are expressed as mean ± standard deviation (SD) in A-C.

Since our analysis aimed to assess the potential of digital biomarkers for personalized classification of PPD, we sought to further examine each RF model’s performance via a confusion matrix. Thus, the average sensitivity, specificity, precision, recall, and F1 score were calculated across all individual models, where the results for the PPD class were 0.79, 0.95, 0.84, 0.79, and 0.81, respectively (Figure 3C). The same metrics for the pre-pregnancy, pregnancy, and postpartum classes were also calculated (Supplementary Figures 1A-C).

In order to ensure the widespread applicability of these algorithms to a diverse range of women, we did not exclude individuals with a history of depression either before or during pregnancy. Therefore, we sought to determine whether having depression pre-pregnancy or during pregnancy impacted individual model performance, specifically for recognizing the PPD class. To answer this question, we computed the average sensitivity, specificity, precision, recall, and F1 score within the group of women experiencing PPD, categorized based on their depression history: 1) no prior history of depression; 2) history pre-pregnancy; 3) history during pregnancy; or 4) history of both pre-pregnancy and during pregnancy. Notably, the findings revealed no statistically significant variations in any of these metrics between women with a history of depression during the pre-pregnancy and/or pregnancy time periods and those without such a history (Supplementary Figure 2). Promisingly, this suggests the potential for a forthcoming technology focused on detecting PPD through digital biomarkers to be relevant for women with or without a previous history of depression before and/or during pregnancy.

### Individualized ML models for PPD recognition were specific

To validate our approach of using digital biomarkers in individualized ML models for PPD detection, we aimed to test our strategy in a cohort of women who had given birth but did not experience PPD. Given that women without PPD didn’t have a distinct PPD-specific time period as observed in the PPD cohort, we introduced a fourth time segment in the non-PPD cohort (hereafter referred to as the PPD-equivalent time period). Following the same ML pipeline as the PPD cohort, individualized RF models were built for women in the non-PPD cohort. If our conjecture holds, we anticipate observing elevated model metrics during the pre-pregnancy and pregnancy time periods, accompanied by diminished performance in the postpartum and PPD-equivalent time segments. This expectation arises from the idea that digital biomarkers remain unaltered during the postpartum and PPD-equivalent time periods, resulting in the model’s inability to differentiate between them.

In line with our hypothesis, the sensitivity, specificity, precision, recall, and F1 scores substantiated that ML models effectively identified the pre-pregnancy (0.89, 0.91, 0.88, 0.89, and 0.88, respectively) and pregnancy (0.85, 0.91, 0.87, 0.85, and 0.86, respectively) time intervals through digital biomarkers (Figure 4). When compared to model performance in the pre-pregnancy and pregnancy time periods, there was no significant reduction in model performance during the postpartum period (0.74, 0.96, 0.76, 0.74, and 0.75, respectively); however, a noticeable decline in performance was observed during the PPD and PPD-equivalent time periods(0.52, 0.99, 0.69, 0.52, and 0.61, respectively) (Figure 4). To further assess potential variations in the classification performance between the PPD and PPD-equivalent time periods, we carried out a t-test comparing the average sensitivity, specificity, precision, recall, and F1 score between the PPD and non-PPD cohorts for these respective time periods. The findings indicated a statistically significant decrease in sensitivity, precision, recall, and F1 score when predicting the PPD-equivalent time period in the non-PPD cohort, as opposed to predicting the PPD time period within the PPD cohort (Figure 5). On the other hand, specificity remained largely unchanged (Figure 5). Collectively, these outcomes offered a layer of validation to the effectiveness of this approach in identifying PPD, reinforcing the agreement that personalized models utilizing digital biomarkers can indeed effectively recognize PPD.

**Figure 4:**
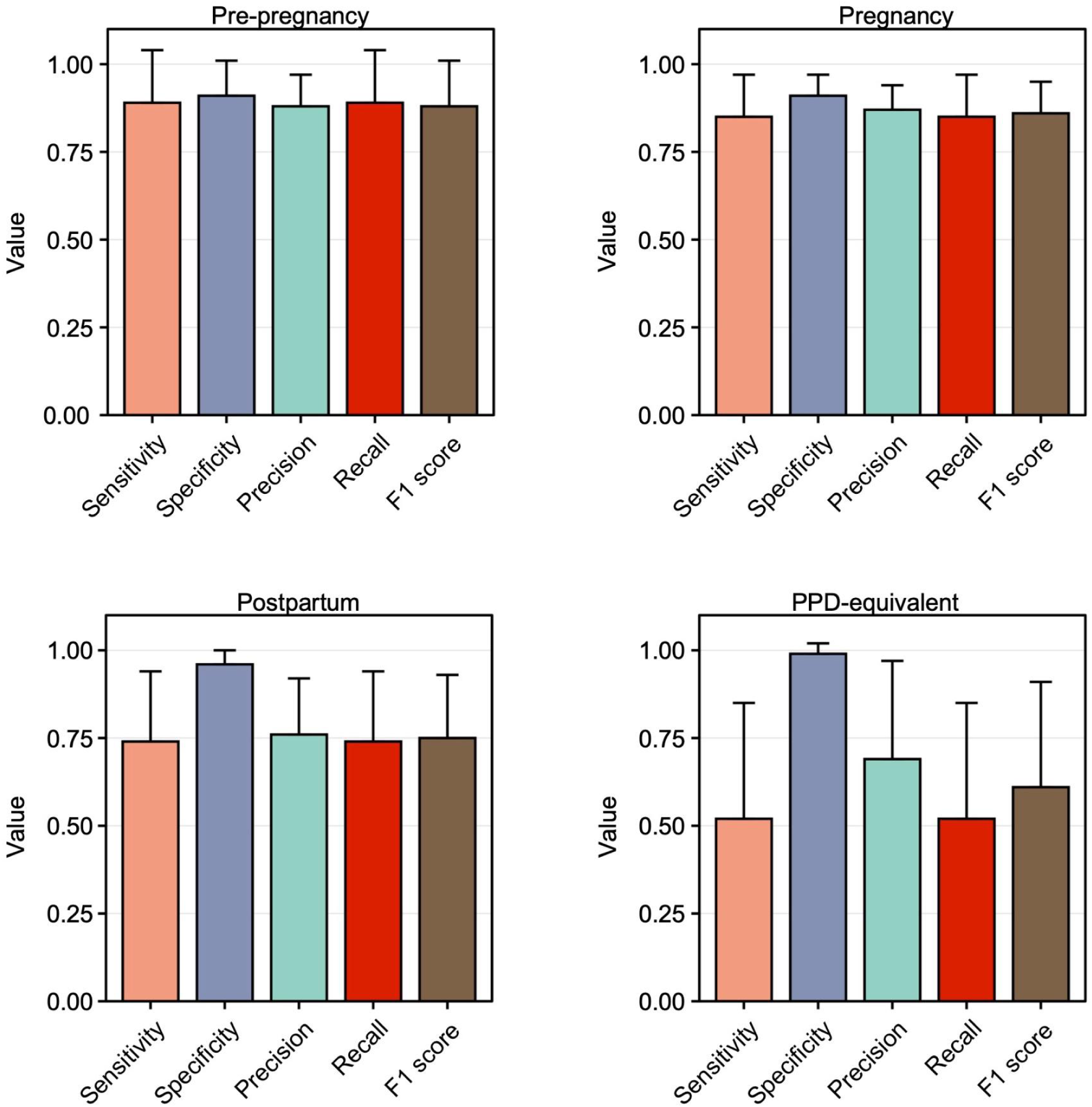
ML models did not accurately detect the PPD-equivalent time period in women without PPD. The sensitivity, specificity, precision, recall, and F1 score of individualized ML models in women without PPD for predicting the pre-pregnancy (top left), pregnancy (top right), postpartum (bottom left), and PPD-equivalent (bottom right) time periods. The sensitivity, specificity, precision, recall, and F1 score were diminished for recognizing the PPD time period compared to the pre-pregnancy, pregnancy, and postpartum time periods. Data are expressed as mean ± SD.

**Figure 5:**
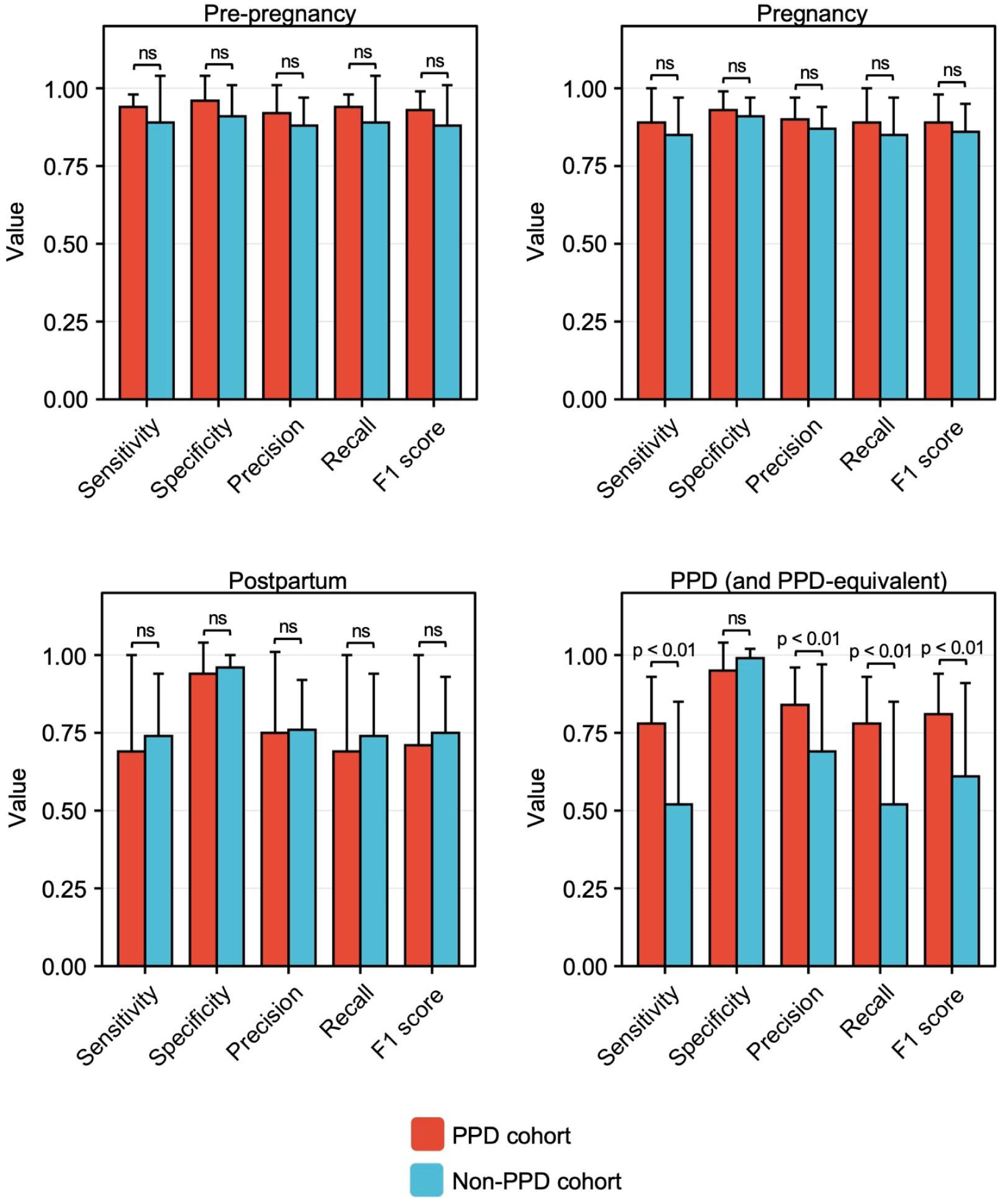
Individualized ML models for PPD recognition outperformed those in women without PPD detecting the PPD-equivalent time period. The sensitivity, specificity, precision, recall, and F1 score across individual RF models for women in the non-PPD cohort for the pre-pregnancy (top left), pregnancy (top right), postpartum (bottom left), and PPD or PPD-equivalent time periods (bottom right). On average, individualized model performance was not significantly different for sensitivity, specificity, precision, recall, and F1 score for predicting the pre-pregnancy, pregnancy, or postpartum time periods between women in the PPD and non-PPD cohorts. Individualized model performance was reduced for sensitivity, precision, recall, and F1 score, while specificity did not differ between the PPD and non-PPD cohorts. Data are expressed as mean ± SD.

### Calories burned during basal metabolism (calories BMR) was the most predictive digital biomarker of PPD

In order to elucidate which digital biomarkers were most predictive of the PPD class, we performed SHapley Additive exPlanations (SHAP) to explain individual predictions for each digital biomarker across all random forest intra-individual models^26^. Features were sorted based on their predictive value for the PPD class within each individual model, and subsequently, the occurrences of each digital biomarker ranking in the top five across all intra-individual models were tallied, where an example beeswarm plot for one woman is shown in Supplementary Figure 3. This process aimed to identify whether any digital biomarkers consistently played a crucial role in predicting the PPD class. The results displayed that the top five features most frequently ranked in the top five were calories BMR, average HR, Q1 HR, lightly active minutes, and minimum HR (Figure 6A). Interestingly, calories BMR ranked in the top five features predictive of the PPD class in 95-100% of the models and was the number one rated digital biomarker in 80-85% of individualized models (Figure 6A).

**Figure 6:**
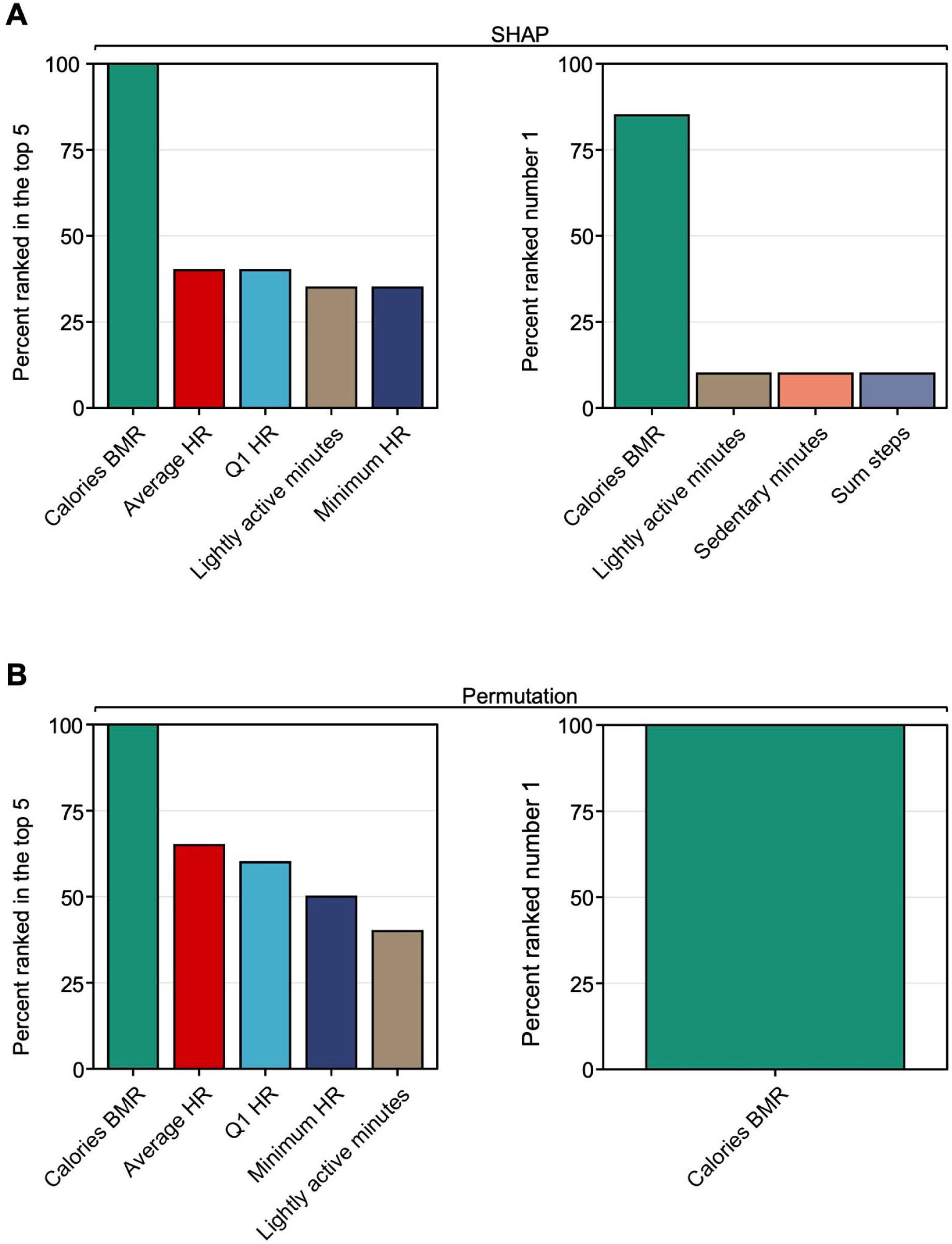
The variable importance rankings demonstrated that calories burned during the basal metabolic rate (calories BMR) is the most predictive digital biomarker of the PPD class. **A:** The percentage of women with the top five digital biomarkers ranked among the top five most predictive features of the PPD class (top left) and the most predictive feature of the PPD class (top right) based on SHAP. **B:** The percent of women with the overall top five digital biomarkers ranked among the top five most predictive features of the PPD class (bottom left) and the most predictive feature of the PPD class (bottom right) based on a permutation-based approach of variable importance. Calories BMR, average HR, Q1 HR, lightly active minutes, and minimum HR most frequently emerged as the top five digital biomarkers with the highest predictive value for the PPD time period within the PPD cohort. Calories BMR most often ranked as the number one digital biomarker predictive of PPD.

To add a layer of robustness to our approach assessing which features were most predictive of the PPD class, the variable importance of each digital biomarker was also calculated using a permutation approach^27^. Consistent with our findings obtained using SHAP, the top five ranking digital biomarkers for the PPD class were calories BMR, average HR, Q1 HR, minimum HR, and lightly active minutes (Figure 6B). Calories BMR again ranked in the top five digital biomarkers predictive of the PPD class 95-100% of the time and ranking number one 95-100% of the time (Figure 6B).

Because of the intriguing observation that calories BMR was highly predictive of PPD across all models, we sought to better understand its relationship with the PPD class in our models using SHAP dependence plots to visualize and calculate the Pearson correlation coefficient between the PPD time period with the pre-pregnancy, pregnancy, or postpartum time periods (see Supplementary Figures 4A-C for example plots from individual women). Across all individual SHAP dependence plots of calories BMR filtered in the pre-pregnancy/PPD time periods, our initial observation revealed that 95-100% of women exhibited a significant Pearson correlation coefficient (Figure 7A). Among these, 60-65% displayed a positive relationship, indicating an elevated calories BMR during the PPD period relative to pre-pregnancy (Figure 7A). Compared to pre-pregnancy/PPD, it was observed that 75-80% and 85-90% of individualized SHAP dependence plots of calories BMR during pregnancy/PPD and postpartum/PPD time periods exhibited a significant Pearson correlation coefficient, respectively (Figure 7A). Of those, 60-65% and 85-90% of women during pregnancy/PPD and postpartum/PPD demonstrated a negative relationship, respectively, suggesting a decrease in calories BMR is predictive of PPD relative to pregnancy and postpartum time periods (Figure 7A). SHAP dependence plots were also generated for individualized models of the other top four digital biomarkers predictive of PPD (average HR, Q1 HR, minimum HR, and lightly active minutes) in pre-pregnancy/PPD, pregnancy/PPD, and postpartum/PPD time periods (Figure 7A). Notably, during the pre-pregnancy/PPD time periods, half of the women exhibited a positive relationship in plots of lightly active minutes, indicating an increase in lightly active minutes associated with PPD in those models (Figure 7A). To examine the rise in lightly active minutes relative to other digital biomarkers of motion (sedentary minutes, fairly active minutes, and very active minutes), we calculated the ratio of the number of lightly active minutes to each of the three other digital biomarkers across all individuals. Here, we observed that the average (and standard deviation) ratios of lightly active minutes to sedentary minutes, fairly active minutes, and very active minutes were 0.35 (0.49), 17.7 (4.92), and 21.72 (5.11), respectively (Supplementary Figure 5).

**Figure 7:**
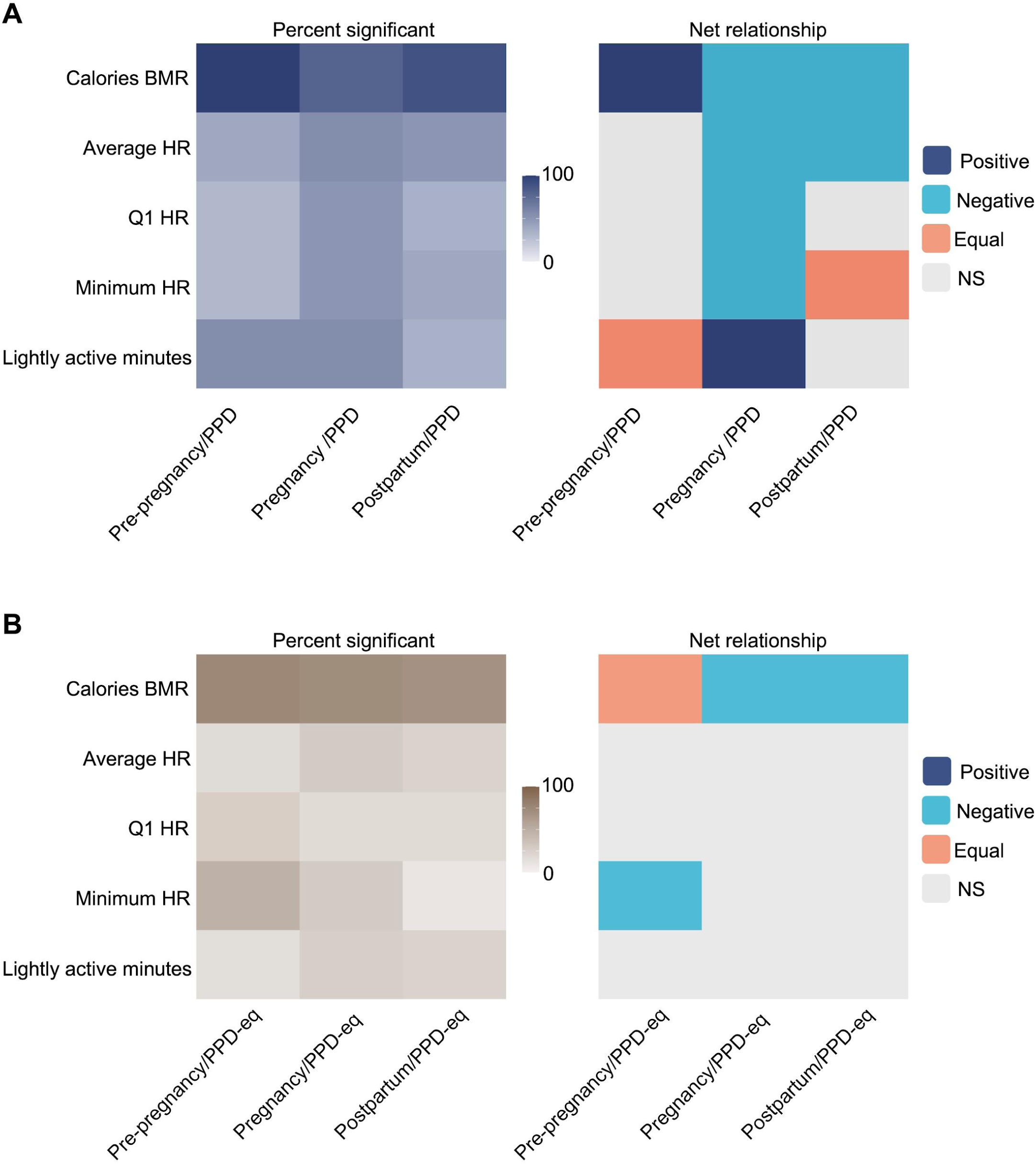
The direction of digital biomarkers in ML models for PPD classification was heterogeneous. **A:** The percent of women in the PPD cohort with a significant Pearson correlation (left) and the net relationship (right) for the top five overall ranked digital biomarkers for PPD classification. **B:** The percent of women in the non-PPD cohort with a significant Pearson correlation (left) and the net relationship (right) for the top five overall ranked digital biomarkers for PPD-equivalent classification. The proportion of women displaying a significant Pearson correlation coefficient between SHAP values and digital biomarkers varied in both the PPD and non-PPD cohorts. Among those demonstrating a significant relationship in SHAP dependence plots during the pre-pregnancy/PPD (and pre-pregnancy/PPD-equivalent) time periods, the correlation pattern for SHAP values and calories BMR differed: the majority of women exhibited a positive correlation in the PPD cohort, while there was no uniform pattern amongst women in the PPD-negative cohort. For women in the pregnancy/PPD and postpartum/PPD (and PPD-equivalent) time periods, the majority of women demonstrated a negative relationship between SHAP values and calories BMR in both the PPD-positive and negative cohorts.

For a more comprehensive evaluation of the connection between calories BMR and PPD, we also crafted SHAP dependence plots from individualized ML models for women without PPD. When first assessing the number of women with a significant correlation in SHAP dependence plots of pre-pregnancy/PPD-equivalent, pregnancy/PPD-equivalent, and postpartum/PPD-equivalent time periods, the results displayed that 75-80%, 70-75%, and 65-70% had a significant relationship, respectively (Figure 7B). Of those, there were an equal number of women with a positive and negative relationship in the pre-pregnancy/PPD-equivalent time periods, compared to the PPD cohort where the majority of women (60-65%) exhibited a positive relationship (Figure 7B). This implies that among women in the PPD cohort, an escalation in calories BMR corresponds to a higher likelihood of PPD when compared to the pre-pregnancy time period (Figures 7A and 7B). On the other hand, within the non-PPD cohort, there is no uniform pattern of association between calories BMR during the pre-pregnancy and the PPD-equivalent time periods across all women, highlighting the distinctive nature of our observation. During the pregnancy/PPD-equivalent and postpartum/PPD-equivalent time frames, 80-85% and 75-80% of women, respectively, exhibited a significant correlation in SHAP dependence plots between calories BMR and Shapley values (Figure 7B). As anticipated, this follows a similar pattern to women in the PPD cohort (Figure 7A). These findings implied that a reduction in caloric intake and BMR is linked to PPD (or PPD-equivalent) time periods in contrast to the pregnancy or postpartum periods (Figures 7A and 7B).

To showcase the effectiveness of our approach employing individualized ML models for PPD detection, we constructed an ML model using conventional techniques. In this endeavor, we harnessed the PPD and PPD-equivalent time periods of the PPD and non-PPD cohorts, respectively, enabling a comparative assessment of our individualized approach compared to conventional methods using a binomial model for the classification of individuals with or without PPD. By evaluating model outcomes through metrics such as sensitivity, specificity, precision, recall, and F1 score, we found that the average performance of the individualized model surpassed that of the cohort-based strategy (Figure 8). Specifically, in the individualized approach, we observed sensitivity, specificity, precision, recall, and F1 score values of 0.78, 0.95, 0.84, 0.78, and 0.81, respectively, in contrast to 0.54, 0.55, 0.49, 0.54, and 0.52 for the cohort-based approach (Figure 8).

**Figure 8:**
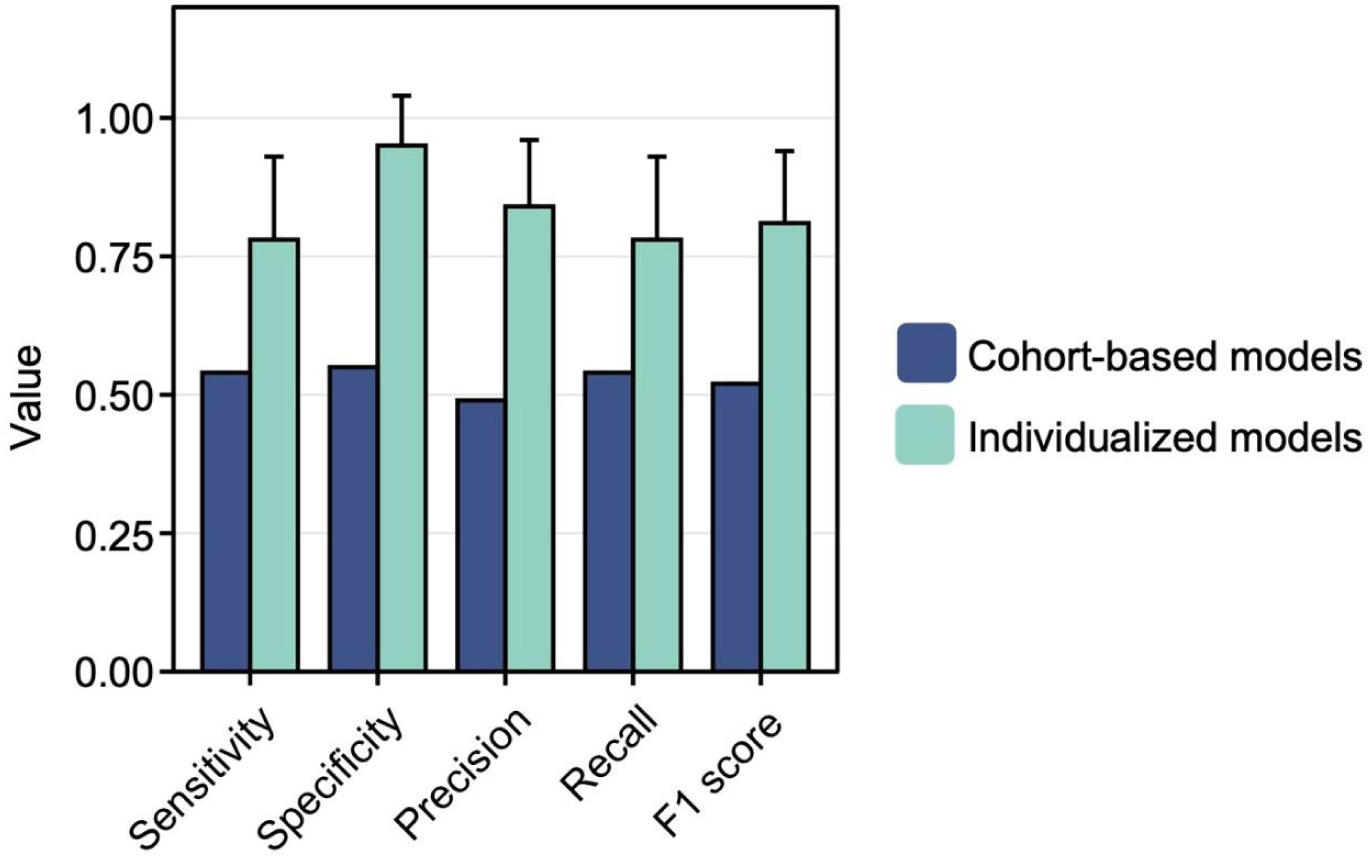
Individualized ML models outperformed a cohort-based model for PPD recognition. The sensitivity, specificity, precision, recall, and F1 score of individualized ML models in women in the PPD cohort detecting the PPD time period were compared to a cohort-based model for PPD classification. Data are expressed as mean ± SD. On average, the sensitivity, specificity, precision, recall, and F1 score were elevated in individualized ML models using digital biomarkers among women in the PPD cohort, specifically for the PPD time period, compared to a conventional binomial model designed for PPD or non-PPD classification using digital biomarkers.

## Discussion

Here, our multifaceted analysis demonstrated that 1) digital biomarkers differed between phases of pre-, during, and post-pregnancy periods (up to 2 years pre-pregnancy, pregnancy, postpartum, and PPD) (Supplementary Tables 3-6); 2) personalized N-of-1 ML models using digital biomarkers from consumer-grade wearables were able to classify PPD and other time periods of pregnancy (Figure 3 and Supplementary Figure 1); 3) a history of depression before or during pregnancy did not impact individualized ML model performance for PPD recognition (Supplementary Figure 2); 4) calories BMR, average HR, Q1 HR, lightly active minutes, and minimum HR were the most influential digital biomarkers in predicting the PPD time period across all individualized models (Figures 6A and 6B); and 5) individualized ML models for PPD recognition outperformed the traditional cohort-based model approach (Figure 8). The results presented in this work provide a new opportunity for the potential to leverage passively collected digital biomarkers from consumer-grade wearables to facilitate early detection of PPD.

To the best of our knowledge, this is the first study presenting that individualized ML models using passively collected digital biomarkers from consumer-grade wearables can recognize PPD. PPD is most commonly diagnosed using the EPDS, which suffers from the following limitations: 1) postpartum women must attend follow-up visits assessed by care providers for PPD screening, where the rate of postpartum visits is highly variable; 2) using the EPDS only captures the mental health of a woman at a single point in time; and 3) the EPDS uses self-reported symptoms, which may not be representative of a patient’s actual mental health status^28–30^. For these reasons, the development of our approach using passively monitored digital biomarkers from consumer wearable technology may serve as an effective tool for facilitating the detection of PPD in an individualized fashion, especially in non-clinical settings.

Because of the variation in digital biomarkers detected between women across different time periods, our limited sample size, and the availability of continuous intra-individual data, our study was geared towards an individualized analytic approach (Supplementary Table 2). The observed variability across individuals is consistent to previous studies that have emphasized the heterogeneous nature of depression prompting individualized methodologies^16,19,31–33^. Moreover, ITSA and Tukey HSD results revealed that digital biomarkers were significantly altered between time periods within each woman (Supplementary Tables 3-6). Overall, there were numerous individual-level alterations, which can be explained by the considerable heterogeneity in depressive symptoms^19^. Collectively, these data suggest that digital biomarkers were significantly different across time periods within each person leading us to believe that individualized ML models would be able to accurately discriminate between PPD and other time periods involved with pregnancy.

Our study also highlights the strength of utilizing individualized N-of-1 ML models using digital biomarkers for identifying PPD. Our findings underscored the models’ ability to differentiate between distinct pregnancy phases—namely, pre-pregnancy, pregnancy, postpartum, and PPD time periods (Figure 3 and Supplementary Figure 1). Notably, our approach’s validity was confirmed by the noticeable decrease in model performance during the PPD-equivalent time period for the non-PPD cohort compared to the PPD time period within the PPD cohort (Figures 4 and 5). This observation demonstrated the distinct behavioral shifts that are observed during the onset of PPD, effectively captured by digital biomarkers^2^. Furthermore, our results did not indicate a significant variation in individualized model performance across the four pregnancy time periods among women with a history of depression prior to or during pregnancy (Supplementary Figure 2). This accentuated the robust capability of individualized models to differentiate between time periods based on the distinct behavioral characteristics and metabolic shifts linked to PPD post-pregnancy, as opposed to the behavioral changes exhibited by each woman before pregnancy or during pregnancy. This suggests that forthcoming technology centered around detecting PPD through digital biomarkers could have relevance for both individuals with and without a pre-existing history of depression before and/or during pregnancy. Future studies should be conducted in a prospective framework to validate our individualized methodology.

Another crucial finding from our study revealed that the vital digital biomarkers for PPD classification were calories BMR, average HR, Q1 HR, minimum HR, and lightly active minutes, where calories BMR was the most predictive feature (Figures 6A and 6B). Therefore, we constructed SHAP dependence plots to enhance our understanding of the relationship between calories BMR and PPD. Plots during pre-pregnancy/PPD suggested that elevated calories BMR is predictive of PPD, which could be attributed to a variety of factors, such as heightened stress levels or metabolic alterations associated with PPD (Figure 7A)^2,34^. During pregnancy/PPD and postpartum/PPD time periods, the relationship between Shapley and actual values of calories BMR flipped, signifying an increased number of calories BMR was inversely associated with PPD (Figure 7A). The negative relationship in the context of pregnancy/PPD can likely be derived from the metabolic changes during pregnancy, resulting in an increased basal metabolic rate^35^. In the context of postpartum/PPD, we speculate that the negative relationship is because the median duration between the delivery date and PPD diagnosis is 83 days, where patients may not have fully returned to their pre-pregnancy physiological or behavioral patterns, which can take up to six months^36^. As a result, the relationship between Shapley values and actual values of calories BMR may reflect this transitional period and the ongoing postpartum changes experienced by women.

On the other hand, during the pre-pregnancy/PPD-equivalent time periods for women in the non-PPD cohort, SHAP dependence plots failed to unveil a uniform connection between calories BMR and the PPD-equivalent time period, likely due to physiological distinctions, lifestyle changes during pregnancy, and random dissimilarities among women^23,37–39^. However, the comparison of SHAP dependence plots across pregnancy/PPD-equivalent and postpartum/PPD-equivalent time periods for women within the non-PPD cohort exhibited a consistent negative correlation, similar to what was observed in the PPD cohort (Figures 7A and 7B). This trend is likely a result of the common occurrence of an increased basal metabolic rate during pregnancy^35^. In the context of the postpartum/PPD-equivalent time periods, our utilization of an index date set at 83 days after delivery – the median number of days after delivery for PPD diagnosis in the PPD cohort – implies that women likely have not fully returned to their pre-pregnancy physiological baseline^36^. This aligns with the parallel observation seen during the pregnancy/PPD-equivalent time periods, reaffirming the persisting metabolic effect postpartum.

In the PPD cohort, the SHAP dependence plots for average HR, Q1 HR, and minimum HR during pregnancy/PPD also demonstrated a negative relationship indicating that higher values of these digital biomarkers are inversely associated with PPD (Figure 7A). This relationship may be ascribed to the elevated heart rate commonly observed during pregnancy, which is a physiological response resulting from vascular remodeling for promoting augmented blood flow to the uterus^40–42^. Additionally, there was a positive correlation between the increase in lightly active minutes and the occurrence of PPD in the pregnancy/PPD time periods, which may be explained by an inverse relationship between lightly active minutes and fairly active minutes/very active minutes (Supplementary Figure 5). Specifically, a higher number of lightly active minutes is concomitant with a decrease in the amount of time spent in fairly active and very active physical activities, aligning with the well-established understanding that reductions in overall physical activity can contribute to an increase in depressive symptoms^43^. In contrast, among women without PPD, a notable correlation was found solely in the pre-pregnancy/PPD-equivalent time periods concerning minimum HR, where an elevation in minimum HR was linked to the PPD-equivalent time period (Figure 7B). Although a subset of women demonstrated a significant correlation in SHAP dependence plots concerning digital biomarkers of average HR, Q1 HR, or lightly active minutes across pre-pregnancy/PPD-equivalent, pregnancy/PPD-equivalent, or postpartum/PPD-equivalent time periods, the overall proportion of women exhibiting such patterns was insufficient to draw definitive conclusions regarding the relationship between digital biomarkers during the pre-pregnancy, pregnancy, or postpartum time periods compared to the PPD-equivalent time period (Figure 7B). We postulate that the contrasting patterns of digital biomarkers among women in the PPD and non-PPD cohorts imply potential differences in these biomarkers for women who eventually experience PPD. Therefore, it may become plausible to utilize ML models during pre-pregnancy or pregnancy time periods to predict a woman’s risk for future PPD onset.

In general, prior investigations have adhered to conventional ML strategies, revolving around the development of a solitary model. In this paradigm, a model is constructed employing an extensive patient dataset encompassing individuals exhibiting either continuous outcomes (for regression-based models) or categorical outcomes. Subsequently, when a new patient is introduced, the model generates predictions for the patient based on their data and the pre-established model^44^. While this approach carries advantages, it is beset by two primary limitations: 1) reliance on an ample sample size and 2) neglecting to accommodate the diverse and heterogeneous spectrum of depressive symptoms^16^. Hence, a captivating domain of exploration has honed in on crafting intra-individual ML models within the realm of depression. This advancement tackles the constraints of conventional approaches in two ways: first, it sidesteps the need for an extensive sample size, given that the model is solely tailored to a single patient’s data; and second, it conscientiously acknowledges the heterogeneous spectrum of depressive symptoms through a focused evaluation of the unique behaviors exhibited by that specific patient.

The use of individualized models may serve as a superior preference compared to those formulated using cohort-based methodologies. For instance, a cross-sectional study using traditional ML models from Fitbit data from healthy adults to predict depression severity only displayed a moderate AUC range of 0.51-0.66. Moreover, while the results demonstrated commendable specificity (0.98-1), sensitivity exhibited marked inadequacy (0.03-0.13)^45^. Another study aimed to investigate the potential of ML models utilizing digital biomarkers in distinguishing between patients with unipolar and bipolar depression against healthy controls. However, the most successful model exhibited an accuracy rate of 0.73 (73%) and a kappa value of 0.44, which doesn’t qualify as a notably high-performing model^46,47^. Additional investigations have also materialized within a cohort-based framework; nevertheless, these studies grapple with a noteworthy drawback – they incorporated patient mood as a predictive feature in their models. Considering that these studies aimed to predict the severity of depression, it’s unsurprising that these models exhibited heightened performance levels^13,48^.

In order to effectively underscore the viability of personalized ML models over cohort-based methods, our study directly juxtaposed the performance of both approaches (individualized versus cohort-based ML models) side-by-side. Notably, our findings vividly showcased the superior performance achieved through the personalized methodology for the PPD class (average sensitivity = 0.78, specificity = 0.95, precision = 0.84, recall = 0.78, F1 score = 0.81) in comparison to conventional techniques with a cohort-based model (sensitivity = 0.54, specificity = 0.55, precision = 0.49, recall = 0.54, F1 score = 0.52), leveraging digital biomarkers from Fitbit for PPD detection (Figure 8). This outcome accentuated that individualized models present an encouraging avenue for crafting ML models aimed at identifying mood disorders.

Although this study provides a strong foundation for using digital biomarkers to classify PPD, it is not without limitations. First, the study faced constraints due to the restricted number of patients available, which hindered the implementation of conventional ML techniques. However, due to the limited sample size, we opted for an individualized approach, which not only addressed the small sample size but also provided a means to accommodate the inherent variability within individuals^19^. Second, the process of phenotyping PPD patients relied on a PPD diagnosis or medication usage, which could potentially lack specificity in diagnostic codes and miss undiagnosed cases. Third, our approach assumed a standard pregnancy length of nine months, which may not always align with individual variations. Fourth, there are several layers of confounding that occur during the different phases of pregnancy that may indirectly influence digital biomarkers and ML models, especially as it relates to PPD classification, such as 1) significant hormonal changes that impact physical and mental states; 2) metabolic changes that occur as a result of pregnancy; 3) increased levels of stress during pregnancy and the postpartum period; 4) modifications to one’s lifestyle, such as food consumption, during pregnancy and the postpartum period; and 5) alterations in physical activity during the postpartum period as a result of birthing complications^49–53^. Fifth, this study excluded patients with chronic conditions to mitigate the potential influence of those conditions on digital biomarkers. Sixth, sleep data was absent in the AoURP dataset at the time of this analysis, although it might also hold predictive value for PPD. Furthermore, it may be beneficial for subsequent analyses to account for features like seasonal variation that may also indirectly influence behavior^54^.

Overall, the findings from this study suggest it is feasible to characterize PPD in addition to other time periods of pregnancy using passively collected digital biomarkers from consumer-grade wearables. The development of individualized models allows for a personalized approach to capture behavioral differences in the form of digital biomarkers. This research lays a robust foundation for forthcoming applications aimed at enhancing the early detection of PPD, a condition that is often underdiagnosed and undertreated. Moreover, on a broader scale, it indicates the exciting potential for intra-individual ML models to be extended to various health conditions.

## Materials and Methods

### Data source and platform

This study uses the *All of Us* Research Program (AoURP) Registered Tier Dataset v6. Study analysis was performed using the AoURP Researcher workbench cloud platform. All computational phenotyping, data processing, data analysis, and ML were conducted using R. The daily average HR, standard deviation HR, minimum HR, Q1 HR, median HR, Q3 HR, and maximum HR were calculated using the Fitbit Heart Rate Level table. The sum of steps was calculated using the Fitbit Intra Day Steps table. Activity calories, calories BMR, calories out, fairly active minutes, lightly active minutes, marginal calories, sedentary minutes, and very active minutes were taken from the Fitbit Activity Summary table.

### Measures to protect patient privacy

In compliance with the Data and Statistics Dissemination Policy of the AoURP, counts of less than 20 cannot be presented to mitigate the risk of patient re-identification^55^. Since the cohort of patients with PPD presented in this analysis consists of less than 20 patients, percentages were presented as percent ranges (e.g., instead of presenting 53%, the data was presented as 50-55%). Publication of results in this manner has been approved by the AoURP Resource Access Board (RAB). Furthermore, race and ethnicity were not reported due to the limited sample size as requested by the AoURP RAB.

### Computational phenotyping

#### Identifying women with postpartum depression (PPD)

Women with PPD were identified in the following three-fold approach: 1) selecting women with a diagnosis of PPD using the condition data; identifying women with a record of 2) pregnancy or 3) delivery, who have been diagnosed with depression and/or have antidepressant drug exposure during the postpartum period.

The first branch of the three-fold approach to creating a cohort of women with PPD was performed using Observational Medical Outcomes Partnership (OMOP) concept IDs in the Condition table based on the Observational Health Data Science and Informatics (OHDSI) initiative in Supplementary Table 7^56^. For both the second and third branches of the method, we first identified women with a record of delivery (using condition data) or pregnancy (using Condition and Survey tables) based on concept IDs from previously published work in Supplementary Table 7^57^. Next, the data was filtered on the earliest record of delivery/pregnancy to capture and analyze digital biomarker data during the pre-pregnancy period. To estimate the date of pregnancy or delivery (depending on which was available for that individual), the date observed in the electronic health record (EHR) from the AoURP was adjusted by adding or subtracting nine months, which is a typical length of pregnancy^58^. Our next step was to estimate the window of the postpartum period to monitor depressive symptoms, which was defined as starting from the date of delivery and spanning 24 months after that date^59,60^. Consistent with other EHR computational phenotyping studies of PPD, individuals were also classified as being PPD positive if they had a diagnosis of depression in the condition table and/or antidepressant drug exposure within the postpartum window^61^ (Supplementary Table 7). Specific concepts containing the terms “episode”, “remission”, “reactive”, “atypical”, “premenstrual”, “schizoaffective”, and “seasonal” were excluded when identifying individuals with a depression diagnosis since they would not appropriately capture women with a persistent depression during the postpartum period. If a woman in the PPD cohort showed records of depression diagnosis and antidepressant drug exposure, we selected the earliest record to be considered the index date. For women with pregnancy and delivery data available, the index date and data used were based on the delivery record since this provided an elevated level of confidence in defining the postpartum period and, subsequently, whether the depression diagnosis/antidepressant drug exposure occurred during the postpartum period. Lastly, the final PPD cohort was generated by selecting unique women from each of the three branches of our approach.

#### Identifying women without PPD

To establish a cohort of women unaffected by PPD, we applied the identical rationale to the second and third branches of our PPD phenotyping, as described above. Subsequently, women with records indicating PPD or depression diagnosis during the postpartum period from the condition table, or any instances of antidepressant drug usage from the drug exposure table, were excluded.

### Data preparation for analysis and individualized ML models

To prepare the data for analysis and individualized ML models using wearable data, we first merged day-level data from the Fitbit (HR, steps, physical activity, and calories burned; see Supplementary Table 1 for more information on digital biomarkers) for each individual ranging from two years prior through 30 days after the index date to capture their behavior before, during, and after pregnancy. Previous studies have demonstrated that HR, steps, and activity measurements from Fitbit are fairly accurate and can be used for research purposes^62,63^. The decision to choose measures related to HR instead of resting HR was based on the availability of data and the consideration of having enough measurements for each individual to train ML models. Digital biomarker data was filtered on days of “compliant” data, which was characterized by 1) at least 10 hours of Fitbit wear time within a day and 2) between 100 and 45,000 steps, as seen in previous studies^64^. Individuals from the PPD cohort were excluded from individualized ML models if they possessed less than 50 days of total data.

### Statistical analysis

#### Linear mixed-effects models

The lme4 and lmerTest packages in R were utilized to construct hierarchical linear regression models, aiming to assess the presence of noteworthy differences among women and examine the relationship between each time period and digital biomarker^65,66^. To assess if there was a significant level of variation in digital biomarkers between individuals, we processed data to calculate the average value of each digital biomarker during each time period (e.g., average HR during pre-pregnancy, average HR during pregnancy, average HR during postpartum, average HR during PPD, etc.) and conducted linear mixed-effects models with person ID as the random effect. One model was built for each digital biomarker, where the digital biomarker served as the outcome variable, time period was considered the independent variable, and person ID was incorporated as a random effect. The presence of significant variability among individuals was evaluated using the performance package at a significance level of 0.05^67^.

#### Interrupted Time Series Analysis (ITSA), Tukey Honest Significance Differences (HSD) Test, and digital biomarker directionality assessment between time periods

The Interrupted Time Series Analysis (ITSA) was performed using the its.analysis package in R, employing a significance level of 0.05^68^. To compare whether there was a difference in digital biomarkers during different time periods before, during, and after pregnancy, in addition to when patients experienced postpartum depression, four time periods were defined for each individual identified with PPD (pre-pregnancy, pregnancy, postpartum without depression (hereafter referred to as postpartum), and postpartum with depression (PPD)). For each woman, a model was constructed for each digital biomarker, with 250 replications used for bootstrapping. The dependent variable was the digital biomarker value, the “time” parameter was the date, and the interrupting variable was the time period (pre-pregnancy, pregnancy, postpartum, PPD). The mean and standard deviation were calculated for each digital biomarker during each of the four time periods for each woman. Furthermore, a Tukey’s Honest Significant Differences (HSD) test was conducted to assess the statistical significance of the differences in each digital biomarker between each permutation of time periods (PPD - pre-pregnancy, PPD - pregnancy, PPD - postpartum, postpartum - pre-pregnancy, postpartum - pregnancy, and pregnancy - pre-pregnancy) within each individual at a significance level of 0.05^69^. Next, the percentage of women exhibiting a significant relationship was calculated for each digital biomarker in each group comparison (e.g., PPD - pre-pregnancy). To determine the overall trend in digital biomarker change between pairs of time periods (e.g., PPD and pre-pregnancy, PPD and pregnancy, PPD and postpartum, etc.), the average difference across all individuals was computed for each digital biomarker. This average also included non-significant differences, as they still contributed insights into the directionality of digital biomarkers during those time periods, even if the differences were not statistically significant. Lastly, a two-sided unpaired t-test at a significance level of 0.05 was conducted to assess the statistical significance of the net difference compared to zero, with positive change defined as an average value greater than zero and negative change defined as an average value less than zero.

### Building ML models

#### Individualized ML models for women in the PPD cohort

Individualized ML models were developed with the objective of determining the potential of digital biomarkers to differentiate among four distinct pregnancy phases, including pre-pregnancy, pregnancy, postpartum period without depression (i.e., postpartum), and postpartum period with depression (i.e., PPD). Therefore, multinomial models were developed with time period as the outcome with all 16 digital biomarkers as the features in the model (see Supplementary Table 1 for a list of digital biomarkers included). Initially, our intention was to examine the model’s capacity to discriminate between time periods with and without PPD, thereby constructing binomial classification models. However, we recognized the presence of repeated measurements (multiple days of data) during pre-pregnancy, pregnancy, and postpartum time frames. Consequently, due to the repetitive nature of our outcome measurements, we opted for constructing multinomial ML models to effectively discern among the four identified time periods, where the PPD time period is treated as both a time period and diagnosis.

To build intra-individual models, the data was filtered on each woman, where they were considered PPD negative ranging from two years prior through 15 days before the index date and PPD positive 14 days prior through 30 days after the index date. We selected 14 days preceding the index date as the first day of being positive for PPD, as the criteria for diagnosis states that patients must display five depressive symptoms lasting two weeks^22^. The timeframe of 30 days following the index date was chosen due to the fact that some individuals in the PPD cohort received antidepressant medication on the day of their diagnosis, which can begin to take effect after about four weeks of usage^70^. For each individual, the data were centered and scaled before building models using three repeats of 10-fold cross-validation using a tune length of five with random forest, generalized linear models (GLM), support vector machine (SVM), and K-nearest neighbors (KNN), as these algorithms have been used in previous studies assessing depression with wearables^15,71^. Models were built using the Caret package in R and evaluated using a combination of the Kappa statistic and multiclass AUC (referred to as mAUC), which are standard metrics for classification ML models^24,72–74^. Model performance for each time period was further assessed using a confusion matrix, which calculated sensitivity, specificity, precision, recall, and F1 score^74^.

#### Comparing individualized ML model performance between women with a history of depression before or during pregnancy

To initially ascertain the presence of depression history before or during pregnancy within the PPD cohort, we determined the date of delivery (utilizing condition data) or the date of pregnancy (employing condition and survey data) based on the concept IDs detailed in Supplementary Table 7^57^. Depending on the available data for each woman, the date of pregnancy was calculated by subtracting nine months from the date of delivery, whereas the date of delivery was calculated by adding nine months to the date of pregnancy, representing a standard pregnancy duration^58^. In cases where both delivery and pregnancy records existed, priority was given to the date of delivery due to its heightened reliability.

For the evaluation of individualized ML model performance within the PPD cohort, concerning women with a history of depression, the cohort was categorized into four subgroups encompassing 1) no prior depression history; 2) depression prior to pregnancy; 3) depression during pregnancy; and 4) depression both prior to and during pregnancy. To examine potential disparities in individualized ML model performance, a two-sided unpaired t-test was conducted with a significance threshold of 0.05. This analysis was executed to compare the no-depression history group with the groups of women exhibiting depression pre-pregnancy, during pregnancy, or both prior to and during pregnancy. Sensitivity, specificity, precision, recall, and F1 score metrics were subjected to this statistical comparison process.

#### Individualized ML models for women without PPD

To construct personalized models for women unaffected by PPD, we implemented an analogous approach to the one used for women in the PPD cohort. It is worth noting that women without PPD would not have a “fourth” time period (i.e., postpartum with depression in women with PPD), as they did not experience PPD. In order to ensure comparability and effectively gauge model performance between women with and without PPD, we devised a PPD-equivalent time period for the non-PPD cohort, mirroring the PPD time period. Considering that the median time to diagnose PPD was found to be 83 days following delivery, we ensured uniformity by setting the index date as 83 days after delivery. This index date was chosen to represent the PPD-equivalent time period for women in the non-PPD cohort. As we established an index date aligned with that of the PPD cohort, the interval of 14 days prior to the index date was not considered the PPD-equivalent time period for these women, as they did not actually experience PPD. Subsequently, individualized ML models were constructed in a manner akin to those in the PPD cohort using the RF algorithm (since this algorithm yielded optimal results in the PPD cohort), using three repetitions of 10-fold cross-validation and a tuning length of five. Similar to the approach developed for women in the PPD cohort, model performance was evaluated using sensitivity, specificity, precision, recall, and F1 score^73,74^. Models were not assessed by mAUC or kappa since model performance was only reduced in the PPD-equivalent time period and not in the pre-pregnancy, pregnancy, or postpartum time periods compared to those in the PPD cohort.

#### Comparing individualized ML model performance for women in the PPD and non-PPD cohorts

For comparing the performance of individualized ML models in the PPD cohort to the non-PPD cohort, we performed a two-sided unpaired t-test with a significance level of 0.05.

### Variable importance

#### SHAP approach

We used the RF ML models to generate a ranking of digital biomarkers for each individual since they had the best performance. Following that, Shapley values were computed for each measurement within each individualized model for the PPD class using the iml package in R^75^. To determine the feature ranking within individual models, we computed the average absolute value of Shapley values across all measurements for each digital biomarker and sorted the rankings from largest to smallest (see Supplementary Figure 3 for an example beeswarm plot from one woman in the PPD cohort). We then tallied the number of models in which each biomarker ranked among the top five most predictive for the PPD class to produce an overall ranking of digital biomarkers. Furthermore, we determined the most predictive feature of PPD by totaling the number of models in which each digital biomarker ranked as the top predictor for the PPD class.

#### Permutation approach

To enhance the robustness of our approach, variable importance was also computed using a permutation-based method in the Caret package in R^74^. Subsequently, the features were sorted based on the magnitude of values assigned for the variable importance regarding the PPD class. Employing a similar methodology as with SHAP, we tabulated the number of models in which each digital biomarker ranked among the top five most predictive for the PPD class, yielding a comprehensive ranking of digital biomarkers. The frequency with which each feature ranked as the foremost predictive digital biomarker was also recorded for the PPD class.

### SHAP dependence plots

SHAP dependence plots were generated using the gpplot2 package in R^76^. For each individual, plots were generated by graphing the Shapley value against the corresponding actual value for the digital biomarker. Given that the outcome of models was multinomial (pre-pregnancy, pregnancy, postpartum, or PPD), three separate SHAP dependence plots were generated for each individual using calories BMR data during PPD with one other time period (i.e., one plot for pre-pregnancy and PPD (referred to as pre-pregnancy/PPD), one plot for pregnancy and PPD (referred to as pregnancy/PPD), and one plot for postpartum and PPD (referred to as postpartum/PPD) to more easily analyze the relationship between calories BMR in a binomial context between PPD relative to one other time period (see Supplementary Figures 4A-C for an example from one woman for each time period). This process was repeated for women in the non-PPD cohort in a similar fashion to the PPD cohort, specifically PPD-equivalent versus pre-pregnancy (pre-pregnancy/PPD-equivalent), PPD-equivalent versus pregnancy (pregnancy/PPD-equivalent), and PPD-equivalent versus postpartum (postpartum/PPD-equivalent). The Pearson correlation coefficient and its corresponding p-value were computed at a significance level of 0.05, followed by calculating the percentages of women with and without a significant correlation. If a significant correlation was observed, we further determined its direction (positive or negative) and calculated the percentages of women with a positive or negative correlation. The overall consensus regarding the relationship was determined by comparing the percentage of positive and negative correlations for each digital biomarker across all individuals, thereby identifying which direction had a greater rate. In cases where the percentage of women with a significant correlation was less than 40%, the direction was not assessed due to the small sample size, which may not be representative of the population.

### Building an ML model for PPD using a cohort-based approach

For the construction of an ML model assessing whether or not a woman has PPD, our focus was on utilizing the PPD and PPD-equivalent time periods sourced from both the PPD and non-PPD cohorts. We proceeded to develop a binomial RF classification model, in which 75% of individuals from each cohort were designated for the training set, and the remaining 25% were assigned to the test set, utilizing the Caret package within R^74^. To ensure the reliability of model performance assessment, we diligently executed train and test set divisions based on individual person IDs, thereby preventing any overlap of women between the two sets that could potentially distort results^77^. The model’s target outcome pertained to a binary classification of whether an individual exhibited PPD or not, relying on all 16 digital biomarkers as input (refer to Supplementary Table 1 for a comprehensive description of the employed digital biomarkers). The data was normalized through centering and scaling procedures. Notably, repeated cross-validation was omitted due to the presence of repeated measurements stemming from various person IDs. The model’s construction integrated a tune length of five. The evaluation of the models was performed using the same metrics of kappa and AUC (in this instance, not multiclass since the outcome was binary). Subsequently, a confusion matrix was generated to calculate sensitivity, specificity, precision, recall, and F1 score^24,72–74^.

### Large language models

ChatGPT (GPT-3.5), developed by OpenAI (https://openai.com/) was used to edit some portions of the manuscript by offering synonym suggestions, language enhancements, grammar refinements, and style improvements. It’s important to note that all recommendations made by ChatGPT were meticulously reviewed by the author and were not utilized for generating ideas or content.

## Data Availability

The data utilized for this analysis can be accessed via the *All of Us* Researcher Workbench by individuals who have successfully completed the requisite training.

https://www.researchallofus.org/

## Acknowledgments

First and foremost, the *All of Us* Research Program would not be possible without the partnership of its participants. The *All of Us* Research Program is supported by the National Institutes of Health, Office of the Director: Regional Medical Centers: 1 OT2 OD026549; 1 OT2 OD026554; 1 OT2 OD026557; 1 OT2 OD026556; 1 OT2 OD026550; 1 OT2 OD 026552; 1 OT2 OD026553; 1 OT2 OD026548; 1 OT2 OD026551; 1 OT2 OD026555; IAA #: AOD 16037; Federally Qualified Health Centers: HHSN 263201600085U; Data and Research Center: 5 U2C OD023196; Biobank: 1 U24 OD023121; The Participant Center: U24 OD023176; Participant Technology Systems Center: 1 U24 OD023163; Communications and Engagement: 3 OT2 OD023205; 3 OT2 OD023206; and Community Partners: 1 OT2 OD025277; 3 OT2 OD025315; 1 OT2 OD025337; 1 OT2 OD025276.

