## Supplementary Tables 1-6 and Supplementary Figures 1-5 for "Harnessing consumer wearable digital biomarkers for individualized recognition of postpartum depression using the *All of Us* Research Program dataset"

### Supplementary Tables and Figures

**Supplementary Table 1: Digital biomarkers measured by the Fitbit available in the *All of Us* Research Program (AoURP) dataset v6.**

| Name | Category | Description |
|---|---|---|
| Average HR | Heart rate | Mean heart rate per date |
| SD HR | Heart rate | Heart rate’s standard deviation per date |
| Minimum HR | Heart rate | Minimum heart rate per date |
| Q1 HR | Heart rate | Heart rate’s first quartile per date |
| Median HR | Heart rate | Median heart rate per date |
| Q3 HR | Heart rate | Heart rate’s third quartile per date |
| Maximum HR | Heart rate | Maximum heart rate per date |
| Sum steps | Activity | Sum of steps per date |
| Activity calories | Activity | Sum of calories burned when not at rest per date |
| Calories BMR | Activity | Sum of calories burned at rest per date |
| Calories out | Activity | Sum of total calories burned per date |
| Fairly active minutes | Activity | Sum of fairly active minutes per date |
| Lightly active minutes | Activity | Sum of lightly active minutes per date |
| Marginal calories | Activity | Difference in calories burned at rest and during activity per date |
| Sedentary minutes | Activity | Sum of sedentary minutes per date |
| Very active minutes | Activity | Sum of very active minutes per date |

**Supplementary Table 2: Random effect p-value from linear mixed effects model with person ID as the random effect.**

| Digital biomarker | Degrees of freedom | p-value |
|---|---|---|
| Average HR | 1 | 0.00E+00 |
| SD HR | 1 | 0.00E+00 |
| Minimum HR | 1 | 0.00E+00 |
| Q1 HR | 1 | 0.00E+00 |
| Median HR | 1 | 0.00E+00 |
| Q3 HR | 1 | 0.00E+00 |
| Maximum HR | 1 | 5.78E-164 |
| Sum steps | 1 | 0.00E+00 |
| Activity calories | 1 | 0.00E+00 |
| Calories BMR | 1 | 0.00E+00 |
| Calories out | 1 | 0.00E+00 |
| Fairly active minutes | 1 | 0.00E+00 |
| Lightly active minutes | 1 | 0.00E+00 |
| Marginal calories | 1 | 0.00E+00 |
| Sedentary minutes | 1 | 3.22E-285 |
| Very active minutes | 1 | 0.00E+00 |

*Statistical tests were run at a significance level of 0.05.

**Supplementary Table 3: Percentage of women with a significant difference between time periods from the interrupted time series analysis (ITSA).**

| Digital biomarker | Percent significant |
|---|---|
| Average HR | 65-70 |
| SD HR | 75-80 |
| Minimum HR | 65-70 |
| Q1 HR | 70-75 |
| Median HR | 65-70 |
| Q3 HR | 70-75 |
| Maximum HR | 65-70 |
| Sum steps | 65-70 |
| Activity calories | 75-80 |
| Calories BMR | 50-55 |
| Calories out | 65-70 |
| Fairly active minutes | 75-80 |
| Lightly active minutes | 75-80 |
| Marginal calories | 75-80 |
| Sedentary minutes | 60-65 |
| Very active minutes | 60-65 |

*Statistical tests were run at a significance level of 0.05.

**Supplementary Table 4: Percentage of women with a significant difference between all time periods from Tukey HSD tests.**

| Digital biomarker | Group | Percent significant |
|---|---|---|
| Average HR | PPD-Pre_pregnancy | 55-60 |
| Average HR | PPD-Pregnancy | 75-80 |
| Average HR | PPD-Postpartum | 50-55 |
| Average HR | Postpartum-Pre_pregnancy | 50-55 |
| Average HR | Postpartum-Pregnancy | 80-85 |
| Average HR | Pregnancy-Pre_pregnancy | 90-95 |
| SD HR | PPD-Pre_pregnancy | 60-65 |
| SD HR | PPD-Pregnancy | 50-55 |
| SD HR | PPD-Postpartum | 25-30 |
| SD HR | Postpartum-Pre_pregnancy | 75-80 |
| SD HR | Postpartum-Pregnancy | 30-35 |
| SD HR | Pregnancy-Pre_pregnancy | 90-95 |
| Minimum HR | PPD-Pre_pregnancy | 55-60 |
| Minimum HR | PPD-Pregnancy | 50-55 |
| Minimum HR | PPD-Postpartum | 50-55 |
| Minimum HR | Postpartum-Pre_pregnancy | 60-65 |
| Minimum HR | Postpartum-Pregnancy | 90-95 |
| Minimum HR | Pregnancy-Pre_pregnancy | 95-100 |
| Q1 HR | PPD-Pre_pregnancy | 45-50 |
| Q1 HR | PPD-Pregnancy | 65-70 |
| Q1 HR | PPD-Postpartum | 50-55 |
| Q1 HR | Postpartum-Pre_pregnancy | 35-40 |
| Q1 HR | Postpartum-Pregnancy | 80-85 |
| Q1 HR | Pregnancy-Pre_pregnancy | 95-100 |
| Median HR | PPD-Pre_pregnancy | 35-40 |
| Median HR | PPD-Pregnancy | 75-80 |
| Median HR | PPD-Postpartum | 45-50 |
| Median HR | Postpartum-Pre_pregnancy | 35-40 |
| Median HR | Postpartum-Pregnancy | 80-85 |
| Median HR | Pregnancy-Pre_pregnancy | 95-100 |
| Q3 HR | PPD-Pre_pregnancy | 45-50 |

| | | |
|---|---|---|
| Q3 HR | PPD-Pregnancy | 75-80 |
| Q3 HR | PPD-Postpartum | 40-45 |
| Q3 HR | Postpartum-Pre_pregnancy | 35-40 |
| Q3 HR | Postpartum-Pregnancy | 80-85 |
| Q3 HR | Pregnancy-Pre_pregnancy | 90-95 |
| Maximum HR | PPD-Pre_pregnancy | 60-65 |
| Maximum HR | PPD-Pregnancy | 60-65 |
| Maximum HR | PPD-Postpartum | 25-30 |
| Maximum HR | Postpartum-Pre_pregnancy | 35-40 |
| Maximum HR | Postpartum-Pregnancy | 40-45 |
| Maximum HR | Pregnancy-Pre_pregnancy | 35-40 |
| Sum steps | PPD-Pre_pregnancy | 80-85 |
| Sum steps | PPD-Pregnancy | 65-70 |
| Sum steps | PPD-Postpartum | 20-25 |
| Sum steps | Postpartum-Pre_pregnancy | 60-65 |
| Sum steps | Postpartum-Pregnancy | 40-45 |
| Sum steps | Pregnancy-Pre_pregnancy | 80-85 |
| Activity calories | PPD-Pre_pregnancy | 60-65 |
| Activity calories | PPD-Pregnancy | 60-65 |
| Activity calories | PPD-Postpartum | 45-50 |
| Activity calories | Postpartum-Pre_pregnancy | 60-65 |
| Activity calories | Postpartum-Pregnancy | 50-55 |
| Activity calories | Pregnancy-Pre_pregnancy | 90-95 |
| Calories BMR | PPD-Pre_pregnancy | 90-95 |
| Calories BMR | PPD-Pregnancy | 85-90 |
| Calories BMR | PPD-Postpartum | 60-65 |
| Calories BMR | Postpartum-Pre_pregnancy | 95-100 |
| Calories BMR | Postpartum-Pregnancy | 90-95 |
| Calories BMR | Pregnancy-Pre_pregnancy | 80-85 |
| Calories out | PPD-Pre_pregnancy | 60-65 |
| Calories out | PPD-Pregnancy | 60-65 |
| Calories out | PPD-Postpartum | 40-45 |
| Calories out | Postpartum-Pre_pregnancy | 35-40 |
| Calories out | Postpartum-Pregnancy | 50-55 |
| Calories out | Pregnancy-Pre_pregnancy | 60-65 |

| | | |
|---|---|---|
| Fairly active minutes | PPD-Pre_pregnancy | 80-85 |
| Fairly active minutes | PPD-Pregnancy | 50-55 |
| Fairly active minutes | PPD-Postpartum | 25-30 |
| Fairly active minutes | Postpartum-Pre_pregnancy | 60-65 |
| Fairly active minutes | Postpartum-Pregnancy | 40-45 |
| Fairly active minutes | Pregnancy-Pre_pregnancy | 55-60 |
| Lightly active minutes | PPD-Pre_pregnancy | 70-75 |
| Lightly active minutes | PPD-Pregnancy | 60-65 |
| Lightly active minutes | PPD-Postpartum | 30-35 |
| Lightly active minutes | Postpartum-Pre_pregnancy | 35-40 |
| Lightly active minutes | Postpartum-Pregnancy | 50-55 |
| Lightly active minutes | Pregnancy-Pre_pregnancy | 70-75 |
| Marginal calories | PPD-Pre_pregnancy | 60-65 |
| Marginal calories | PPD-Pregnancy | 65-70 |
| Marginal calories | PPD-Postpartum | 25-30 |
| Marginal calories | Postpartum-Pre_pregnancy | 60-65 |
| Marginal calories | Postpartum-Pregnancy | 50-55 |
| Marginal calories | Pregnancy-Pre_pregnancy | 90-95 |
| Sedentary minutes | PPD-Pre_pregnancy | 25-30 |
| Sedentary minutes | PPD-Pregnancy | 30-35 |
| Sedentary minutes | PPD-Postpartum | 25-30 |
| Sedentary minutes | Postpartum-Pre_pregnancy | 75-80 |
| Sedentary minutes | Postpartum-Pregnancy | 40-45 |
| Sedentary minutes | Pregnancy-Pre_pregnancy | 45-50 |
| Very active minutes | PPD-Pre_pregnancy | 70-75 |
| Very active minutes | PPD-Pregnancy | 45-50 |
| Very active minutes | PPD-Postpartum | 10-15 |
| Very active minutes | Postpartum-Pre_pregnancy | 60-65 |
| Very active minutes | Postpartum-Pregnancy | 30-35 |
| Very active minutes | Pregnancy-Pre_pregnancy | 55-60 |

*Statistical tests were run at a significance level of 0.05.

**Supplementary Table 5: Mean difference and direction of digital biomarkers between all time periods.**

| Digital biomarker | Group | Mean difference | SD difference | p-value | Net direction |
|---|---|---|---|---|---|
| Average HR | PPD-Pre_pregnancy | -0.02 | 2.99 | 0.98 | Not significant |
| Average HR | PPD-Pregnancy | -4.76 | 4.82 | 0 | Negative |
| Average HR | PPD-Postpartum | -0.56 | 3.45 | 0.54 | Not significant |
| Average HR | Postpartum-Pre_pregnancy | -1.58 | 3.66 | 0.26 | Not significant |
| Average HR | Postpartum-Pregnancy | -3.9 | 5.46 | 0.05 | Not significant |
| Average HR | Pregnancy-Pre_pregnancy | 4.97 | 4.86 | 0.01 | Positive |
| SD HR | PPD-Pre_pregnancy | -1.76 | 1.58 | 0 | Negative |
| SD HR | PPD-Pregnancy | -0.47 | 1.14 | 0.16 | Not significant |
| SD HR | PPD-Postpartum | 0.09 | 1.17 | 0.78 | Not significant |
| SD HR | Postpartum-Pre_pregnancy | -1.82 | 1.72 | 0.02 | Negative |
| SD HR | Postpartum-Pregnancy | -0.45 | 1.34 | 0.31 | Not significant |
| SD HR | Pregnancy-Pre_pregnancy | -1.18 | 1.28 | 0.01 | Negative |
| Minimum HR | PPD-Pre_pregnancy | 1.08 | 2.56 | 0.19 | Not significant |
| Minimum HR | PPD-Pregnancy | -4.02 | 4.32 | 0.01 | Negative |
| Minimum HR | PPD-Postpartum | -0.27 | 3.73 | 0.79 | Not significant |
| Minimum HR | Postpartum-Pre_pregnancy | 0.08 | 4.23 | 0.96 | Not significant |
| Minimum HR | Postpartum-Pregnancy | -3.26 | 5.53 | 0.1 | Not significant |
| Minimum HR | Pregnancy-Pre_pregnancy | 4.94 | 3.96 | 0 | Positive |
| Q1 HR | PPD-Pre_pregnancy | 1.34 | 3.06 | 0.18 | Not significant |
| Q1 HR | PPD-Pregnancy | -4.41 | 5 | 0.01 | Negative |
| Q1 HR | PPD-Postpartum | -0.55 | 3.54 | 0.56 | Not significant |
| Q1 HR | Postpartum-Pre_pregnancy | -0.24 | 3.71 | 0.86 | Not significant |
| Q1 HR | Postpartum-Pregnancy | -3.61 | 5.65 | 0.07 | Not significant |
| Q1 HR | Pregnancy-Pre_pregnancy | 5.81 | 5.07 | 0 | Positive |
| Median HR | PPD-Pre_pregnancy | 0.59 | 2.88 | 0.51 | Not significant |
| Median HR | PPD-Pregnancy | -4.43 | 5.33 | 0.01 | Negative |
| Median HR | PPD-Postpartum | -0.63 | 3.41 | 0.49 | Not significant |
| Median HR | Postpartum-Pre_pregnancy | -0.97 | 3.21 | 0.42 | Not significant |
| Median HR | Postpartum-Pregnancy | -3.64 | 5.92 | 0.08 | Not significant |
| Median HR | Pregnancy-Pre_pregnancy | 5.46 | 4.89 | 0 | Positive |

| | | | | | |
|---|---|---|---|---|---|
| Q3 HR | PPD-Pre_pregnancy | -0.75 | 2.97 | 0.42 | Not significant |
| Q3 HR | PPD-Pregnancy | -4.94 | 5.08 | 0 | Negative |
| Q3 HR | PPD-Postpartum | -0.65 | 3.63 | 0.5 | Not significant |
| Q3 HR | Postpartum-Pre_pregnancy | -2.22 | 3.59 | 0.12 | Not significant |
| Q3 HR | Postpartum-Pregnancy | -4.01 | 5.83 | 0.06 | Not significant |
| Q3 HR | Pregnancy-Pre_pregnancy | 4.69 | 4.9 | 0.01 | Positive |
| Maximum HR | PPD-Pre_pregnancy | -5.95 | 9.11 | 0.06 | Not significant |
| Maximum HR | PPD-Pregnancy | -5.36 | 6.95 | 0.02 | Negative |
| Maximum HR | PPD-Postpartum | 0.44 | 7.08 | 0.81 | Not significant |
| Maximum HR | Postpartum-Pre_pregnancy | -7.29 | 7.75 | 0.03 | Negative |
| Maximum HR | Postpartum-Pregnancy | -4.89 | 6.34 | 0.04 | Negative |
| Maximum HR | Pregnancy-Pre_pregnancy | -0.01 | 5.77 | 0.99 | Not significant |
| Sum steps | PPD-Pre_pregnancy | -3659.25 | 2266.34 | 0 | Negative |
| Sum steps | PPD-Pregnancy | -1633.4 | 1920.59 | 0.01 | Negative |
| Sum steps | PPD-Postpartum | 38.29 | 1182.07 | 0.9 | Not significant |
| Sum steps | Postpartum-Pre_pregnancy | -3357.43 | 1431.2 | 0 | Negative |
| Sum steps | Postpartum-Pregnancy | -1262.69 | 1477.86 | 0.02 | Negative |
| Sum steps | Pregnancy-Pre_pregnancy | -1694.59 | 1328.11 | 0 | Negative |
| Activity calories | PPD-Pre_pregnancy | -295.71 | 239.34 | 0 | Negative |
| Activity calories | PPD-Pregnancy | -81.06 | 183.73 | 0.14 | Not significant |
| Activity calories | PPD-Postpartum | -23.84 | 157.73 | 0.57 | Not significant |
| Activity calories | Postpartum-Pre_pregnancy | -270.1 | 214.75 | 0.01 | Negative |
| Activity calories | Postpartum-Pregnancy | -57.63 | 194.46 | 0.37 | Not significant |
| Activity calories | Pregnancy-Pre_pregnancy | -178.29 | 162.57 | 0 | Negative |
| Calories BMR | PPD-Pre_pregnancy | 19.97 | 93.84 | 0.5 | Not significant |
| Calories BMR | PPD-Pregnancy | 7.23 | 62.83 | 0.69 | Not significant |
| Calories BMR | PPD-Postpartum | -4.55 | 38.84 | 0.66 | Not significant |
| Calories BMR | Postpartum-Pre_pregnancy | 14.92 | 118.06 | 0.73 | Not significant |
| Calories BMR | Postpartum-Pregnancy | 6.52 | 43.23 | 0.64 | Not significant |
| Calories BMR | Pregnancy-Pre_pregnancy | 14.9 | 95.82 | 0.62 | Not significant |
| Calories out | PPD-Pre_pregnancy | -217.82 | 206.13 | 0.01 | Negative |
| Calories out | PPD-Pregnancy | -70.08 | 141.95 | 0.1 | Not significant |
| Calories out | PPD-Postpartum | -25.75 | 132.44 | 0.46 | Not significant |
| Calories out | Postpartum-Pre_pregnancy | -208.54 | 273.36 | 0.07 | Not significant |
| Calories out | Postpartum-Pregnancy | -49.87 | 180.69 | 0.41 | Not significant |

| | | | | | |
|---|---|---|---|---|---|
| Calories out | Pregnancy-Pre_pregnancy | -116.69 | 181.65 | 0.06 | Not significant |
| Fairly active minutes | PPD-Pre_pregnancy | -12.98 | 12.21 | 0.01 | Negative |
| Fairly active minutes | PPD-Pregnancy | -6.47 | 8.15 | 0.01 | Negative |
| Fairly active minutes | PPD-Postpartum | -0.73 | 7.95 | 0.73 | Not significant |
| Fairly active minutes | Postpartum-Pre_pregnancy | -13.21 | 13.37 | 0.03 | Negative |
| Fairly active minutes | Postpartum-Pregnancy | -4.82 | 5.67 | 0.02 | Negative |
| Fairly active minutes | Pregnancy-Pre_pregnancy | -6.74 | 8.01 | 0.02 | Negative |
| Lightly active minutes | PPD-Pre_pregnancy | -34.38 | 56.84 | 0.07 | Not significant |
| Lightly active minutes | PPD-Pregnancy | 6.4 | 50.3 | 0.65 | Not significant |
| Lightly active minutes | PPD-Postpartum | -2.46 | 37.7 | 0.8 | Not significant |
| Lightly active minutes | Postpartum-Pre_pregnancy | -32.2 | 40.76 | 0.06 | Not significant |
| Lightly active minutes | Postpartum-Pregnancy | 5.41 | 48.64 | 0.73 | Not significant |
| Lightly active minutes | Pregnancy-Pre_pregnancy | -29.45 | 27.63 | 0.01 | Negative |
| Marginal calories | PPD-Pre_pregnancy | -195.21 | 143.91 | 0 | Negative |
| Marginal calories | PPD-Pregnancy | -70.76 | 100.28 | 0.03 | Negative |
| Marginal calories | PPD-Postpartum | -15.2 | 88.55 | 0.52 | Not significant |
| Marginal calories | Postpartum-Pre_pregnancy | -180.45 | 144.17 | 0.01 | Negative |
| Marginal calories | Postpartum-Pregnancy | -49.62 | 108.47 | 0.18 | Not significant |
| Marginal calories | Pregnancy-Pre_pregnancy | -108.13 | 107.12 | 0.01 | Negative |
| Sedentary minutes | PPD-Pre_pregnancy | 50.68 | 55.36 | 0.01 | Positive |
| Sedentary minutes | PPD-Pregnancy | 21.65 | 70.09 | 0.29 | Not significant |
| Sedentary minutes | PPD-Postpartum | -24.22 | 74.85 | 0.23 | Not significant |
| Sedentary minutes | Postpartum-Pre_pregnancy | 102.6 | 95.3 | 0.02 | Positive |
| Sedentary minutes | Postpartum-Pregnancy | 70.59 | 68.82 | 0.01 | Positive |
| Sedentary minutes | Pregnancy-Pre_pregnancy | 37.19 | 81.98 | 0.16 | Not significant |
| Very active minutes | PPD-Pre_pregnancy | -11.67 | 9.09 | 0 | Negative |
| Very active minutes | PPD-Pregnancy | -6.02 | 6.81 | 0.01 | Negative |
| Very active minutes | PPD-Postpartum | 0.08 | 3.97 | 0.94 | Not significant |
| Very active minutes | Postpartum-Pre_pregnancy | -10.62 | 7.67 | 0.01 | Negative |
| Very active minutes | Postpartum-Pregnancy | -5.37 | 6.11 | 0.02 | Negative |
| Very active minutes | Pregnancy-Pre_pregnancy | -5.25 | 5.75 | 0.01 | Negative |

*Statistical tests were run at a significance level of 0.05.

**Supplementary Table 6: Net direction of digital biomarkers between pairs of time periods.**

| Group | Direction | Digital biomarker |
|---|---|---|
| PPD-Pre_pregnancy | Positive | Sedentary minutes |
| PPD-Pre_pregnancy | Negative | SD HR, Sum steps, Activity calories, Calories out, Fairly active minutes, Marginal calories, Very active minutes |
| PPD-Pre_pregnancy | Not significant | Average HR, Minimum HR, Q1 HR, Median HR, Q3 HR, Maximum HR, Calories BMR, Lightly active minutes |
| PPD-Pregnancy | Negative | Average HR, Minimum HR, Q1 HR, Median HR, Q3 HR, Maximum HR, Sum steps, Fairly active minutes, Marginal calories, Very active minutes |
| PPD-Pregnancy | Not significant | SD HR, Activity calories, Calories BMR, Calories out, Lightly active minutes, Sedentary minutes |
| PPD-Postpartum | Positive | None |
| PPD-Postpartum | Negative | None |
| PPD-Postpartum | Not significant | Average HR, SD HR, Minimum HR, Q1 HR, Median HR, Q3 HR, Maximum HR, Sum steps, Activity calories, Calories BMR, Calories out, Fairly active minutes, Lightly active minutes, Marginal calories, Sedentary minutes, Very active minutes |
| Postpartum-Pre_pregnancy | Positive | Sedentary minutes |
| Postpartum-Pre_pregnancy | Negative | SD HR, Maximum HR, Sum steps, Activity calories, Fairly active minutes, Marginal calories, Very active minutes |
| Postpartum-Pre_pregnancy | Not significant | Average HR, Minimum HR, Q1 HR, Median HR, Q3 HR, Calories BMR, Calories out, Lightly active minutes |
| Postpartum-Pregnancy | Positive | Sedentary minutes |
| Postpartum-Pregnancy | Negative | Maximum HR, Sum steps, Fairly active minutes, Very active minutes |
| Postpartum-Pregnancy | Not significant | Average HR, SD HR, Minimum HR, Q1 HR, Median HR, Q3 HR, Activity calories, Calories BMR, Calories out, Lightly active minutes, Marginal calories |
| Pregnancy-Pre_pregnancy | Positive | Average HR, Minimum HR, Q1 HR, Median HR, Q3 HR |
| Pregnancy-Pre_pregnancy | Negative | SD HR, Sum steps, Activity calories, Fairly active minutes, Lightly active minutes, Marginal calories, Very active minutes |
| Pregnancy-Pre_pregnancy | Not significant | Maximum HR, Calories BMR, Calories out, Sedentary minutes |

*Statistical tests were run at a significance level of 0.05.

**Supplementary Figure 1: Individualized RF models exhibited a strong performance for identifying all four time periods.**

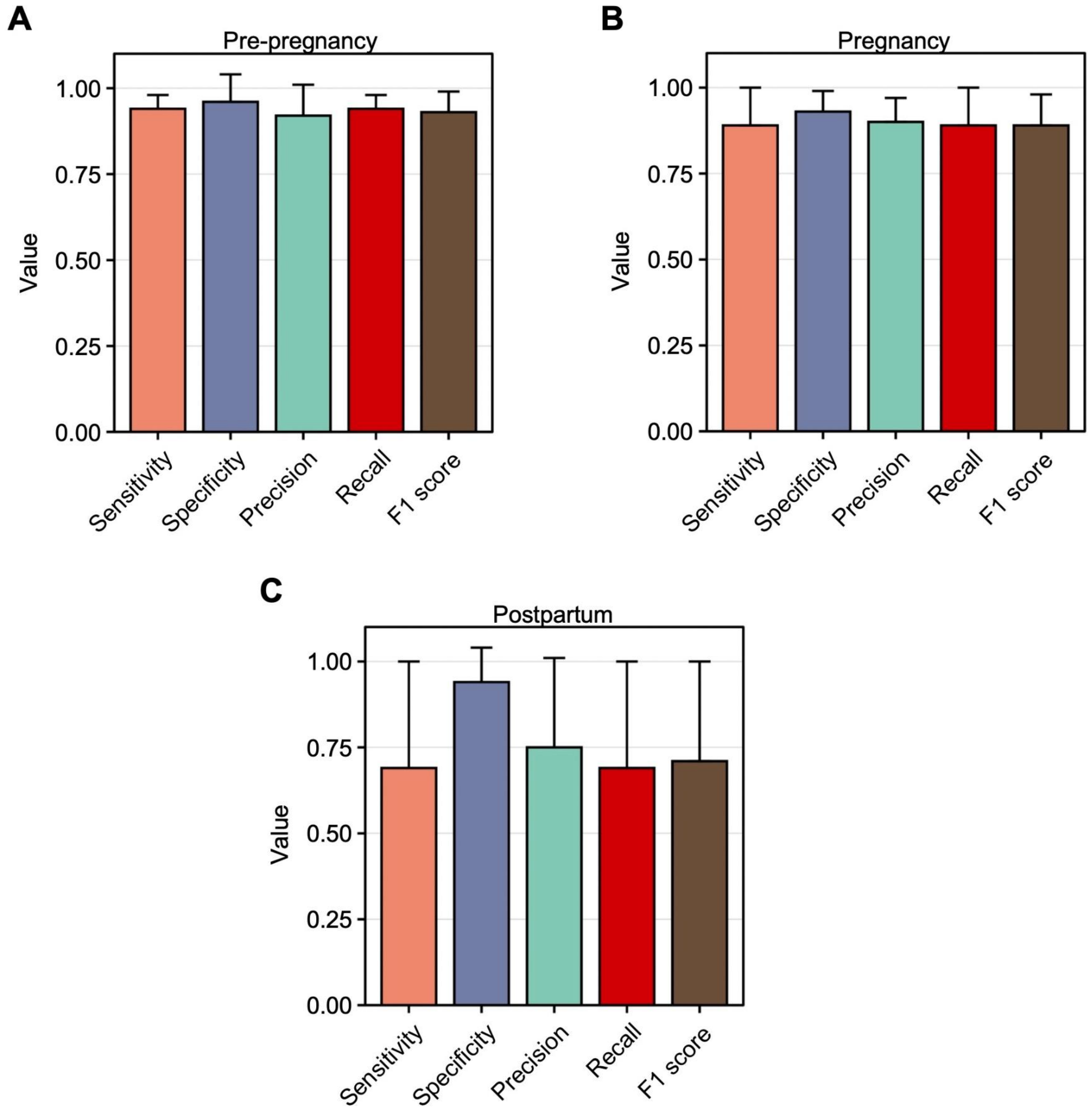


**A:** The sensitivity, specificity, precision, recall, and F1 score across individual RF models for the pre-pregnancy class.

**B:** The sensitivity, specificity, precision, recall, and F1 score across individual RF models for the pregnancy class.

**C:** The sensitivity, specificity, precision, recall, and F1 score across individual RF models for the postpartum class.

Individualized RF models exhibited robust performance in terms of sensitivity, specificity, precision, recall, and F1 score for recognizing the pre-pregnancy, pregnancy, and postpartum time periods.

Data are expressed as mean ± SD in A-C.

**Supplementary Figure 2: Prior history of depression before or duing pregnancy did not impact model performance for recognizing the PPD time period.**

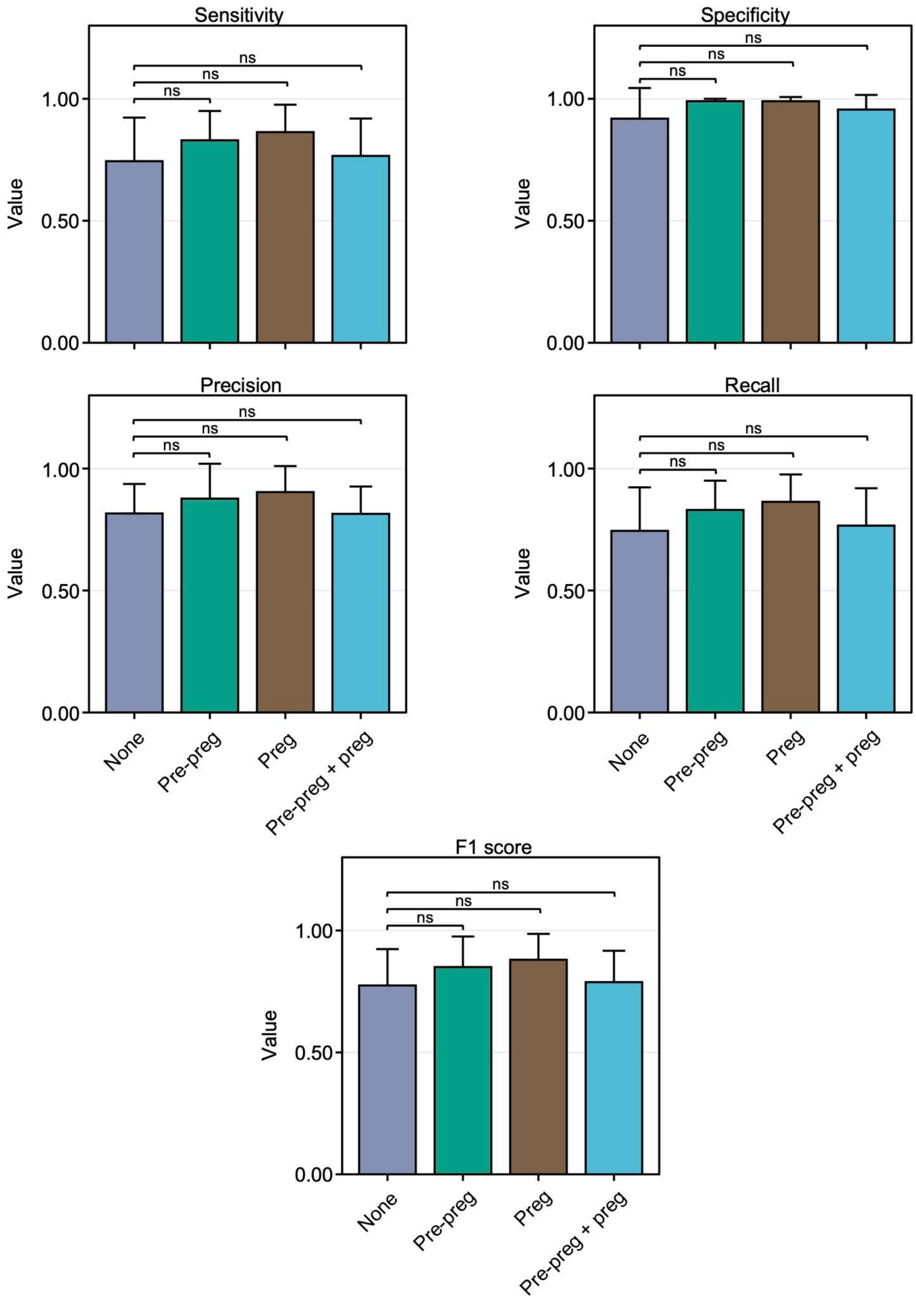

The sensitivity (top left), specificity (top right), precision (middle left), recall (middle right), and F1 score (bottom middle) of individualized ML models in women from the PPD cohort with 1) no prior history of depression (None), 2) history pre-pregnancy (Pre-preg), 3) history during pregnancy (Preg), or 4) history of both pre-pregnancy and during pregnancy (Pre-preg + preg). Among women in the PPD cohort, the average sensitivity, specificity, precision, recall, and F1 score displayed no variation across those with no history of depression, history of depression prior to pregnancy, history of depression during pregnancy, or history of depression both prior to and during pregnancy. Data are expressed as mean ± SD.

**Supplementary Figure 3: SHapley Additive exPlanations (SHAP) beeswarm plots were used for variable importance in individualized ML models.**

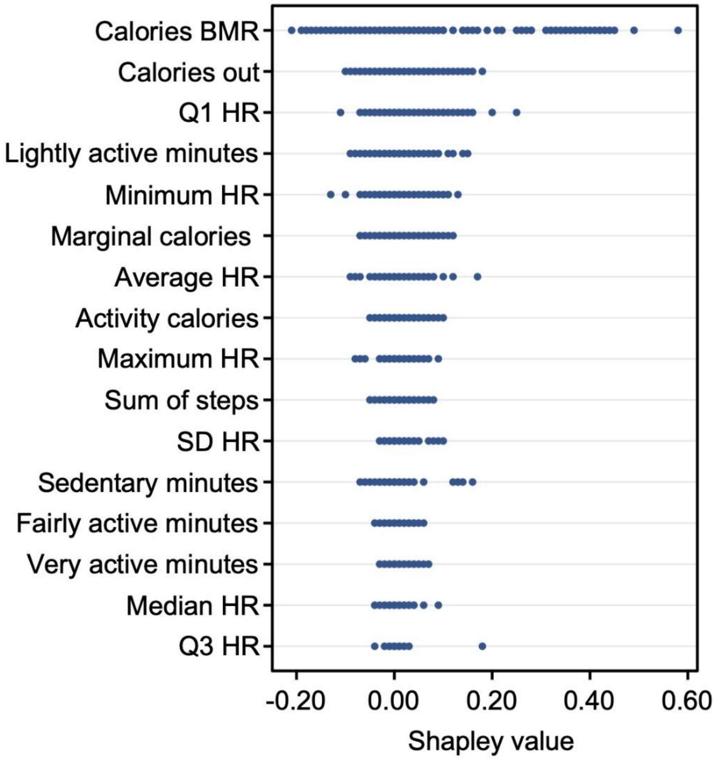


An example of a beeswarm SHapley Additive exPlanations (SHAP) plot for assessing variable importance in individualized ML models.

An example SHAP beeswarm plot showcasing the ranking of digital biomarkers according to their average Absolute Shapley values within an individualized model for predicting the PPD time period. In this example, the five most predictive features for the PPD time period were calories BMR, calories out, Q1 HR, lightly active minutes, and minimum HR.

**Supplementary Figure 4: An example of SHAP dependence plots assessing the relationship between digital biomarkers and PPD.**

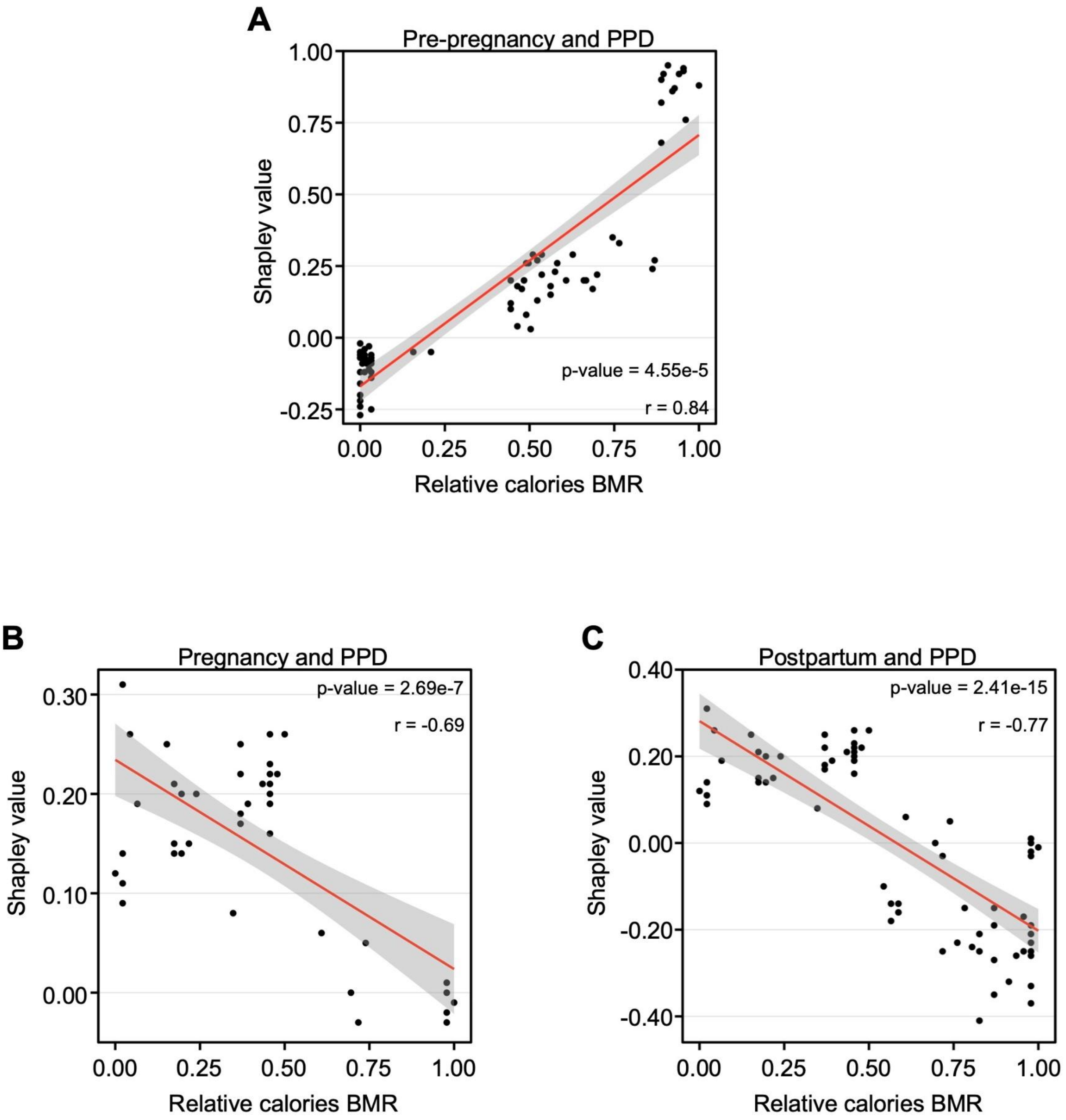


**A:** An example of a SHAP dependence plot during pre-pregnancy/PPD time periods.

**B:** An example of a SHAP dependence plot during pregnancy/PPD time periods.

**C:** An example of a SHAP dependence plot during postpartum/PPD time periods.

Example SHAP dependence dependence plots from individualized models demonstrated a correlation between Shapley values and actual values of calories BMR between the pre-pregnancy/PPD time periods in addition to a negative correlation during the pregnancy/PPD and postpartum/PPD time periods. This process was repeated for each individual to calculate the percentage of women with a significant correlation. Of those with a significant correlation, the percentage of women with a positive or negative correlation between Shapley values and actual values of the digital biomarker of interest was calculated.

**Supplementary Figure 5: The relationship between digital biomarkers provided insights into PPD classification in individualized N-of-1 ML models.**

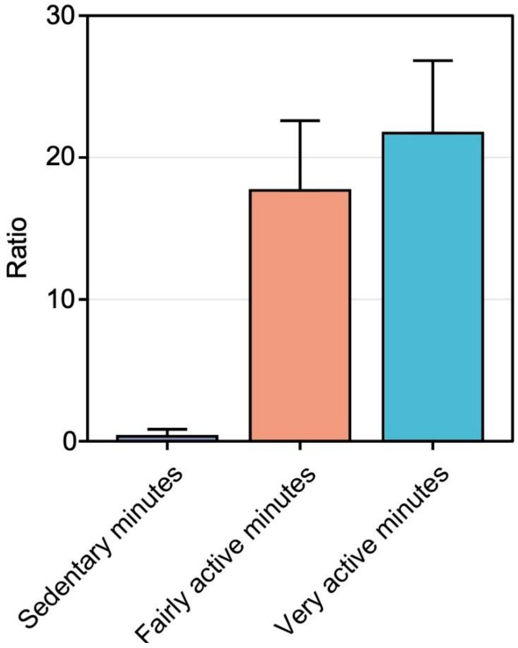


The ratio of lightly active minutes to sedentary minutes, fairly active minutes, and very active minutes across all individuals during the pregnancy and PPD time periods. The ratio of lightly active minutes to very active minutes surpassed that of lightly active minutes to fairly active minutes, which in turn is greater than the ratio of lightly active minutes to sedentary minutes. Data are expressed as mean ± SD.
