## Supplementary Table 7 for "Harnessing consumer wearable digital biomarkers for individualized recognition of postpartum depression using the *All of Us* Research Program dataset"

**Supplementary Table 7: OMOP concept IDs used in computational phenotyping of PPD and non-PPD cohorts.**

| Name | Concept Id | Variable | Variable class | Data source | Notes |
|---|---|---|---|---|---|
| Delivery by cesarean section for footling breech presentation | 46273629 | Cesarean section | Delivery | Conditions | |
| Delivery by cesarean section for flexed breech presentation | 46273305 | Cesarean section | Delivery | Conditions | |
| Delivery by cesarean section for breech presentation | 46273304 | Cesarean section | Delivery | Conditions | |
| Emergency lower segment cesarean section with inverted T incision | 46270991 | Cesarean section | Delivery | Conditions | |
| Other specified other caesarean delivery | 44513733 | Cesarean section | Delivery | Conditions | |
| Other specified elective caesarean delivery | 44513729 | Cesarean section | Delivery | Conditions | |
| Cesarean section through J shaped incision of uterus | 42872493 | Cesarean section | Delivery | Conditions | |
| Cesarean section through inverted T shaped incision of uterus | 42872492 | Cesarean section | Delivery | Conditions | |
| Triplet liveborn in hospital by cesarean section | 42539210 | Cesarean section | Delivery | Conditions | |
| Emergency lower segment cesarean section with bilateral tubal ligation | 42537021 | Cesarean section | Delivery | Conditions | |
| Elective lower segment cesarean section with bilateral tubal ligation | 42536960 | Cesarean section | Delivery | Conditions | |
| Emergency upper segment cesarean section with bilateral tubal ligation | 42536954 | Cesarean section | Delivery | Conditions | |
| Elective upper segment cesarean section with bilateral tubal ligation | 42536952 | Cesarean section | Delivery | Conditions | |
| Single liveborn born in hospital by cesarean section | 40483521 | Cesarean section | Delivery | Conditions | |
| Liveborn born in hospital by cesarean section | 40483126 | Cesarean section | Delivery | Conditions | |
| Twin liveborn born in hospital by cesarean section | 40483101 | Cesarean section | Delivery | Conditions | |
| Cesarean section w/o CC/MCC | 38001486 | Cesarean section | Delivery | Conditions | |
| Cesarean section w CC/MCC | 38001485 | Cesarean section | Delivery | Conditions | |
| Lower uterine segment cesarean section | 37312440 | Cesarean section | Delivery | Conditions | |
| Preterm delivery following Cesarean section | 37110284 | Cesarean section | Delivery | Conditions | |
| Born by emergency cesarean section | 4250010 | Cesarean section | Delivery | Conditions | |
| Vaginal cesarean section | 4228344 | Cesarean section | Delivery | Conditions | |
| Classical cesarean section | 4223536 | Cesarean section | Delivery | Conditions | |
| Born by elective cesarean section | 4212794 | Cesarean section | Delivery | Conditions | |
| Extraperitoneal cesarean section | 4211824 | Cesarean section | Delivery | Conditions | |
| Born by cesarean section | 4192676 | Cesarean section | Delivery | Conditions | |
| Placenta previa found before labor AND delivery by cesarean section without hemorrhage | 4172142 | Cesarean section | Delivery | Conditions | |
| Anesthesia for cesarean section | 4171820 | Cesarean section | Delivery | Conditions | |
| Emergency cesarean section | 4167089 | Cesarean section | Delivery | Conditions | |
| Abdominal delivery for shoulder dystocia | 4130321 | Cesarean section | Delivery | Conditions | |
| Emergency lower segment cesarean section | 4127252 | Cesarean section | Delivery | Conditions | |
| Delivered by cesarean delivery following previous cesarean delivery | 4119336 | Cesarean section | Delivery | Conditions | |
| Delivered by cesarean section - pregnancy at term | 4118802 | Cesarean section | Delivery | Conditions | |
| Elective cesarean section | 4075182 | Cesarean section | Delivery | Conditions | |
| Elective lower segment cesarean section | 4075161 | Cesarean section | Delivery | Conditions | |
| Elective upper segment cesarean section | 4075160 | Cesarean section | Delivery | Conditions | |
| Delivery by emergency cesarean section | 4066112 | Cesarean section | Delivery | Conditions | |
| Cesarean section - pregnancy at term | 4066111 | Cesarean section | Delivery | Conditions | |
| Cesarean section following previous cesarean section | 4065739 | Cesarean section | Delivery | Conditions | |
| Multiple delivery, all by cesarean section | 4063163 | Cesarean section | Delivery | Conditions | |
| Delivery by elective cesarean section | 4061457 | Cesarean section | Delivery | Conditions | |
| Emergency upper segment cesarean section | 4032767 | Cesarean section | Delivery | Conditions | |
| Cesarean section | 4015701 | Cesarean section | Delivery | Conditions | |
| Cesarean delivery only, following attempted vaginal delivery after previous cesarean delivery; including postpartum care | 2110324 | Cesarean section | Delivery | Conditions | |
| Cesarean delivery only, following attempted vaginal delivery after previous cesarean delivery | 2110323 | Cesarean section | Delivery | Conditions | |
| Cesarean delivery only; including postpartum care | 2110317 | Cesarean section | Delivery | Conditions | |
| Cesarean delivery only | 2110316 | Cesarean section | Delivery | Conditions | |
| Anesthesia for cesarean delivery following neuraxial labor analgesia/anesthesia (List separately in addition to code for primary procedure performed) | 2101814 | Cesarean section | Delivery | Conditions | |
| Anesthesia for cesarean delivery only | 2101807 | Cesarean section | Delivery | Conditions | |
| Neuraxial analgesia/anesthesia for labor ending in a cesarean delivery (includes any repeat subarachnoid needle placement and drug injection and/or any necessary replacement of an epidural catheter during labor) | 2101016 | Cesarean section | Delivery | Conditions | |
| Anesthesia for intraperitoneal procedures in lower abdomen including laparoscopy; cesarean section | 2101013 | Cesarean section | Delivery | Conditions | |
| Other cesarean section of unspecified type | 2004802 | Cesarean section | Delivery | Conditions | |
| Cesarean section of other specified type | 2004789 | Cesarean section | Delivery | Conditions | |
| Low cervical cesarean section | 2004786 | Cesarean section | Delivery | Conditions | |
| Cesarean delivery - delivered | 437942 | Cesarean section | Delivery | Conditions | |
| Deliveries by cesarean | 193277 | Cesarean section | Delivery | Conditions | |
| Delivery by cesarean section for footling breech presentation | 46273629 | Delivery record only (DELIV) | Delivery | Conditions | |
| Delivery by cesarean section for flexed breech presentation | 46273305 | Delivery record only (DELIV) | Delivery | Conditions | |
| Delivery by cesarean section for breech presentation | 46273304 | Delivery record only (DELIV) | Delivery | Conditions | |
| Vacuum assisted vaginal delivery | 45757174 | Delivery record only (DELIV) | Delivery | Conditions | |
| Other specified cephalic vaginal delivery with abnormal presentation of head at delivery without instrument | 44513756 | Delivery record only (DELIV) | Delivery | Conditions | |
| Other specified vacuum delivery | 44513752 | Delivery record only (DELIV) | Delivery | Conditions | |
| Other specified forceps cephalic delivery | 44513747 | Delivery record only (DELIV) | Delivery | Conditions | |

| | | | | |
|---|---|---|---|---|
| Low forceps cephalic delivery | 44513746 | Delivery record only (DELIV) | Delivery | Conditions |
| Mid forceps cephalic delivery NEC | 44513745 | Delivery record only (DELIV) | Delivery | Conditions |
| High forceps cephalic delivery NEC | 44513743 | Delivery record only (DELIV) | Delivery | Conditions |
| Other specified other breech delivery | 44513740 | Delivery record only (DELIV) | Delivery | Conditions |
| Other specified breech extraction delivery | 44513736 | Delivery record only (DELIV) | Delivery | Conditions |
| Multiple liveborn in hospital by vaginal delivery | 42539267 | Delivery record only (DELIV) | Delivery | Conditions |
| Mid vacuum delivery | 42535817 | Delivery record only (DELIV) | Delivery | Conditions |
| Outlet vacuum delivery | 42535816 | Delivery record only (DELIV) | Delivery | Conditions |
| Born by mid-cavity forceps delivery | 37310404 | Delivery record only (DELIV) | Delivery | Conditions |
| Born by low forceps delivery | 37310393 | Delivery record only (DELIV) | Delivery | Conditions |
| Born by high forceps delivery | 37310369 | Delivery record only (DELIV) | Delivery | Conditions |
| Single liveborn born in hospital by vaginal delivery | 36713074 | Delivery record only (DELIV) | Delivery | Conditions |
| Vaginal delivery, medical personnel present | 4331179 | Delivery record only (DELIV) | Delivery | Conditions |
| Forceps delivery with rotation of fetal head | 4324549 | Delivery record only (DELIV) | Delivery | Conditions |
| Pinard maneuver | 4294827 | Delivery record only (DELIV) | Delivery | Conditions |
| Trial forceps delivery | 4290245 | Delivery record only (DELIV) | Delivery | Conditions |
| Mid forceps delivery | 4266683 | Delivery record only (DELIV) | Delivery | Conditions |
| Kristeller maneuver | 4250441 | Delivery record only (DELIV) | Delivery | Conditions |
| Born by breech delivery | 4250009 | Delivery record only (DELIV) | Delivery | Conditions |
| Frank breech delivery | 4244672 | Delivery record only (DELIV) | Delivery | Conditions |
| Bracht maneuver | 4240325 | Delivery record only (DELIV) | Delivery | Conditions |
| Delivery by Malstrom's extraction | 4234710 | Delivery record only (DELIV) | Delivery | Conditions |
| Partial breech delivery | 4234421 | Delivery record only (DELIV) | Delivery | Conditions |
| Delivery by Kielland rotation | 4232028 | Delivery record only (DELIV) | Delivery | Conditions |
| High forceps delivery | 4231702 | Delivery record only (DELIV) | Delivery | Conditions |
| Partial breech extraction | 4230533 | Delivery record only (DELIV) | Delivery | Conditions |
| Delivery by Malstrom's extraction with episiotomy | 4223638 | Delivery record only (DELIV) | Delivery | Conditions |
| Footling breech delivery | 4217642 | Delivery record only (DELIV) | Delivery | Conditions |
| Born by forceps delivery | 4217586 | Delivery record only (DELIV) | Delivery | Conditions |
| Breech extraction | 4213387 | Delivery record only (DELIV) | Delivery | Conditions |
| Spontaneous vertex delivery | 4205240 | Delivery record only (DELIV) | Delivery | Conditions |
| Total breech delivery with forceps to aftercoming head | 4204679 | Delivery record only (DELIV) | Delivery | Conditions |
| Delivery by vacuum extraction | 4189205 | Delivery record only (DELIV) | Delivery | Conditions |
| Premature pregnancy delivered | 4175637 | Delivery record only (DELIV) | Delivery | Conditions |
| Neville-Barnes forceps delivery | 4173513 | Delivery record only (DELIV) | Delivery | Conditions |
| Abnormal delivery | 4170626 | Delivery record only (DELIV) | Delivery | Conditions |
| Simpson's forceps delivery | 4170152 | Delivery record only (DELIV) | Delivery | Conditions |
| Vaginal delivery with forceps including postpartum care | 4167018 | Delivery record only (DELIV) | Delivery | Conditions |
| Brow delivery | 4166775 | Delivery record only (DELIV) | Delivery | Conditions |
| Total breech extraction | 4166247 | Delivery record only (DELIV) | Delivery | Conditions |
| Face delivery | 4164221 | Delivery record only (DELIV) | Delivery | Conditions |
| Delivered by low forceps delivery | 4156948 | Delivery record only (DELIV) | Delivery | Conditions |
| Deliveries by spontaneous breech delivery | 4154615 | Delivery record only (DELIV) | Delivery | Conditions |
| Delivered by mid-cavity forceps delivery | 4153284 | Delivery record only (DELIV) | Delivery | Conditions |
| Barton's forceps delivery | 4147974 | Delivery record only (DELIV) | Delivery | Conditions |
| Midforceps delivery without rotation | 4143647 | Delivery record only (DELIV) | Delivery | Conditions |
| Breech presentation, no version | 4137400 | Delivery record only (DELIV) | Delivery | Conditions |
| Delivery of the after coming head | 4130319 | Delivery record only (DELIV) | Delivery | Conditions |
| Instrumental delivery | 4128031 | Delivery record only (DELIV) | Delivery | Conditions |
| Term pregnancy delivered | 4127705 | Delivery record only (DELIV) | Delivery | Conditions |
| Mauriceau Smellie Veit maneuver | 4127251 | Delivery record only (DELIV) | Delivery | Conditions |
| Lovset's maneuver | 4127250 | Delivery record only (DELIV) | Delivery | Conditions |
| Outlet forceps delivery | 4127249 | Delivery record only (DELIV) | Delivery | Conditions |
| Nonrotational forceps delivery | 4127248 | Delivery record only (DELIV) | Delivery | Conditions |
| Born after precipitate delivery | 4119050 | Delivery record only (DELIV) | Delivery | Conditions |
| Abnormal head presentation delivery | 4118904 | Delivery record only (DELIV) | Delivery | Conditions |
| Normal delivery - occipitoanterior | 4118903 | Delivery record only (DELIV) | Delivery | Conditions |
| Forceps delivery | 4114637 | Delivery record only (DELIV) | Delivery | Conditions |
| Breech extraction with internal podalic version | 4114636 | Delivery record only (DELIV) | Delivery | Conditions |
| Delivery by double application of forceps | 4106406 | Delivery record only (DELIV) | Delivery | Conditions |
| Prague maneuver | 4103850 | Delivery record only (DELIV) | Delivery | Conditions |
| Wigand-Martin maneuver | 4102595 | Delivery record only (DELIV) | Delivery | Conditions |
| Delivery by Scanzoni maneuver | 4100410 | Delivery record only (DELIV) | Delivery | Conditions |
| Delivery by vacuum extraction with episiotomy | 4096145 | Delivery record only (DELIV) | Delivery | Conditions |
| Mid forceps delivery with episiotomy | 4093774 | Delivery record only (DELIV) | Delivery | Conditions |
| Low forceps delivery | 4088084 | Delivery record only (DELIV) | Delivery | Conditions |
| Face to pubes birth | 4083524 | Delivery record only (DELIV) | Delivery | Conditions |
| Low forceps delivery with episiotomy | 4075735 | Delivery record only (DELIV) | Delivery | Conditions |
| Low vacuum delivery | 4075191 | Delivery record only (DELIV) | Delivery | Conditions |
| DeLee forceps cephalic delivery with rotation | 4075190 | Delivery record only (DELIV) | Delivery | Conditions |
| Midforceps cephalic delivery with rotation | 4075189 | Delivery record only (DELIV) | Delivery | Conditions |
| High forceps cephalic delivery with rotation | 4075188 | Delivery record only (DELIV) | Delivery | Conditions |

| | | | | |
|---|---|---|---|---|
| Forceps cephalic delivery | 4075187 | Delivery record only (DELIV) | Delivery | Conditions |
| Vacuum delivery before full dilation of cervix | 4075171 | Delivery record only (DELIV) | Delivery | Conditions |
| High vacuum delivery | 4075170 | Delivery record only (DELIV) | Delivery | Conditions |
| Barton forceps cephalic delivery with rotation | 4075169 | Delivery record only (DELIV) | Delivery | Conditions |
| Assisted breech delivery | 4075168 | Delivery record only (DELIV) | Delivery | Conditions |
| Breech extraction delivery with version | 4075165 | Delivery record only (DELIV) | Delivery | Conditions |
| Piper forceps delivery | 4073438 | Delivery record only (DELIV) | Delivery | Conditions |
| Trial of vacuum delivery | 4073424 | Delivery record only (DELIV) | Delivery | Conditions |
| Spontaneous breech delivery | 4073422 | Delivery record only (DELIV) | Delivery | Conditions |
| Non-manipulative cephalic vaginal delivery with abnormal presentation of head at delivery without instrument | 4071630 | Delivery record only (DELIV) | Delivery | Conditions |
| Deliveries by destructive operation | 4066113 | Delivery record only (DELIV) | Delivery | Conditions |
| Delivery by combination of forceps and vacuum extractor | 4065737 | Delivery record only (DELIV) | Delivery | Conditions |
| Piper forceps delivery by application to aftercoming head | 4064824 | Delivery record only (DELIV) | Delivery | Conditions |
| Multiple delivery, all by forceps and vacuum extractor | 4063162 | Delivery record only (DELIV) | Delivery | Conditions |
| Normal delivery but ante- or post- natal conditions present | 4063160 | Delivery record only (DELIV) | Delivery | Conditions |
| Delivered by mid-cavity forceps with rotation | 4062136 | Delivery record only (DELIV) | Delivery | Conditions |
| High forceps delivery with episiotomy | 4035778 | Delivery record only (DELIV) | Delivery | Conditions |
| Complete breech delivery | 4034145 | Delivery record only (DELIV) | Delivery | Conditions |
| Burns Marshall maneuver | 4032766 | Delivery record only (DELIV) | Delivery | Conditions |
| Groin traction at breech delivery | 4032764 | Delivery record only (DELIV) | Delivery | Conditions |
| Forceps application to aftercoming head | 4032762 | Delivery record only (DELIV) | Delivery | Conditions |
| Forceps delivery, face to pubes | 4032761 | Delivery record only (DELIV) | Delivery | Conditions |
| Partial breech delivery with forceps to aftercoming head | 4023797 | Delivery record only (DELIV) | Delivery | Conditions |
| Normal birth | 4014720 | Delivery record only (DELIV) | Delivery | Conditions |
| Postmature pregnancy delivered | 4011047 | Delivery record only (DELIV) | Delivery | Conditions |
| Threatened premature labor - delivered | 3186670 | Delivery record only (DELIV) | Delivery | Conditions |
| Vaginal delivery only, after previous cesarean delivery (with or without episiotomy and/or forceps); including postpartum care | 2110321 | Delivery record only (DELIV) | Delivery | Conditions |
| Routine obstetric care including antepartum care, vaginal delivery (with or without episiotomy, and/or forceps) and postpartum care, after previous cesarean delivery | 2110319 | Delivery record only (DELIV) | Delivery | Conditions |
| Vaginal delivery only (with or without episiotomy and/or forceps); including postpartum care | 2110309 | Delivery record only (DELIV) | Delivery | Conditions |
| Vaginal delivery only (with or without episiotomy and/or forceps) | 2110308 | Delivery record only (DELIV) | Delivery | Conditions |
| Routine obstetric care including antepartum care, vaginal delivery (with or without episiotomy, and/or forceps) and postpartum care | 2110307 | Delivery record only (DELIV) | Delivery | Conditions |
| Other specified instrumental delivery | 2004731 | Delivery record only (DELIV) | Delivery | Conditions |
| Other vacuum extraction | 2004730 | Delivery record only (DELIV) | Delivery | Conditions |
| Other total breech extraction | 2004726 | Delivery record only (DELIV) | Delivery | Conditions |
| Other partial breech extraction | 2004724 | Delivery record only (DELIV) | Delivery | Conditions |
| Other high forceps operation | 2004710 | Delivery record only (DELIV) | Delivery | Conditions |
| Other mid forceps operation | 2004707 | Delivery record only (DELIV) | Delivery | Conditions |
| Forceps, vacuum, and breech delivery | 2004702 | Delivery record only (DELIV) | Delivery | Conditions |
| Vacuum extractor delivery - delivered | 442069 | Delivery record only (DELIV) | Delivery | Conditions |
| Delivery normal | 441641 | Delivery record only (DELIV) | Delivery | Conditions |
| Elderly primigravida - delivered | 440794 | Delivery record only (DELIV) | Delivery | Conditions |
| Deliveries by breech extraction | 440793 | Delivery record only (DELIV) | Delivery | Conditions |
| Deliveries by vacuum extractor | 440790 | Delivery record only (DELIV) | Delivery | Conditions |
| Triplet pregnancy - delivered | 440462 | Delivery record only (DELIV) | Delivery | Conditions |
| Quadruplet pregnancy - delivered | 438481 | Delivery record only (DELIV) | Delivery | Conditions |
| Breech extraction - delivered | 436767 | Delivery record only (DELIV) | Delivery | Conditions |
| Forceps delivery - delivered | 435022 | Delivery record only (DELIV) | Delivery | Conditions |
| Twin pregnancy - delivered | 435018 | Delivery record only (DELIV) | Delivery | Conditions |
| Grand multiparity - delivered | 434110 | Delivery record only (DELIV) | Delivery | Conditions |
| Mother delivered | 433260 | Delivery record only (DELIV) | Delivery | Conditions |
| Breech presentation - delivered | 73537 | Delivery record only (DELIV) | Delivery | Conditions |
| Triplet liveborn in hospital by cesarean section | 42539210 | Delivery record only (DELIV) | Delivery | Conditions |
| Twin liveborn born in hospital by cesarean section | 40483101 | Delivery record only (DELIV) | Delivery | Conditions |
| Multiple delivery, all by cesarean section | 4063163 | Delivery record only (DELIV) | Delivery | Conditions |
| Delayed delivery second twin - delivered | 4064559 | Delivery record only (DELIV) | Delivery | Conditions |
| Locked twins - delivered | 442422 | Delivery record only (DELIV) | Delivery | Conditions |
| Multiple pregnancy with malpresentation - delivered | 442082 | Delivery record only (DELIV) | Delivery | Conditions |
| Emergency lower segment cesarean section with inverted T incision | 46270991 | Delivery record only (DELIV) | Delivery | Conditions |
| Other specified other caesarean delivery | 44513733 | Delivery record only (DELIV) | Delivery | Conditions |
| Other specified elective caesarean delivery | 44513729 | Delivery record only (DELIV) | Delivery | Conditions |
| Cesarean section through J shaped incision of uterus | 42872493 | Delivery record only (DELIV) | Delivery | Conditions |
| Cesarean section through inverted T shaped incision of uterus | 42872492 | Delivery record only (DELIV) | Delivery | Conditions |
| Emergency lower segment cesarean section with bilateral tubal ligation | 42537021 | Delivery record only (DELIV) | Delivery | Conditions |
| Elective lower segment cesarean section with bilateral tubal ligation | 42536960 | Delivery record only (DELIV) | Delivery | Conditions |
| Emergency upper segment cesarean section with bilateral tubal ligation | 42536954 | Delivery record only (DELIV) | Delivery | Conditions |
| Elective upper segment cesarean section with bilateral tubal ligation | 42536952 | Delivery record only (DELIV) | Delivery | Conditions |
| Single liveborn born in hospital by cesarean section | 40483521 | Delivery record only (DELIV) | Delivery | Conditions |
| Liveborn born in hospital by cesarean section | 40483126 | Delivery record only (DELIV) | Delivery | Conditions |

| | | | | |
|---|---|---|---|---|
| Cesarean section w/o CC/MCC | 38001486 | Delivery record only (DELIV) | Delivery | Conditions |
| Cesarean section w CC/MCC | 38001485 | Delivery record only (DELIV) | Delivery | Conditions |
| Lower uterine segment cesarean section | 37312440 | Delivery record only (DELIV) | Delivery | Conditions |
| Preterm delivery following Cesarean section | 37110284 | Delivery record only (DELIV) | Delivery | Conditions |
| Born by emergency cesarean section | 4250010 | Delivery record only (DELIV) | Delivery | Conditions |
| Vaginal cesarean section | 4228344 | Delivery record only (DELIV) | Delivery | Conditions |
| Classical cesarean section | 4223536 | Delivery record only (DELIV) | Delivery | Conditions |
| Born by elective cesarean section | 4212794 | Delivery record only (DELIV) | Delivery | Conditions |
| Extraperitoneal cesarean section | 4211824 | Delivery record only (DELIV) | Delivery | Conditions |
| Born by cesarean section | 4192676 | Delivery record only (DELIV) | Delivery | Conditions |
| Placenta previa found before labor AND delivery by cesarean section without hemorrhage | 4172142 | Delivery record only (DELIV) | Delivery | Conditions |
| Anesthesia for cesarean section | 4171820 | Delivery record only (DELIV) | Delivery | Conditions |
| Emergency cesarean section | 4167089 | Delivery record only (DELIV) | Delivery | Conditions |
| Abdominal delivery for shoulder dystocia | 4130321 | Delivery record only (DELIV) | Delivery | Conditions |
| Emergency lower segment cesarean section | 4127252 | Delivery record only (DELIV) | Delivery | Conditions |
| Delivered by cesarean delivery following previous cesarean delivery | 4119336 | Delivery record only (DELIV) | Delivery | Conditions |
| Delivered by cesarean section - pregnancy at term | 4118802 | Delivery record only (DELIV) | Delivery | Conditions |
| Elective cesarean section | 4075182 | Delivery record only (DELIV) | Delivery | Conditions |
| Elective lower segment cesarean section | 4075161 | Delivery record only (DELIV) | Delivery | Conditions |
| Elective upper segment cesarean section | 4075160 | Delivery record only (DELIV) | Delivery | Conditions |
| Delivery by emergency cesarean section | 4066112 | Delivery record only (DELIV) | Delivery | Conditions |
| Cesarean section - pregnancy at term | 4066111 | Delivery record only (DELIV) | Delivery | Conditions |
| Cesarean section following previous cesarean section | 4065739 | Delivery record only (DELIV) | Delivery | Conditions |
| Delivery by elective cesarean section | 4061457 | Delivery record only (DELIV) | Delivery | Conditions |
| Emergency upper segment cesarean section | 4032767 | Delivery record only (DELIV) | Delivery | Conditions |
| Cesarean section | 4015701 | Delivery record only (DELIV) | Delivery | Conditions |
| Cesarean delivery only, following attempted vaginal delivery after previous cesarean delivery; including postpartum care | 2110324 | Delivery record only (DELIV) | Delivery | Conditions |
| Cesarean delivery only, following attempted vaginal delivery after previous cesarean delivery | 2110323 | Delivery record only (DELIV) | Delivery | Conditions |
| Cesarean delivery only; including postpartum care | 2110317 | Delivery record only (DELIV) | Delivery | Conditions |
| Cesarean delivery only | 2110316 | Delivery record only (DELIV) | Delivery | Conditions |
| Anesthesia for cesarean delivery following neuraxial labor analgesia/anesthesia (List separately in addition to code for primary procedure performed) | 2101814 | Delivery record only (DELIV) | Delivery | Conditions |
| Anesthesia for cesarean delivery only | 2101807 | Delivery record only (DELIV) | Delivery | Conditions |
| Neuraxial analgesia/anesthesia for labor ending in a cesarean delivery (includes any repeat subarachnoid needle placement and drug injection and/or any necessary replacement of an epidural catheter during labor) | 2101016 | Delivery record only (DELIV) | Delivery | Conditions |
| Anesthesia for intraperitoneal procedures in lower abdomen including laparoscopy; cesarean section | 2101013 | Delivery record only (DELIV) | Delivery | Conditions |
| Other cesarean section of unspecified type | 2004802 | Delivery record only (DELIV) | Delivery | Conditions |
| Cesarean section of other specified type | 2004789 | Delivery record only (DELIV) | Delivery | Conditions |
| Low cervical cesarean section | 2004786 | Delivery record only (DELIV) | Delivery | Conditions |
| Cesarean delivery - delivered | 437942 | Delivery record only (DELIV) | Delivery | Conditions |
| Deliveries by cesarean | 193277 | Delivery record only (DELIV) | Delivery | Conditions |
| Cystocele - delivered with postpartum complication | 4129700 | Delivery record only (DELIV) | Delivery | Conditions |
| Hemorrhoids in pregnancy and the puerperium - delivered with postnatal complication | 4066133 | Delivery record only (DELIV) | Delivery | Conditions |
| Puerperal septicemia - delivered with postnatal complication | 4066124 | Delivery record only (DELIV) | Delivery | Conditions |
| Puerperal peritonitis - delivered with postnatal complication | 4066122 | Delivery record only (DELIV) | Delivery | Conditions |
| Puerperal endometritis - delivered with postnatal complication | 4065749 | Delivery record only (DELIV) | Delivery | Conditions |
| Obstetric anesthesia with central nervous system complications - delivered | 4065622 | Delivery record only (DELIV) | Delivery | Conditions |
| Obstetric anesthesia with cardiac complications - delivered | 4065621 | Delivery record only (DELIV) | Delivery | Conditions |
| Secondary postpartum hemorrhage - delivered with postnatal problem | 4065618 | Delivery record only (DELIV) | Delivery | Conditions |
| Rupture of uterus during and after labor - delivered with postnatal problem | 4064960 | Delivery record only (DELIV) | Delivery | Conditions |
| Hypertonic uterine inertia - delivered | 4064834 | Delivery record only (DELIV) | Delivery | Conditions |
| Persistent occipitoposterior or occipitoanterior position - delivered | 4064426 | Delivery record only (DELIV) | Delivery | Conditions |
| Fetus with drug damage - delivered | 4064177 | Delivery record only (DELIV) | Delivery | Conditions |
| Fetus with viral damage via mother - delivered | 4064174 | Delivery record only (DELIV) | Delivery | Conditions |
| Fetus with hereditary disease - delivered | 4064171 | Delivery record only (DELIV) | Delivery | Conditions |
| Fetal distress - delivered | 4063305 | Delivery record only (DELIV) | Delivery | Conditions |
| Fetus with damage due to intrauterine contraceptive device - delivered | 4063296 | Delivery record only (DELIV) | Delivery | Conditions |
| Transverse lie - delivered | 4063171 | Delivery record only (DELIV) | Delivery | Conditions |
| Glycosuria during pregnancy - delivered | 4063028 | Delivery record only (DELIV) | Delivery | Conditions |
| Fatigue during pregnancy - delivered with postnatal complication | 4062572 | Delivery record only (DELIV) | Delivery | Conditions |
| Fatigue during pregnancy - delivered | 4062571 | Delivery record only (DELIV) | Delivery | Conditions |
| Hemorrhoids in pregnancy and the puerperium - delivered | 4062268 | Delivery record only (DELIV) | Delivery | Conditions |
| Obstetric anesthesia with pulmonary complications - delivered with postnatal problem | 4062116 | Delivery record only (DELIV) | Delivery | Conditions |
| Obstetric anesthesia with pulmonary complications - delivered | 4062115 | Delivery record only (DELIV) | Delivery | Conditions |
| Puerperal salpingitis - delivered with postnatal complication | 4061465 | Delivery record only (DELIV) | Delivery | Conditions |
| Obstetric anesthesia with central nervous system complication - delivered with postnatal problem | 4061349 | Delivery record only (DELIV) | Delivery | Conditions |
| Obstetric anesthesia with cardiac complications - delivered with postnatal problem | 4061346 | Delivery record only (DELIV) | Delivery | Conditions |
| Prolonged artificial rupture of membranes - delivered | 4060695 | Delivery record only (DELIV) | Delivery | Conditions |

| | | | | |
|---|---|---|---|---|
| Fetus with radiation damage - delivered | 4060675 | Delivery record only (DELIV) | Delivery | Conditions |
| Uterine scar from previous surgery in pregnancy, childbirth and the puerperium - delivered | 4060558 | Delivery record only (DELIV) | Delivery | Conditions |
| Brow presentation - delivered | 4060540 | Delivery record only (DELIV) | Delivery | Conditions |
| Face presentation - delivered | 4060539 | Delivery record only (DELIV) | Delivery | Conditions |
| Herpes gestationis - delivered | 4060301 | Delivery record only (DELIV) | Delivery | Conditions |
| Umbilical cord tight around neck - delivered | 4060157 | Delivery record only (DELIV) | Delivery | Conditions |
| Oblique lie - delivered | 4059757 | Delivery record only (DELIV) | Delivery | Conditions |
| Eclampsia with postnatal complication | 4058536 | Delivery record only (DELIV) | Delivery | Conditions |
| Antepartum hemorrhage with uterine leiomyoma - delivered | 4058526 | Delivery record only (DELIV) | Delivery | Conditions |
| Antepartum hemorrhage with trauma - delivered | 4058524 | Delivery record only (DELIV) | Delivery | Conditions |
| Glycosuria during pregnancy - delivered with postnatal complication | 4058111 | Delivery record only (DELIV) | Delivery | Conditions |
| Herpes gestationis - delivered with postnatal complication | 4058110 | Delivery record only (DELIV) | Delivery | Conditions |
| Rectocele - delivered with postpartum complication | 4028632 | Delivery record only (DELIV) | Delivery | Conditions |
| Failed trial of labor - delivered | 443521 | Delivery record only (DELIV) | Delivery | Conditions |
| Papyraceous fetus - delivered | 443324 | Delivery record only (DELIV) | Delivery | Conditions |
| Obstetric non-purulent mastitis - delivered | 443294 | Delivery record only (DELIV) | Delivery | Conditions |
| Antepartum hemorrhage with coagulation defect - delivered | 443133 | Delivery record only (DELIV) | Delivery | Conditions |
| Pelvic soft tissue abnormality in pregnancy, childbirth and the puerperium - delivered | 442829 | Delivery record only (DELIV) | Delivery | Conditions |
| Fetus with central nervous system malformation - delivered | 442828 | Delivery record only (DELIV) | Delivery | Conditions |
| Obstetric pyemic and septic pulmonary embolism - delivered | 442420 | Delivery record only (DELIV) | Delivery | Conditions |
| Puerperal cerebrovascular disorder - delivered | 442419 | Delivery record only (DELIV) | Delivery | Conditions |
| Puerperal cerebrovascular disorder - delivered with postnatal complication | 442418 | Delivery record only (DELIV) | Delivery | Conditions |
| Placental polyp - delivered with postnatal complication | 442090 | Delivery record only (DELIV) | Delivery | Conditions |
| Obstetric nipple infection - delivered | 442089 | Delivery record only (DELIV) | Delivery | Conditions |
| Obstetric laceration of cervix - delivered | 442080 | Delivery record only (DELIV) | Delivery | Conditions |
| Obstetric shock - delivered | 442072 | Delivery record only (DELIV) | Delivery | Conditions |
| Maternal hypotension syndrome - delivered | 442070 | Delivery record only (DELIV) | Delivery | Conditions |
| Varicose veins of legs in pregnancy and the puerperium - delivered | 442059 | Delivery record only (DELIV) | Delivery | Conditions |
| Amniotic fluid pulmonary embolism - delivered | 442054 | Delivery record only (DELIV) | Delivery | Conditions |
| Obstetric blood-clot pulmonary embolism - delivered | 442053 | Delivery record only (DELIV) | Delivery | Conditions |
| Obstetric air pulmonary embolism - delivered with postnatal complication | 442052 | Delivery record only (DELIV) | Delivery | Conditions |
| Obstetric air pulmonary embolism - delivered | 442049 | Delivery record only (DELIV) | Delivery | Conditions |
| Obstetric pyemic and septic pulmonary embolism - delivered with postnatal complication | 442048 | Delivery record only (DELIV) | Delivery | Conditions |
| Failed mechanical induction - delivered | 441924 | Delivery record only (DELIV) | Delivery | Conditions |
| Antenatal deep vein thrombosis - delivered | 441649 | Delivery record only (DELIV) | Delivery | Conditions |
| Prolapse of cord - delivered | 441645 | Delivery record only (DELIV) | Delivery | Conditions |
| Maternal distress - delivered with postnatal problem | 441643 | Delivery record only (DELIV) | Delivery | Conditions |
| History of recurrent miscarriage - delivered | 441630 | Delivery record only (DELIV) | Delivery | Conditions |
| Cervical incompetence - delivered | 441369 | Delivery record only (DELIV) | Delivery | Conditions |
| Postpartum coagulation defects - delivered with postnatal problem | 441363 | Delivery record only (DELIV) | Delivery | Conditions |
| Cracked nipple in pregnancy, the puerperium or lactation - delivered with postnatal complication | 440481 | Delivery record only (DELIV) | Delivery | Conditions |
| Prolonged first stage - delivered | 440475 | Delivery record only (DELIV) | Delivery | Conditions |
| Post-term pregnancy - delivered | 439894 | Delivery record only (DELIV) | Delivery | Conditions |
| Superficial thrombophlebitis in pregnancy and the puerperium - delivered with postnatal complication | 439095 | Delivery record only (DELIV) | Delivery | Conditions |
| Superficial thrombophlebitis in pregnancy and the puerperium - delivered | 439094 | Delivery record only (DELIV) | Delivery | Conditions |
| Cord tangled with compression - delivered | 439092 | Delivery record only (DELIV) | Delivery | Conditions |
| Severe pre-eclampsia - delivered with postnatal complication | 439077 | Delivery record only (DELIV) | Delivery | Conditions |
| Severe pre-eclampsia - delivered | 438490 | Delivery record only (DELIV) | Delivery | Conditions |
| Precipitate labor - delivered | 438220 | Delivery record only (DELIV) | Delivery | Conditions |
| Amniotic cavity infection - delivered | 437936 | Delivery record only (DELIV) | Delivery | Conditions |
| Fetal intrauterine distress first noted during labor AND/OR delivery in liveborn infant | 437098 | Delivery record only (DELIV) | Delivery | Conditions |
| Obstetric nipple infection - delivered with postnatal complication | 437055 | Delivery record only (DELIV) | Delivery | Conditions |
| Varicose veins of perineum and vulva in pregnancy and the puerperium - delivered with postnatal complication | 436766 | Delivery record only (DELIV) | Delivery | Conditions |
| Maternal distress - delivered | 436183 | Delivery record only (DELIV) | Delivery | Conditions |
| Hyperemesis gravidarum with metabolic disturbance - delivered | 436173 | Delivery record only (DELIV) | Delivery | Conditions |
| Retracted nipple in pregnancy, the puerperium or lactation - delivered | 435612 | Delivery record only (DELIV) | Delivery | Conditions |
| Varicose veins of perineum and vulva in pregnancy and the puerperium - delivered | 435610 | Delivery record only (DELIV) | Delivery | Conditions |
| Vasa previa - delivered | 435609 | Delivery record only (DELIV) | Delivery | Conditions |
| Cervical incompetence - delivered with postnatal complication | 435607 | Delivery record only (DELIV) | Delivery | Conditions |
| Varicose veins of legs in pregnancy and the puerperium - delivered with postnatal complication | 435330 | Delivery record only (DELIV) | Delivery | Conditions |
| Postnatal deep vein thrombosis - delivered with postnatal complication | 435031 | Delivery record only (DELIV) | Delivery | Conditions |
| Short cord - delivered | 435020 | Delivery record only (DELIV) | Delivery | Conditions |
| Cracked nipple in pregnancy, the puerperium or lactation - delivered | 434715 | Delivery record only (DELIV) | Delivery | Conditions |
| Puerperal pyrexia of unknown origin - delivered with postnatal complication | 434713 | Delivery record only (DELIV) | Delivery | Conditions |
| Prolonged second stage - delivered | 434431 | Delivery record only (DELIV) | Delivery | Conditions |
| Polyhydramnios - delivered | 434427 | Delivery record only (DELIV) | Delivery | Conditions |
| Obstetric blood-clot pulmonary embolism - delivered with postnatal complication | 434112 | Delivery record only (DELIV) | Delivery | Conditions |
| Tuberculosis in pregnancy, childbirth and the puerperium - delivered | 433812 | Delivery record only (DELIV) | Delivery | Conditions |

| | | | | |
|---|---|---|---|---|
| Retracted nipple in pregnancy, the puerperium or lactation - delivered with postnatal complication | 433276 | Delivery record only (DELIV) | Delivery | Conditions |
| Oligohydramnios - delivered | 433274 | Delivery record only (DELIV) | Delivery | Conditions |
| Amniotic fluid pulmonary embolism - delivered with postnatal complication | 432388 | Delivery record only (DELIV) | Delivery | Conditions |
| Vascular lesions of cord - delivered | 321367 | Delivery record only (DELIV) | Delivery | Conditions |
| Benign essential hypertension complicating pregnancy, childbirth and the puerperium - delivered with postnatal complication | 320456 | Delivery record only (DELIV) | Delivery | Conditions |
| Benign essential hypertension complicating pregnancy, childbirth and the puerperium - delivered | 314103 | Delivery record only (DELIV) | Delivery | Conditions |
| Maternal hypotension syndrome - delivered with postnatal problem | 313829 | Delivery record only (DELIV) | Delivery | Conditions |
| Renal hypertension complicating pregnancy, childbirth and the puerperium - delivered with postnatal complication | 201912 | Delivery record only (DELIV) | Delivery | Conditions |
| Obstetric pelvic hematoma - delivered | 201368 | Delivery record only (DELIV) | Delivery | Conditions |
| Placenta previa without hemorrhage - delivered | 201359 | Delivery record only (DELIV) | Delivery | Conditions |
| Premature rupture of membranes - delivered | 200160 | Delivery record only (DELIV) | Delivery | Conditions |
| Renal hypertension complicating pregnancy, childbirth and the puerperium - delivered | 200157 | Delivery record only (DELIV) | Delivery | Conditions |
| Obstructed labor caused by bony pelvis - delivered | 199087 | Delivery record only (DELIV) | Delivery | Conditions |
| Rupture of uterus before labor - delivered | 198816 | Delivery record only (DELIV) | Delivery | Conditions |
| Second degree perineal tear during delivery - delivered | 198499 | Delivery record only (DELIV) | Delivery | Conditions |
| Disproportion - major pelvic abnormality - delivered | 198486 | Delivery record only (DELIV) | Delivery | Conditions |
| Retroverted incarcerated gravid uterus - delivered | 198216 | Delivery record only (DELIV) | Delivery | Conditions |
| Retroverted incarcerated gravid uterus - delivered with postnatal complication | 197616 | Delivery record only (DELIV) | Delivery | Conditions |
| Obstetric inversion of uterus - delivered with postnatal problem | 197048 | Delivery record only (DELIV) | Delivery | Conditions |
| Peripheral neuritis in pregnancy - delivered | 197042 | Delivery record only (DELIV) | Delivery | Conditions |
| Primary uterine inertia - delivered | 196762 | Delivery record only (DELIV) | Delivery | Conditions |
| Obstructed labor caused by pelvic soft tissues - delivered | 196182 | Delivery record only (DELIV) | Delivery | Conditions |
| Liver disorder in pregnancy - delivered | 196170 | Delivery record only (DELIV) | Delivery | Conditions |
| Rhesus isoimmunization - delivered | 195878 | Delivery record only (DELIV) | Delivery | Conditions |
| Obstetric shock - delivered with postnatal problem | 195025 | Delivery record only (DELIV) | Delivery | Conditions |
| Obstructed labor due to fetal malposition - delivered | 194711 | Delivery record only (DELIV) | Delivery | Conditions |
| Fourth degree perineal tear during delivery - delivered | 194429 | Delivery record only (DELIV) | Delivery | Conditions |
| Cesarean wound disruption - delivered with postnatal complication | 194113 | Delivery record only (DELIV) | Delivery | Conditions |
| Placental abruption - delivered | 193539 | Delivery record only (DELIV) | Delivery | Conditions |
| Fetal-maternal hemorrhage - delivered | 193535 | Delivery record only (DELIV) | Delivery | Conditions |
| Third degree perineal tear during delivery - delivered | 193275 | Delivery record only (DELIV) | Delivery | Conditions |
| Obstetric pelvic hematoma - delivered with postnatal problem | 193271 | Delivery record only (DELIV) | Delivery | Conditions |
| Obstetric high vaginal laceration - delivered | 193269 | Delivery record only (DELIV) | Delivery | Conditions |
| Placenta previa with hemorrhage - delivered | 193264 | Delivery record only (DELIV) | Delivery | Conditions |
| First degree perineal tear during delivery - delivered | 192978 | Delivery record only (DELIV) | Delivery | Conditions |
| Pelvic soft tissue abnormality in pregnancy, childbirth and the puerperium - delivered with postnatal complication | 192971 | Delivery record only (DELIV) | Delivery | Conditions |
| Rupture of uterus during and after labor - delivered | 192699 | Delivery record only (DELIV) | Delivery | Conditions |
| Obstetric perineal wound disruption - delivered with postnatal complication | 192387 | Delivery record only (DELIV) | Delivery | Conditions |
| Secondary uterine inertia - delivered | 192384 | Delivery record only (DELIV) | Delivery | Conditions |
| Transient hypertension of pregnancy - delivered with postnatal complication | 141639 | Delivery record only (DELIV) | Delivery | Conditions |
| Eclampsia - delivered with postnatal complication | 138811 | Delivery record only (DELIV) | Delivery | Conditions |
| Transient hypertension of pregnancy - delivered | 137940 | Delivery record only (DELIV) | Delivery | Conditions |
| Pre-eclampsia or eclampsia with pre-existing hypertension - delivered | 135601 | Delivery record only (DELIV) | Delivery | Conditions |
| Pre-eclampsia or eclampsia with pre-existing hypertension - delivered with postnatal complication | 134414 | Delivery record only (DELIV) | Delivery | Conditions |
| Eclampsia - delivered | 133816 | Delivery record only (DELIV) | Delivery | Conditions |
| Genitourinary tract infection in pregnancy - delivered | 81357 | Delivery record only (DELIV) | Delivery | Conditions |
| Breast engorgement in pregnancy, the puerperium or lactation - delivered with postnatal complication | 81086 | Delivery record only (DELIV) | Delivery | Conditions |
| Large fetus causing disproportion - delivered | 80782 | Delivery record only (DELIV) | Delivery | Conditions |
| Genitourinary tract infection in pregnancy - delivered with postnatal complication | 80778 | Delivery record only (DELIV) | Delivery | Conditions |
| Failure of lactation - delivered with postnatal complication | 80479 | Delivery record only (DELIV) | Delivery | Conditions |
| Shoulder dystocia - delivered | 80474 | Delivery record only (DELIV) | Delivery | Conditions |
| Obstetric non-purulent mastitis - delivered with postnatal complication | 79897 | Delivery record only (DELIV) | Delivery | Conditions |
| Unstable lie - delivered | 79889 | Delivery record only (DELIV) | Delivery | Conditions |
| Obstetric breast abscess - delivered with postnatal complication | 78494 | Delivery record only (DELIV) | Delivery | Conditions |
| Breast engorgement in pregnancy, the puerperium or lactation - delivered | 77624 | Delivery record only (DELIV) | Delivery | Conditions |
| Inlet pelvic contraction - delivered | 77621 | Delivery record only (DELIV) | Delivery | Conditions |
| High head at term - delivered | 77615 | Delivery record only (DELIV) | Delivery | Conditions |
| Suppressed lactation - delivered | 77347 | Delivery record only (DELIV) | Delivery | Conditions |
| Failure of lactation - delivered | 77056 | Delivery record only (DELIV) | Delivery | Conditions |
| Generally contracted pelvis - delivered | 77052 | Delivery record only (DELIV) | Delivery | Conditions |
| Suppressed lactation - delivered with postnatal complication | 76769 | Delivery record only (DELIV) | Delivery | Conditions |
| Asymptomatic bacteriuria in pregnancy - delivered | 76761 | Delivery record only (DELIV) | Delivery | Conditions |
| Deep transverse arrest - delivered | 75882 | Delivery record only (DELIV) | Delivery | Conditions |
| Obstetric damage to pelvic joints and ligaments - delivered | 75326 | Delivery record only (DELIV) | Delivery | Conditions |
| Galactorrhea in pregnancy and the puerperium - delivered with postnatal complication | 74717 | Delivery record only (DELIV) | Delivery | Conditions |
| Galactorrhea in pregnancy and the puerperium - delivered | 74440 | Delivery record only (DELIV) | Delivery | Conditions |

| | | | | |
|---|---|---|---|---|
| Obstetric breast abscess - delivered | 74433 | Delivery record only (DELIV) | Delivery | Conditions |
| Outlet pelvic contraction - delivered | 73824 | Delivery record only (DELIV) | Delivery | Conditions |
| Hydrocephalic disproportion - delivered | 73538 | Delivery record only (DELIV) | Delivery | Conditions |
| Asymptomatic bacteriuria in pregnancy - delivered with postnatal complication | 73526 | Delivery record only (DELIV) | Delivery | Conditions |
| Mixed feto-pelvic disproportion - delivered | 72970 | Delivery record only (DELIV) | Delivery | Conditions |
| Fetus with chromosomal abnormality - delivered | 72697 | Delivery record only (DELIV) | Delivery | Conditions |
| Prolapsed arm - delivered | 72692 | Delivery record only (DELIV) | Delivery | Conditions |
| Delivery of Products of Conception, External Approach | 2784578 | Delivery record only (DELIV) | Delivery | Conditions |
| Manual Extraction of Products of Conception, Retained, Via Natural or Artificial Opening Endoscopic | 43018345 | Delivery record only (DELIV) | Delivery | Conditions |
| Manual Extraction of Products of Conception, Retained, Via Natural or Artificial Opening | 43018344 | Delivery record only (DELIV) | Delivery | Conditions |
| Extraction of Products of Conception, Retained, Via Natural or Artificial Opening Endoscopic | 2784574 | Delivery record only (DELIV) | Delivery | Conditions |
| Extraction of Products of Conception, Retained, Via Natural or Artificial Opening | 2784573 | Delivery record only (DELIV) | Delivery | Conditions |
| Extraction of Products of Conception, Other, Via Natural or Artificial Opening | 2784572 | Delivery record only (DELIV) | Delivery | Conditions |
| Extraction of Products of Conception, Internal Version, Via Natural or Artificial Opening | 2784571 | Delivery record only (DELIV) | Delivery | Conditions |
| Extraction of Products of Conception, Vacuum, Via Natural or Artificial Opening | 2784570 | Delivery record only (DELIV) | Delivery | Conditions |
| Extraction of Products of Conception, High Forceps, Via Natural or Artificial Opening | 2784569 | Delivery record only (DELIV) | Delivery | Conditions |
| Extraction of Products of Conception, Mid Forceps, Via Natural or Artificial Opening | 2784568 | Delivery record only (DELIV) | Delivery | Conditions |
| Extraction of Products of Conception, Low Forceps, Via Natural or Artificial Opening | 2784567 | Delivery record only (DELIV) | Delivery | Conditions |
| Extraction of Products of Conception, Extraperitoneal, Open Approach | 2784566 | Delivery record only (DELIV) | Delivery | Conditions |
| Extraction of Products of Conception, Low, Open Approach | 2784565 | Delivery record only (DELIV) | Delivery | Conditions |
| Extraction of Products of Conception, High, Open Approach | 2784564 | Delivery record only (DELIV) | Delivery | Conditions |
| Premature 28 week quadruplet | 3174212 | Livebirth (LB) | Delivery | Conditions |
| Premature birth of newborn sextuplets | 4008884 | Livebirth (LB) | Delivery | Conditions |
| Single live birth | 4014295 | Livebirth (LB) | Delivery | Conditions |
| Twins - both live born | 4014296 | Livebirth (LB) | Delivery | Conditions |
| Twins - one still and one live born | 4014455 | Livebirth (LB) | Delivery | Conditions |
| Triplets - all live born | 4014456 | Livebirth (LB) | Delivery | Conditions |
| Triplets - two live and one stillborn | 4015421 | Livebirth (LB) | Delivery | Conditions |
| Triplets - one live and two stillborn | 4015422 | Livebirth (LB) | Delivery | Conditions |
| Premature birth of newborn twins | 4029786 | Livebirth (LB) | Delivery | Conditions |
| Term birth of identical twins, both living | 4049333 | Livebirth (LB) | Delivery | Conditions |
| Term birth of newborn male | 4052512 | Livebirth (LB) | Delivery | Conditions |
| Term birth of newborn | 4054968 | Livebirth (LB) | Delivery | Conditions |
| Term birth of multiple newborns | 4066292 | Livebirth (LB) | Delivery | Conditions |
| Premature birth of multiple newborns | 4069200 | Livebirth (LB) | Delivery | Conditions |
| Premature birth of newborn quintuplets | 4082863 | Livebirth (LB) | Delivery | Conditions |
| Livebirth | 4092289 | Livebirth (LB) | Delivery | Conditions |
| Triplet birth | 4094046 | Livebirth (LB) | Delivery | Conditions |
| Premature birth of fraternal twins, both living | 4097427 | Livebirth (LB) | Delivery | Conditions |
| Twin birth | 4101844 | Livebirth (LB) | Delivery | Conditions |
| Premature birth of newborn triplets | 4107401 | Livebirth (LB) | Delivery | Conditions |
| Premature birth of identical twins, both living | 4142021 | Livebirth (LB) | Delivery | Conditions |
| Premature birth of newborn quadruplets | 4145125 | Livebirth (LB) | Delivery | Conditions |
| Premature birth of fraternal twins, one living, one stillborn | 4147043 | Livebirth (LB) | Delivery | Conditions |
| Multiple birth | 4163851 | Livebirth (LB) | Delivery | Conditions |
| Premature birth of newborn male | 4172135 | Livebirth (LB) | Delivery | Conditions |
| Term birth of newborn twins | 4178275 | Livebirth (LB) | Delivery | Conditions |
| Term birth of newborn quadruplets | 4193224 | Livebirth (LB) | Delivery | Conditions |
| Term birth of newborn sextuplets | 4199146 | Livebirth (LB) | Delivery | Conditions |
| Term birth of fraternal twins, both living | 4227728 | Livebirth (LB) | Delivery | Conditions |
| Premature birth of newborn female | 4241228 | Livebirth (LB) | Delivery | Conditions |
| Term birth of identical twins, one living, one stillborn | 4243026 | Livebirth (LB) | Delivery | Conditions |
| Term birth of newborn female | 4246654 | Livebirth (LB) | Delivery | Conditions |
| Premature birth of newborn | 4272248 | Livebirth (LB) | Delivery | Conditions |
| Term birth of newborn triplets | 4307237 | Livebirth (LB) | Delivery | Conditions |
| Premature birth of identical twins, one living, one stillborn | 4310910 | Livebirth (LB) | Delivery | Conditions |
| Term birth of fraternal twins, one living, one stillborn | 4330570 | Livebirth (LB) | Delivery | Conditions |
| Term birth of newborn quintuplets | 4336090 | Livebirth (LB) | Delivery | Conditions |
| Twin live born in hospital by vaginal delivery | 42535052 | Livebirth (LB) | Delivery | Conditions |
| Triplets, some live born | 45757165 | Livebirth (LB) | Delivery | Conditions |
| Quadruplets, all live born | 45757166 | Livebirth (LB) | Delivery | Conditions |
| Quintuplets, all live born | 45757167 | Livebirth (LB) | Delivery | Conditions |
| Quintuplets, some live born | 45757168 | Livebirth (LB) | Delivery | Conditions |
| Sextuplets, some live born | 45757169 | Livebirth (LB) | Delivery | Conditions |
| Quadruplet birth | 45765500 | Livebirth (LB) | Delivery | Conditions |
| Quintuplet birth | 45765501 | Livebirth (LB) | Delivery | Conditions |
| Sextuplet birth | 45765502 | Livebirth (LB) | Delivery | Conditions |
| Sextuplets, all live born | 45772082 | Livebirth (LB) | Delivery | Conditions |
| Quadruplets, some live born | 45773428 | Livebirth (LB) | Delivery | Conditions |
| Multiple liveborn in hospital by vaginal delivery | 42539267 | Livebirth (LB) | Delivery | Conditions |

| | | | | |
|---|---|---|---|---|
| Triplet liveborn in hospital by cesarean section | 42539210 | Livebirth (LB) | Delivery | Conditions |
| Single liveborn born in hospital by cesarean section | 40483521 | Livebirth (LB) | Delivery | Conditions |
| Liveborn born in hospital by cesarean section | 40483126 | Livebirth (LB) | Delivery | Conditions |
| Twin liveborn born in hospital by cesarean section | 40483101 | Livebirth (LB) | Delivery | Conditions |
| Twin liveborn born in hospital | 40483084 | Livebirth (LB) | Delivery | Conditions |
| Liveborn born in hospital | 40482735 | Livebirth (LB) | Delivery | Conditions |
| Twin liveborn born outside hospital | 36717637 | Livebirth (LB) | Delivery | Conditions |
| Multiple liveborn other than twins born outside hospital | 36713469 | Livebirth (LB) | Delivery | Conditions |
| Multiple liveborn other than twins born in hospital | 36713468 | Livebirth (LB) | Delivery | Conditions |
| Singleton liveborn unspecified as to place of birth | 36713467 | Livebirth (LB) | Delivery | Conditions |
| Singleton liveborn born outside hospital | 36713466 | Livebirth (LB) | Delivery | Conditions |
| Singleton liveborn born in hospital | 36713465 | Livebirth (LB) | Delivery | Conditions |
| Single liveborn born in hospital by vaginal delivery | 36713074 | Livebirth (LB) | Delivery | Conditions |
| Are you currently pregnant? | 1585811 | Pregnancy | Pregnancy | Survey |
| Pregnant | 4299535 | Pregnancy | Pregnancy | Conditions |
| High risk pregnancy | 4188598 | Pregnancy | Pregnancy | Conditions |
| High risk pregnancy due to history of preterm labor | 40480278 | Pregnancy | Pregnancy | Conditions |
| Intrauterine pregnancy | 197031 | Pregnancy | Pregnancy | Conditions |
| Unplanned pregnancy | 4307820 | Pregnancy | Pregnancy | Conditions |
| Normal pregnancy in primigravida | 36684864 | Pregnancy | Pregnancy | Conditions |
| Combined tubal and intrauterine pregnancy | 40483581 | Pregnancy | Pregnancy | Conditions |
| Pregnant - on history | 4061686 | Pregnancy | Pregnancy | Conditions |
| Surrogate pregnancy | 4012559 | Pregnancy | Pregnancy | Conditions |
| Pregnancy unplanned but wanted | 4060237 | Pregnancy | Pregnancy | Conditions |
| High risk pregnancy due to recurrent pregnancy loss | 37018878 | Pregnancy | Pregnancy | Conditions |
| Normal pregnancy | 4217975 | Pregnancy | Pregnancy | Conditions |
| Unwanted pregnancy | 4239301 | Pregnancy | Pregnancy | Conditions |
| Pregnant - urine test confirms | 4059981 | Pregnancy | Pregnancy | Conditions |
| 24 HR paroxetine hydrochloride 37.5 MG Extended Release Oral Tablet [Paxil] | 35604762 | Antidepressant | Depression | Drugs |
| venlafaxine 100 MG Oral Tablet [Effexor] | 19039817 | Antidepressant | Depression | Drugs |
| imipramine | 778268 | Antidepressant | Depression | Drugs |
| vilazodone | 40234834 | Antidepressant | Depression | Drugs |
| doxepin hydrochloride 150 MG Oral Capsule | 40224891 | Antidepressant | Depression | Drugs |
| nortriptyline 50 MG Oral Capsule | 19019413 | Antidepressant | Depression | Drugs |
| clomipramine hydrochloride 50 MG Oral Capsule | 40163070 | Antidepressant | Depression | Drugs |
| fluoxetine 10 MG Oral Tablet | 19079985 | Antidepressant | Depression | Drugs |
| escitalopram 20 MG Oral Tablet [Lexapro] | 715966 | Antidepressant | Depression | Drugs |
| amitriptyline hydrochloride 100 MG Oral Tablet | 40162683 | Antidepressant | Depression | Drugs |
| citalopram 20 MG Oral Tablet [Citalopraam] | 19113503 | Antidepressant | Depression | Drugs |
| trazodone hydrochloride 50 MG Oral Tablet [Desyrel] | 40163474 | Antidepressant | Depression | Drugs |
| mirtazapine 45 MG Disintegrating Oral Tablet | 725177 | Antidepressant | Depression | Drugs |
| mirtazapine 15 MG Disintegrating Oral Tablet | 19070747 | Antidepressant | Depression | Drugs |
| doxepin | 738156 | Antidepressant | Depression | Drugs |
| escitalopram 5 MG Oral Tablet | 715962 | Antidepressant | Depression | Drugs |
| venlafaxine 100 MG Oral Tablet | 19079835 | Antidepressant | Depression | Drugs |
| escitalopram 10 MG Oral Tablet [Lexapro] | 715965 | Antidepressant | Depression | Drugs |
| 12 HR bupropion hydrochloride 200 MG Extended Release Oral Tablet | 40221864 | Antidepressant | Depression | Drugs |
| milnacipran | 19080226 | Antidepressant | Depression | Drugs |
| 24 HR venlafaxine 150 MG Extended Release Oral Capsule [Effexor] | 743794 | Antidepressant | Depression | Drugs |
| desipramine hydrochloride 100 MG Oral Tablet | 40237422 | Antidepressant | Depression | Drugs |
| citalopram 40 MG Oral Tablet [Cipramil] | 19054100 | Antidepressant | Depression | Drugs |
| venlafaxine 37.5 MG Oral Tablet [Effexor] | 19039819 | Antidepressant | Depression | Drugs |
| doxepin hydrochloride 10 MG Oral Capsule | 40224875 | Antidepressant | Depression | Drugs |
| amitriptyline hydrochloride 10 MG Oral Tablet [Elavil] | 40162673 | Antidepressant | Depression | Drugs |
| 24 HR venlafaxine 225 MG Extended Release Oral Tablet | 19132561 | Antidepressant | Depression | Drugs |
| venlafaxine 75 MG Oral Tablet | 743752 | Antidepressant | Depression | Drugs |
| 24 HR desvenlafaxine succinate 100 MG Extended Release Oral Tablet [Pristiq] | 19129666 | Antidepressant | Depression | Drugs |
| Smoking Cessation 12 HR bupropion hydrochloride 150 MG Extended Release Oral Tablet [Zyban] | 40221863 | Antidepressant | Depression | Drugs |
| 24 HR venlafaxine 75 MG Extended Release Oral Capsule [Effexor] | 743793 | Antidepressant | Depression | Drugs |
| fluoxetine 40 MG Oral Capsule [Prozac] | 755735 | Antidepressant | Depression | Drugs |
| duloxetine 20 MG Delayed Release Oral Capsule [Cymbalta] | 715295 | Antidepressant | Depression | Drugs |
| 24 HR bupropion hydrochloride 300 MG Extended Release Oral Tablet [Wellbutrin] | 40221876 | Antidepressant | Depression | Drugs |
| duloxetine 30 MG Delayed Release Oral Capsule [Cymbalta] | 715298 | Antidepressant | Depression | Drugs |
| vortioxetine 20 MG Oral Tablet | 44506612 | Antidepressant | Depression | Drugs |
| fluvoxamine maleate 100 MG Oral Tablet | 40174725 | Antidepressant | Depression | Drugs |
| nortriptyline 75 MG Oral Capsule | 19019414 | Antidepressant | Depression | Drugs |
| venlafaxine 37.5 MG Oral Tablet | 743720 | Antidepressant | Depression | Drugs |
| doxepin hydrochloride 25 MG Oral Capsule | 40224897 | Antidepressant | Depression | Drugs |
| fluoxetine 20 MG Oral Capsule [Selfemra] | 19130466 | Antidepressant | Depression | Drugs |
| desipramine hydrochloride 50 MG Oral Tablet | 40237441 | Antidepressant | Depression | Drugs |
| bupropion hydrochloride 75 MG Oral Tablet | 40222092 | Antidepressant | Depression | Drugs |

| | | | | |
|---|---|---|---|---|
| trazodone hydrochloride 300 MG Oral Tablet | 40163468 | Antidepressant | Depression | Drugs |
| sertraline 25 MG Oral Tablet [Zoloft] | 739202 | Antidepressant | Depression | Drugs |
| 24 HR paroxetine hydrochloride 12.5 MG Extended Release Oral Tablet | 35604757 | Antidepressant | Depression | Drugs |
| citalopram 10 MG Oral Tablet [Celexa] | 797633 | Antidepressant | Depression | Drugs |
| 24 HR venlafaxine 150 MG Extended Release Oral Tablet | 19132560 | Antidepressant | Depression | Drugs |
| 24 HR bupropion hydrochloride 150 MG Extended Release Oral Tablet [Wellbutrin] | 40221873 | Antidepressant | Depression | Drugs |
| bupropion hydrochloride 100 MG Oral Tablet | 40222060 | Antidepressant | Depression | Drugs |
| mirtazapine 45 MG Oral Tablet | 725179 | Antidepressant | Depression | Drugs |
| paroxetine hydrochloride 30 MG Oral Tablet | 35604581 | Antidepressant | Depression | Drugs |
| fluvoxamine maleate 25 MG Oral Tablet | 40174735 | Antidepressant | Depression | Drugs |
| milnacipran hydrochloride 50 MG Oral Tablet [Savella] | 19080256 | Antidepressant | Depression | Drugs |
| 24 HR desvenlafaxine succinate 100 MG Extended Release Oral Tablet | 1593107 | Antidepressant | Depression | Drugs |
| fluoxetine 4 MG/ML Oral Solution | 19077464 | Antidepressant | Depression | Drugs |
| doxepin hydrochloride 10 MG Oral Capsule [Sinequan] | 40224877 | Antidepressant | Depression | Drugs |
| trazodone hydrochloride 100 MG Oral Tablet [Desyrel] | 40163461 | Antidepressant | Depression | Drugs |
| fluvoxamine maleate 100 MG Oral Tablet [Luvox] | 40174726 | Antidepressant | Depression | Drugs |
| citalopram 20 MG Oral Tablet [Celexa] | 797619 | Antidepressant | Depression | Drugs |
| fluoxetine 20 MG Oral Tablet | 755700 | Antidepressant | Depression | Drugs |
| venlafaxine 25 MG Oral Tablet | 743718 | Antidepressant | Depression | Drugs |
| alanine 20.7 MG/ML / arginine 11.5 MG/ML / glycine 10.3 MG/ML / histidine 4.8 MG/ML / isoleucine 6 MG/ML / leucine 7.3 MG/ML / lysine 5.8 MG/ML / methionine 4 MG/ML / phenylalanine 5.6 MG/ML / proline 6.8 MG/ML / serine 5 MG/ML / threonine 4.2 MG/ML /... | 19131343 | Antidepressant | Depression | Drugs |
| vortioxetine 10 MG Oral Tablet [Trintellix] | 42629569 | Antidepressant | Depression | Drugs |
| trazodone hydrochloride 150 MG Oral Tablet [Desyrel] | 40163465 | Antidepressant | Depression | Drugs |
| vilazodone hydrochloride 40 MG Oral Tablet [Viibryd] | 40234847 | Antidepressant | Depression | Drugs |
| vortioxetine 20 MG Oral Tablet [Trintellix] | 42629573 | Antidepressant | Depression | Drugs |
| doxepin hydrochloride 100 MG Oral Capsule | 40224885 | Antidepressant | Depression | Drugs |
| 12 HR bupropion hydrochloride 100 MG Extended Release Oral Tablet [Wellbutrin] | 40221858 | Antidepressant | Depression | Drugs |
| fluvoxamine | 751412 | Antidepressant | Depression | Drugs |
| vilazodone hydrochloride 20 MG Oral Tablet [Viibryd] | 40234843 | Antidepressant | Depression | Drugs |
| amitriptyline hydrochloride 25 MG Oral Tablet [Endep] | 40162719 | Antidepressant | Depression | Drugs |
| doxepin hydrochloride 50 MG Oral Capsule | 40224903 | Antidepressant | Depression | Drugs |
| desipramine hydrochloride 10 MG Oral Tablet | 40237418 | Antidepressant | Depression | Drugs |
| imipramine hydrochloride 25 MG Oral Tablet | 778296 | Antidepressant | Depression | Drugs |
| tranylcypromine 10 MG Oral Tablet [Parnate] | 19004820 | Antidepressant | Depression | Drugs |
| 24 HR venlafaxine 37.5 MG Extended Release Oral Capsule [Effexor] | 743795 | Antidepressant | Depression | Drugs |
| venlafaxine 50 MG Oral Tablet | 743753 | Antidepressant | Depression | Drugs |
| {7 (vilazodone hydrochloride 10 MG Oral Tablet [Viibryd]) / 7 (vilazodone hydrochloride 20 MG Oral Tablet [Viibryd]) / 16 (vilazodone hydrochloride 40 MG Oral Tablet [Viibryd]) } Pack [Viibryd 10/20/40 30 Day Pack] | 40235110 | Antidepressant | Depression | Drugs |
| escitalopram 5 MG Oral Tablet [Lexapro] | 19102734 | Antidepressant | Depression | Drugs |
| phenelzine 15 MG Oral Tablet | 733898 | Antidepressant | Depression | Drugs |
| 12 HR bupropion hydrochloride 150 MG Extended Release Oral Tablet [Budeprion] | 40221860 | Antidepressant | Depression | Drugs |
| nortriptyline 10 MG Oral Capsule [Aventyl] | 40239274 | Antidepressant | Depression | Drugs |
| amitriptyline hydrochloride 75 MG Oral Tablet | 40162737 | Antidepressant | Depression | Drugs |
| clomipramine hydrochloride 25 MG Oral Capsule | 40163063 | Antidepressant | Depression | Drugs |
| trimipramine 100 MG Oral Capsule | 705794 | Antidepressant | Depression | Drugs |
| mirtazapine 7.5 MG Oral Tablet | 19112586 | Antidepressant | Depression | Drugs |
| doxepin hydrochloride 75 MG Oral Capsule | 40224915 | Antidepressant | Depression | Drugs |
| mirtazapine 15 MG Oral Tablet [Remeron] | 725172 | Antidepressant | Depression | Drugs |
| protriptyline hydrochloride 10 MG Oral Tablet | 40175376 | Antidepressant | Depression | Drugs |
| paroxetine hydrochloride 10 MG Oral Tablet | 35604569 | Antidepressant | Depression | Drugs |
| paroxetine mesylate 40 MG Oral Tablet [Pexeva] | 722153 | Antidepressant | Depression | Drugs |
| citalopram 40 MG Oral Tablet [Celexa] | 797620 | Antidepressant | Depression | Drugs |
| mirtazapine 30 MG Oral Tablet [Remeron] | 725173 | Antidepressant | Depression | Drugs |
| vilazodone hydrochloride 10 MG Oral Tablet [Viibryd] | 40234839 | Antidepressant | Depression | Drugs |
| 12 HR bupropion hydrochloride 200 MG Extended Release Oral Tablet [Wellbutrin] | 40221866 | Antidepressant | Depression | Drugs |
| nortriptyline 50 MG Oral Capsule [Pamelor] | 19041194 | Antidepressant | Depression | Drugs |
| desvenlafaxine | 717607 | Antidepressant | Depression | Drugs |
| amitriptyline hydrochloride 50 MG Oral Tablet [Vanatrip] | 40162732 | Antidepressant | Depression | Drugs |
| clomipramine | 798834 | Antidepressant | Depression | Drugs |
| doxepin hydrochloride 25 MG Oral Capsule [Sinequan] | 40224899 | Antidepressant | Depression | Drugs |
| nefazodone hydrochloride 100 MG Oral Tablet | 40238415 | Antidepressant | Depression | Drugs |
| fluvoxamine maleate 50 MG Oral Tablet | 40174739 | Antidepressant | Depression | Drugs |
| desipramine | 716968 | Antidepressant | Depression | Drugs |
| nefazodone | 714684 | Antidepressant | Depression | Drugs |
| 24 HR paroxetine hydrochloride 25 MG Extended Release Oral Tablet | 35604759 | Antidepressant | Depression | Drugs |
| fluoxetine 10 MG Oral Capsule [Prozac] | 19029479 | Antidepressant | Depression | Drugs |
| 12 HR bupropion hydrochloride 90 MG / naltrexone hydrochloride 8 MG Extended Release Oral Tablet [Contrave] | 45774490 | Antidepressant | Depression | Drugs |
| duloxetine 40 MG Delayed Release Oral Capsule [Irenka] | 46233914 | Antidepressant | Depression | Drugs |
| nefazodone hydrochloride 150 MG Oral Tablet | 40238419 | Antidepressant | Depression | Drugs |

| | | | | |
|---|---|---|---|---|
| paroxetine hydrochloride 25 MG Extended Release Oral Tablet [Paxil] | 722159 | Antidepressant | Depression | Drugs |
| Smoking Cessation 12 HR bupropion hydrochloride 150 MG Extended Release Oral Tablet | 36249641 | Antidepressant | Depression | Drugs |
| paroxetine mesylate 30 MG Oral Tablet [Pexeva] | 722152 | Antidepressant | Depression | Drugs |
| 24 HR paroxetine hydrochloride 12.5 MG Extended Release Oral Tablet [Paxil] | 35604758 | Antidepressant | Depression | Drugs |
| paroxetine mesylate 20 MG Oral Tablet [Pexeva] | 722121 | Antidepressant | Depression | Drugs |
| desipramine hydrochloride 25 MG Oral Tablet | 40237432 | Antidepressant | Depression | Drugs |
| imipramine hydrochloride 50 MG Oral Tablet | 778299 | Antidepressant | Depression | Drugs |
| 24 HR venlafaxine 75 MG Extended Release Oral Tablet | 19132563 | Antidepressant | Depression | Drugs |
| 24 HR desvenlafaxine succinate 50 MG Extended Release Oral Tablet [Pristiq] | 19129683 | Antidepressant | Depression | Drugs |
| fluoxetine 60 MG Oral Tablet | 40243919 | Antidepressant | Depression | Drugs |
| nefazodone hydrochloride 200 MG Oral Tablet | 40238423 | Antidepressant | Depression | Drugs |
| paroxetine hydrochloride 20 MG Oral Tablet [Paxil] | 19035312 | Antidepressant | Depression | Drugs |
| fluvoxamine maleate 50 MG Oral Tablet [Luvox] | 40174740 | Antidepressant | Depression | Drugs |
| protriptyline hydrochloride 5 MG Oral Tablet | 40175380 | Antidepressant | Depression | Drugs |
| fluoxetine 20 MG Oral Capsule [Prozac] | 19004895 | Antidepressant | Depression | Drugs |
| vortioxetine 10 MG Oral Tablet | 44506611 | Antidepressant | Depression | Drugs |
| tranylcypromine 10 MG Oral Tablet | 703544 | Antidepressant | Depression | Drugs |
| nortriptyline 25 MG Oral Capsule [Aventyl] | 40239277 | Antidepressant | Depression | Drugs |
| sertraline 20 MG/ML Oral Solution | 40165282 | Antidepressant | Depression | Drugs |
| fluoxetine 10 MG Oral Capsule [Selfemra] | 19130449 | Antidepressant | Depression | Drugs |
| 12 HR bupropion hydrochloride 150 MG Extended Release Oral Tablet [Wellbutrin] | 40221862 | Antidepressant | Depression | Drugs |
| phenelzine | 733896 | Antidepressant | Depression | Drugs |
| desipramine hydrochloride 75 MG Oral Tablet | 40237445 | Antidepressant | Depression | Drugs |
| 24 HR venlafaxine 37.5 MG Extended Release Oral Tablet | 19132562 | Antidepressant | Depression | Drugs |
| PMDD fluoxetine 20 MG Oral Tablet [Sarafem] | 755854 | Antidepressant | Depression | Drugs |
| 24 HR desvenlafaxine succinate 50 MG Extended Release Oral Tablet | 1593112 | Antidepressant | Depression | Drugs |
| 24 HR desvenlafaxine 100 MG Extended Release Oral Tablet [Khedezla] | 43559999 | Antidepressant | Depression | Drugs |
| escitalopram 1 MG/ML Oral Solution | 715964 | Antidepressant | Depression | Drugs |
| mirtazapine 15 MG Disintegrating Oral Tablet [Remeron] | 725206 | Antidepressant | Depression | Drugs |
| amitriptyline hydrochloride 150 MG Oral Tablet | 40162695 | Antidepressant | Depression | Drugs |
| alanine 12.8 MG/ML / arginine 9.8 MG/ML / glycine 12.8 MG/ML / histidine 3 MG/ML / isoleucine 7.2 MG/ML / leucine 9.4 MG/ML / lysine 7.2 MG/ML / methionine 4 MG/ML / phenylalanine 4.4 MG/ML / proline 8.6 MG/ML / serine 4.2 MG/ML / threonine 5.2 MG/ML /... | 19133483 | Antidepressant | Depression | Drugs |
| bupropion hydrochloride 75 MG Oral Tablet [Wellbutrin] | 40222093 | Antidepressant | Depression | Drugs |
| doxepin hydrochloride 50 MG/ML Topical Cream | 40224909 | Antidepressant | Depression | Drugs |
| amoxapine 25 MG Oral Tablet | 713132 | Antidepressant | Depression | Drugs |
| 24 HR desvenlafaxine succinate 25 MG Extended Release Oral Tablet | 46221214 | Antidepressant | Depression | Drugs |
| bupropion hydrochloride 100 MG Oral Tablet [Wellbutrin] | 40222061 | Antidepressant | Depression | Drugs |
| doxepin hydrochloride 75 MG Oral Capsule [Sinequan] | 40224917 | Antidepressant | Depression | Drugs |
| venlafaxine 75 MG Oral Tablet [Effexor] | 19039822 | Antidepressant | Depression | Drugs |
| paroxetine hydrochloride 10 MG Oral Tablet [Paxil] | 722064 | Antidepressant | Depression | Drugs |
| alanine 10.4 MG/ML / arginine 5.75 MG/ML / calcium chloride 0.004 MEQ/ML / dibasic potassium phosphate 2.61 MG/ML / glucose 150 MG/ML / glycine 5.15 MG/ML / histidine 2.4 MG/ML / isoleucine 3 MG/ML / leucine 3.65 MG/ML / lysine 2.9 MG/ML / magnesium ch... | 19131082 | Antidepressant | Depression | Drugs |
| maprotiline hydrochloride 25 MG Oral Tablet | 42800984 | Antidepressant | Depression | Drugs |
| protriptyline hydrochloride 5 MG Oral Tablet [Vivactil] | 40175381 | Antidepressant | Depression | Drugs |
| imipramine hydrochloride 25 MG Oral Tablet [Tofranil] | 778297 | Antidepressant | Depression | Drugs |
| mirtazapine 45 MG Oral Tablet [Remeron] | 725174 | Antidepressant | Depression | Drugs |
| 24 HR desvenlafaxine 50 MG Extended Release Oral Tablet [Khedezla] | 43560000 | Antidepressant | Depression | Drugs |
| imipramine hydrochloride 10 MG Oral Tablet [Tofranil] | 778359 | Antidepressant | Depression | Drugs |
| imipramine hydrochloride 10 MG Oral Tablet | 778357 | Antidepressant | Depression | Drugs |
| doxepin 3 MG Oral Tablet [Silenor] | 40173385 | Antidepressant | Depression | Drugs |
| imipramine hydrochloride 50 MG Oral Tablet [Tofranil] | 19134040 | Antidepressant | Depression | Drugs |
| doxepin hydrochloride 50 MG Oral Capsule [Sinequan] | 40224905 | Antidepressant | Depression | Drugs |
| milnacipran hydrochloride 100 MG Oral Tablet [Savella] | 19080251 | Antidepressant | Depression | Drugs |
| 24 HR bupropion hydrochloride 150 MG Extended Release Oral Tablet [Budeprion] | 40221872 | Antidepressant | Depression | Drugs |
| 24 HR desvenlafaxine succinate 25 MG Extended Release Oral Tablet [Pristiq] | 46221216 | Antidepressant | Depression | Drugs |
| paroxetine mesylate 10 MG Oral Tablet [Pexeva] | 19113754 | Antidepressant | Depression | Drugs |
| 24 HR fluvoxamine maleate 150 MG Extended Release Oral Capsule | 40173466 | Antidepressant | Depression | Drugs |
| amitriptyline hydrochloride 75 MG Oral Tablet [Endep] | 40162739 | Antidepressant | Depression | Drugs |
| 24 HR desvenlafaxine 100 MG Extended Release Oral Tablet | 19129664 | Antidepressant | Depression | Drugs |
| amitriptyline hydrochloride 100 MG Oral Tablet [Endep] | 40162685 | Antidepressant | Depression | Drugs |
| citalopram 2 MG/ML Oral Solution | 19075393 | Antidepressant | Depression | Drugs |
| doxepin hydrochloride 150 MG Oral Capsule [Sinequan] | 40224893 | Antidepressant | Depression | Drugs |
| vortioxetine 5 MG Oral Tablet [Trintellix] | 42629575 | Antidepressant | Depression | Drugs |
| duloxetine 40 MG Delayed Release Oral Capsule | 19123437 | Antidepressant | Depression | Drugs |
| 24 HR bupropion hydrochloride 450 MG Extended Release Oral Tablet [Forfivo] | 42707260 | Antidepressant | Depression | Drugs |
| alanine 8.8 MG/ML / arginine 4.89 MG/ML / calcium chloride 0.004 MEQ/ML / dibasic potassium phosphate 2.61 MG/ML / glucose 50 MG/ML / glycine 4.38 MG/ML / histidine 2.04 MG/ML / isoleucine 2.55 MG/ML / leucine 3.11 MG/ML / lysine 2.47 MG/ML / magnesium... | 19131065 | Antidepressant | Depression | Drugs |

| | | | | |
|---|---|---|---|---|
| bupropion hydrochloride 150 MG Extended Release Oral Tablet | 40222065 | Antidepressant | Depression | Drugs |
| PMDD fluoxetine 10 MG Oral Tablet [Sarafem] | 755853 | Antidepressant | Depression | Drugs |
| paroxetine hydrochloride 12.5 MG Extended Release Oral Tablet [Paxil] | 722158 | Antidepressant | Depression | Drugs |
| amitriptyline hydrochloride 25 MG / perphenazine 2 MG Oral Tablet | 40162704 | Antidepressant | Depression | Drugs |
| doxepin hydrochloride 10 MG/ML Oral Solution | 40224881 | Antidepressant | Depression | Drugs |
| doxepin 3 MG Oral Tablet | 40173384 | Antidepressant | Depression | Drugs |
| paroxetine hydrochloride 37.5 MG Extended Release Oral Tablet [Paxil] | 722160 | Antidepressant | Depression | Drugs |
| paroxetine hydrochloride 30 MG Oral Tablet [Paxil] | 19035313 | Antidepressant | Depression | Drugs |
| bupropion hydrochloride 300 MG Extended Release Oral Tablet | 40222084 | Antidepressant | Depression | Drugs |
| amitriptyline hydrochloride 50 MG Oral Tablet [Domical] | 19025666 | Antidepressant | Depression | Drugs |
| 24 HR desvenlafaxine 50 MG Extended Release Oral Tablet | 19129681 | Antidepressant | Depression | Drugs |
| 12 HR bupropion hydrochloride 90 MG / naltrexone hydrochloride 8 MG Extended Release Oral Tablet | 45774486 | Antidepressant | Depression | Drugs |
| amitriptyline hydrochloride 10 MG / perphenazine 2 MG Oral Tablet | 40162658 | Antidepressant | Depression | Drugs |
| nortriptyline 75 MG Oral Capsule [Pamelor] | 19041328 | Antidepressant | Depression | Drugs |
| amitriptyline hydrochloride 25 MG Oral Tablet [Elavil] | 40162718 | Antidepressant | Depression | Drugs |
| alanine 8.8 MG/ML / arginine 4.89 MG/ML / calcium chloride 0.004 MEQ/ML / dibasic potassium phosphate 2.61 MG/ML / glucose 100 MG/ML / glycine 4.38 MG/ML / histidine 2.04 MG/ML / isoleucine 2.55 MG/ML / leucine 3.11 MG/ML / lysine 2.47 MG/ML / magnesiu... | 40235428 | Antidepressant | Depression | Drugs |
| {3 (0.2 ML) (esketamine 140 MG/ML Nasal Spray [Spravato]) } Pack [Spravato 84 MG Dose Kit] | 1366735 | Antidepressant | Depression | Drugs |
| alanine 10.4 MG/ML / arginine 5.75 MG/ML / glucose 200 MG/ML / glycine 5.15 MG/ML / histidine 2.4 MG/ML / isoleucine 3 MG/ML / leucine 3.65 MG/ML / lysine 2.9 MG/ML / methionine 2 MG/ML / phenylalanine 2.8 MG/ML / proline 3.4 MG/ML / serine 2.5 MG/ML /... | 19130815 | Antidepressant | Depression | Drugs |
| venlafaxine 25 MG Oral Tablet [Effexor] | 19039818 | Antidepressant | Depression | Drugs |
| 24 HR paroxetine hydrochloride 37.5 MG Extended Release Oral Tablet | 35604761 | Antidepressant | Depression | Drugs |
| doxepin 6 MG Oral Tablet | 40173388 | Antidepressant | Depression | Drugs |
| bupropion hydrochloride 100 MG Extended Release Oral Tablet | 40222057 | Antidepressant | Depression | Drugs |
| amitriptyline hydrochloride 50 MG Oral Tablet [Endep] | 40162731 | Antidepressant | Depression | Drugs |
| milnacipran hydrochloride 25 MG Oral Tablet [Savella] | 19080258 | Antidepressant | Depression | Drugs |
| duloxetine hydrochloride 60 MG Extended Release Oral Capsule | 715292 | Antidepressant | Depression | Drugs |
| mirtazapine 30 MG Disintegrating Oral Tablet | 19070748 | Antidepressant | Depression | Drugs |
| paroxetine mesylate 7.5 MG Oral Capsule [Brisdelle] | 43532895 | Antidepressant | Depression | Drugs |
| amoxapine | 713109 | Antidepressant | Depression | Drugs |
| 24 HR bupropion hydrochloride 450 MG Extended Release Oral Tablet | 42707259 | Antidepressant | Depression | Drugs |
| 12 HR bupropion hydrochloride 100 MG Extended Release Oral Tablet [Budeprion] | 40221857 | Antidepressant | Depression | Drugs |
| nefazodone hydrochloride 150 MG Oral Tablet [Serzone] | 40238420 | Antidepressant | Depression | Drugs |
| paroxetine hydrochloride 40 MG Oral Tablet [Paxil] | 19046308 | Antidepressant | Depression | Drugs |
| paroxetine mesylate 7.5 MG Oral Capsule | 43532894 | Antidepressant | Depression | Drugs |
| vortioxetine | 44507700 | Antidepressant | Depression | Drugs |
| imipramine pamoate 75 MG Oral Capsule | 778322 | Antidepressant | Depression | Drugs |
| nortriptyline 25 MG Oral Capsule [Pamelor] | 19041175 | Antidepressant | Depression | Drugs |
| 24 HR bupropion hydrochloride 300 MG Extended Release Oral Tablet [Budeprion] | 40221875 | Antidepressant | Depression | Drugs |
| Sprinkle duloxetine 60 MG Delayed Release Oral Capsule | 37496767 | Antidepressant | Depression | Drugs |
| doxepin 6 MG Oral Tablet [Silenor] | 40173389 | Antidepressant | Depression | Drugs |
| amoxapine 50 MG Oral Tablet | 713133 | Antidepressant | Depression | Drugs |
| 24 HR bupropion hydrobromide 522 MG Extended Release Oral Tablet [Aplenzin] | 40221882 | Antidepressant | Depression | Drugs |
| doxepin hydrochloride 100 MG Oral Capsule [Sinequan] | 40224887 | Antidepressant | Depression | Drugs |
| trazodone hydrochloride 300 MG Oral Tablet [Desyrel] | 40163469 | Antidepressant | Depression | Drugs |
| milnacipran hydrochloride 25 MG Oral Tablet | 19080257 | Antidepressant | Depression | Drugs |
| {5 (milnacipran hydrochloride 12.5 MG Oral Tablet) / 8 (milnacipran hydrochloride 25 MG Oral Tablet) / 42 (milnacipran hydrochloride 50 MG Oral Tablet) } Pack | 19133788 | Antidepressant | Depression | Drugs |
| vortioxetine 5 MG Oral Tablet | 44506613 | Antidepressant | Depression | Drugs |
| amitriptyline hydrochloride 25 MG Oral Tablet [Domical] | 19025664 | Antidepressant | Depression | Drugs |
| clomipramine hydrochloride 75 MG Oral Capsule | 40163076 | Antidepressant | Depression | Drugs |
| PMDD fluoxetine 20 MG Oral Tablet | 740118 | Antidepressant | Depression | Drugs |
| 24 HR paroxetine hydrochloride 25 MG Extended Release Oral Tablet [Paxil] | 35604760 | Antidepressant | Depression | Drugs |
| phenelzine 15 MG Oral Tablet [Nardil] | 19004818 | Antidepressant | Depression | Drugs |
| alanine 21.7 MG/ML / arginine 14.7 MG/ML / aspartate 4.34 MG/ML / glutamate 7.49 MG/ML / glycine 10.4 MG/ML / histidine 8.94 MG/ML / isoleucine 7.49 MG/ML / leucine 10.4 MG/ML / lysine 11.8 MG/ML / methionine 7.49 MG/ML / phenylalanine 10.4 MG/ML / pro... | 19130883 | Antidepressant | Depression | Drugs |
| bupropion hydrochloride 300 MG Extended Release Oral Tablet [Budeprion] | 40222085 | Antidepressant | Depression | Drugs |
| desipramine hydrochloride 150 MG Oral Tablet [Norpramin] | 40237429 | Antidepressant | Depression | Drugs |
| escitalopram 1 MG/ML Oral Solution [Lexapro] | 19102744 | Antidepressant | Depression | Drugs |
| desipramine hydrochloride 150 MG Oral Tablet | 40237428 | Antidepressant | Depression | Drugs |
| {5 (milnacipran hydrochloride 12.5 MG Oral Tablet [Savella]) / 8 (milnacipran hydrochloride 25 MG Oral Tablet [Savella]) / 42 (milnacipran hydrochloride 50 MG Oral Tablet [Savella]) } Pack [Savella 4-Week Titration] | 19133789 | Antidepressant | Depression | Drugs |
| 24 HR trazodone hydrochloride 150 MG Extended Release Oral Tablet | 40171493 | Antidepressant | Depression | Drugs |
| clomipramine hydrochloride 75 MG Oral Capsule [Anafranil] | 40163077 | Antidepressant | Depression | Drugs |
| desipramine hydrochloride 100 MG Oral Tablet [Norpramin] | 40237423 | Antidepressant | Depression | Drugs |
| desipramine hydrochloride 25 MG Oral Tablet [Pertofrane] | 40237434 | Antidepressant | Depression | Drugs |

| | | | | |
|---|---|---|---|---|
| doxepin hydrochloride 50 MG/ML Topical Cream [Zonalon] | 40224911 | Antidepressant | Depression | Drugs |
| 5-hydroxytryptophan | 1363516 | Antidepressant | Depression | Drugs |
| clomipramine hydrochloride 50 MG Oral Capsule [Anafranil] | 40163071 | Antidepressant | Depression | Drugs |
| amitriptyline hydrochloride 50 MG Oral Tablet [Elavil] | 40162730 | Antidepressant | Depression | Drugs |
| vilazodone hydrochloride 40 MG Oral Tablet | 40234846 | Antidepressant | Depression | Drugs |
| venlafaxine 50 MG Oral Tablet [Effexor] | 19039820 | Antidepressant | Depression | Drugs |
| nortriptyline 2 MG/ML Oral Solution | 19079114 | Antidepressant | Depression | Drugs |
| fluoxetine 4 MG/ML Oral Solution [Prozac] | 19004896 | Antidepressant | Depression | Drugs |
| imipramine pamoate 100 MG Oral Capsule | 778355 | Antidepressant | Depression | Drugs |
| 24 HR trazodone hydrochloride 300 MG Extended Release Oral Tablet | 40171495 | Antidepressant | Depression | Drugs |
| vortioxetine 10 MG Oral Tablet [Brintellix] | 44506623 | Antidepressant | Depression | Drugs |
| nefazodone hydrochloride 250 MG Oral Tablet | 40238429 | Antidepressant | Depression | Drugs |
| 24 HR fluvoxamine maleate 100 MG Extended Release Oral Capsule [Luvox] | 40173465 | Antidepressant | Depression | Drugs |
| amitriptyline hydrochloride 150 MG Oral Tablet [Endep] | 40162697 | Antidepressant | Depression | Drugs |
| amitriptyline hydrochloride 12.5 MG / chlordiazepoxide 5 MG Oral Tablet | 40162691 | Antidepressant | Depression | Drugs |
| Smoking Cessation 12 HR bupropion hydrochloride 150 MG Extended Release Oral Tablet [Buproban] | 40221861 | Antidepressant | Depression | Drugs |
| alanine 11 MG/ML / arginine 8.5 MG/ML / glycine 11 MG/ML / histidine 2.6 MG/ML / isoleucine 6.2 MG/ML / leucine 8.1 MG/ML / lysine 6.24 MG/ML / methionine 3.4 MG/ML / phenylalanine 3.8 MG/ML / proline 7.5 MG/ML / serine 3.7 MG/ML / threonine 4.6 MG/ML... | 19133498 | Antidepressant | Depression | Drugs |
| desipramine hydrochloride 25 MG Oral Tablet [Norpramin] | 40237433 | Antidepressant | Depression | Drugs |
| milnacipran hydrochloride 50 MG Oral Tablet | 19080255 | Antidepressant | Depression | Drugs |
| vilazodone hydrochloride 20 MG Oral Tablet | 40234842 | Antidepressant | Depression | Drugs |
| nefazodone hydrochloride 50 MG Oral Tablet | 40238435 | Antidepressant | Depression | Drugs |
| desipramine hydrochloride 10 MG Oral Tablet [Norpramin] | 40237419 | Antidepressant | Depression | Drugs |
| {3 (0.2 ML) (esketamine 140 MG/ML Nasal Spray) } Pack | 1366734 | Antidepressant | Depression | Drugs |
| paroxetine hydrochloride 2 MG/ML Oral Suspension [Paxil] | 722066 | Antidepressant | Depression | Drugs |
| fluoxetine 90 MG Delayed Release Oral Capsule | 755741 | Antidepressant | Depression | Drugs |
| mirtazapine 30 MG Disintegrating Oral Tablet [Remeron] | 19128280 | Antidepressant | Depression | Drugs |
| vortioxetine 5 MG Oral Tablet [Brintellix] | 44506616 | Antidepressant | Depression | Drugs |
| fluoxetine 50 MG / olanzapine 12 MG Oral Capsule | 19102554 | Antidepressant | Depression | Drugs |
| milnacipran hydrochloride 100 MG Oral Tablet | 19080230 | Antidepressant | Depression | Drugs |
| paroxetine mesylate 10 MG Oral Tablet | 35604589 | Antidepressant | Depression | Drugs |
| bupropion hydrochloride 200 MG Extended Release Oral Tablet | 40222075 | Antidepressant | Depression | Drugs |
| 24 HR trazodone hydrochloride 300 MG Extended Release Oral Tablet [Oleptro] | 40171496 | Antidepressant | Depression | Drugs |
| paroxetine hydrochloride 2 MG/ML Oral Suspension | 722068 | Antidepressant | Depression | Drugs |
| mirtazapine 45 MG Disintegrating Oral Tablet [Remeron] | 725208 | Antidepressant | Depression | Drugs |
| vortioxetine 20 MG Oral Tablet [Brintellix] | 44506620 | Antidepressant | Depression | Drugs |
| imipramine pamoate 100 MG Oral Capsule [Tofranil-PM] | 778360 | Antidepressant | Depression | Drugs |
| doxepin hydrochloride 10 MG/ML Oral Solution [Sinequan] | 40224882 | Antidepressant | Depression | Drugs |
| {7 (vilazodone hydrochloride 10 MG Oral Tablet [Viibryd]) / 23 (vilazodone hydrochloride 20 MG Oral Tablet [Viibryd]) } Pack [Viibryd Starter Pack 10/20 30 Day Pack] | 46234549 | Antidepressant | Depression | Drugs |
| desipramine hydrochloride 50 MG Oral Tablet [Norpramin] | 40237442 | Antidepressant | Depression | Drugs |
| fluoxetine 50 MG / olanzapine 6 MG Oral Capsule | 19102555 | Antidepressant | Depression | Drugs |
| tryptophan 500 MG Oral Capsule | 19022489 | Antidepressant | Depression | Drugs |
| nefazodone hydrochloride 100 MG Oral Tablet [Serzone] | 40238416 | Antidepressant | Depression | Drugs |
| sertraline 20 MG/ML Oral Solution [Zoloft] | 40165283 | Antidepressant | Depression | Drugs |
| citalopram 2 MG/ML Oral Solution [Celexa] | 797621 | Antidepressant | Depression | Drugs |
| {2 (0.2 ML) (esketamine 140 MG/ML Nasal Spray [Spravato]) } Pack [Spravato 56 MG Dose Kit] | 1366737 | Antidepressant | Depression | Drugs |
| 24 HR fluvoxamine maleate 100 MG Extended Release Oral Capsule | 40173459 | Antidepressant | Depression | Drugs |
| 24 HR bupropion hydrobromide 348 MG Extended Release Oral Tablet | 40221879 | Antidepressant | Depression | Drugs |
| bupropion hydrochloride 150 MG Extended Release Oral Tablet [Zyban] | 40222069 | Antidepressant | Depression | Drugs |
| vilazodone hydrochloride 10 MG Oral Tablet | 40234838 | Antidepressant | Depression | Drugs |
| milnacipran hydrochloride 12.5 MG Oral Tablet [Savella] | 19080254 | Antidepressant | Depression | Drugs |
| amoxapine 100 MG Oral Tablet | 713110 | Antidepressant | Depression | Drugs |
| alanine 7.1 MG/ML / arginine 9.5 MG/ML / cysteine 0.16 MG/ML / glycine 14 MG/ML / histidine 2.8 MG/ML / isoleucine 6.9 MG/ML / leucine 9.1 MG/ML / lysine 7.3 MG/ML / methionine 5.3 MG/ML / phenylalanine 5.6 MG/ML / phosphoric acid 1.2 MG/ML / proline 1... | 40227648 | Antidepressant | Depression | Drugs |
| protriptyline | 754270 | Antidepressant | Depression | Drugs |
| paroxetine mesylate 20 MG Oral Tablet | 35604702 | Antidepressant | Depression | Drugs |
| clomipramine hydrochloride 25 MG Oral Capsule [Anafranil] | 40163064 | Antidepressant | Depression | Drugs |
| imipramine pamoate 150 MG Oral Capsule | 778325 | Antidepressant | Depression | Drugs |
| fluvoxamine maleate 25 MG Oral Tablet [Luvox] | 40174736 | Antidepressant | Depression | Drugs |
| bupropion hydrochloride 150 MG Extended Release Oral Tablet [Wellbutrin] | 40222068 | Antidepressant | Depression | Drugs |
| venlafaxine 37.5 MG Extended Release Oral Tablet | 743760 | Antidepressant | Depression | Drugs |
| paroxetine hydrochloride 12.5 MG Extended Release Oral Tablet | 19115249 | Antidepressant | Depression | Drugs |
| fluoxetine 90 MG Delayed Release Oral Capsule [Prozac] | 19122226 | Antidepressant | Depression | Drugs |
| tranylcypromine | 703470 | Antidepressant | Depression | Drugs |
| nefazodone hydrochloride 250 MG Oral Tablet [Serzone] | 40238430 | Antidepressant | Depression | Drugs |
| nortriptyline 10 MG Oral Capsule [Pamelor] | 19041173 | Antidepressant | Depression | Drugs |

| | | | | |
|---|---|---|---|---|
| alanine 10.4 MG/ML / arginine 5.75 MG/ML / glucose 150 MG/ML / glycine 5.15 MG/ML / histidine 2.4 MG/ML / isoleucine 3 MG/ML / leucine 3.65 MG/ML / lysine 2.9 MG/ML / methionine 2 MG/ML / phenylalanine 2.8 MG/ML / proline 3.4 MG/ML / serine 2.5 MG/ML /... | 19130811 | Antidepressant | Depression | Drugs |
| amitriptyline hydrochloride 100 MG Oral Tablet [Elavil] | 40162684 | Antidepressant | Depression | Drugs |
| fluoxetine 25 MG / olanzapine 6 MG Oral Capsule | 19102552 | Antidepressant | Depression | Drugs |
| 24 HR bupropion hydrobromide 348 MG Extended Release Oral Tablet [Aplenzin] | 40221880 | Antidepressant | Depression | Drugs |
| amoxapine 150 MG Oral Tablet | 713111 | Antidepressant | Depression | Drugs |
| doxepin hydrochloride 100 MG Oral Capsule [Adapin] | 40224886 | Antidepressant | Depression | Drugs |
| alanine 8.8 MG/ML / arginine 4.89 MG/ML / glucose 250 MG/ML / glycine 4.38 MG/ML / histidine 2.04 MG/ML / isoleucine 2.55 MG/ML / leucine 3.11 MG/ML / lysine 2.47 MG/ML / methionine 1.7 MG/ML / phenylalanine 2.38 MG/ML / proline 2.89 MG/ML / serine 2.1... | 19130792 | Antidepressant | Depression | Drugs |
| doxepin hydrochloride 50 MG/ML Topical Cream [Prudoxin] | 40224910 | Antidepressant | Depression | Drugs |
| paroxetine hydrochloride 37.5 MG Extended Release Oral Tablet | 722157 | Antidepressant | Depression | Drugs |
| maprotiline | 794147 | Antidepressant | Depression | Drugs |
| maprotiline hydrochloride 50 MG Oral Tablet [Ludiomil] | 42800989 | Antidepressant | Depression | Drugs |
| nefazodone hydrochloride 300 MG Oral Tablet | 40238433 | Antidepressant | Depression | Drugs |
| nortriptyline 2 MG/ML Oral Solution [Aventyl] | 40239275 | Antidepressant | Depression | Drugs |
| amitriptyline hydrochloride 10 MG/ML Injectable Solution [Elavil] | 40162679 | Antidepressant | Depression | Drugs |
| fluoxetine 10 MG Oral Capsule [Sarafem] | 755821 | Antidepressant | Depression | Drugs |
| fluoxetine 20 MG Oral Capsule [Sarafem] | 755852 | Antidepressant | Depression | Drugs |
| PMDD fluoxetine 15 MG Oral Tablet | 19131597 | Antidepressant | Depression | Drugs |
| fluoxetine 10 MG Oral Tablet [Prozac] | 755734 | Antidepressant | Depression | Drugs |
| trimipramine 25 MG Oral Capsule [Surmontil] | 19039155 | Antidepressant | Depression | Drugs |
| amitriptyline hydrochloride 50 MG / perphenazine 4 MG Oral Tablet [Triavil] | 40162726 | Antidepressant | Depression | Drugs |
| isocarboxazid 10 MG Oral Tablet | 19004819 | Antidepressant | Depression | Drugs |
| trimipramine 50 MG Oral Capsule | 705797 | Antidepressant | Depression | Drugs |
| protriptyline hydrochloride 10 MG Oral Tablet [Vivactil] | 40175377 | Antidepressant | Depression | Drugs |
| imipramine pamoate 75 MG Oral Capsule [Tofranil-PM] | 778323 | Antidepressant | Depression | Drugs |
| nefazodone hydrochloride 50 MG Oral Tablet [Serzone] | 40238436 | Antidepressant | Depression | Drugs |
| alanine 5.4 MG/ML / arginine 12 MG/ML / aspartate 3.2 MG/ML / cysteine 0.24 MG/ML / glutamate 5 MG/ML / glycine 3.6 MG/ML / histidine 4.8 MG/ML / isoleucine 8.2 MG/ML / leucine 1.4 MG/ML / lysine 1.2 MG/ML / methionine 3.4 MG/ML / phenylalanine 4.8 MG/... | 40243299 | Antidepressant | Depression | Drugs |
| desipramine hydrochloride 75 MG Oral Tablet [Norpramin] | 40237446 | Antidepressant | Depression | Drugs |
| imipramine pamoate 150 MG Oral Capsule [Tofranil-PM] | 778326 | Antidepressant | Depression | Drugs |
| paroxetine hydrochloride 25 MG Extended Release Oral Tablet | 722156 | Antidepressant | Depression | Drugs |
| alanine 11 MG/ML / arginine 8.5 MG/ML / glycine 11 MG/ML / histidine 2.6 MG/ML / isoleucine 6.2 MG/ML / leucine 8.1 MG/ML / lysine 6.24 MG/ML / magnesium chloride 0.00502 MEQ/ML / methionine 3.4 MG/ML / phenylalanine 3.8 MG/ML / potassium chloride 0.06... | 40243286 | Antidepressant | Depression | Drugs |
| 24 HR trazodone hydrochloride 150 MG Extended Release Oral Tablet [Oleptro] | 40171494 | Antidepressant | Depression | Drugs |
| alanine 8.8 MG/ML / arginine 4.89 MG/ML / glucose 50 MG/ML / glycine 4.38 MG/ML / histidine 2.04 MG/ML / isoleucine 2.55 MG/ML / leucine 3.11 MG/ML / lysine 2.47 MG/ML / methionine 1.7 MG/ML / phenylalanine 2.38 MG/ML / proline 2.89 MG/ML / serine 2.13... | 19130795 | Antidepressant | Depression | Drugs |
| amitriptyline hydrochloride 25 MG / chlordiazepoxide 10 MG Oral Tablet | 40162701 | Antidepressant | Depression | Drugs |
| alanine 10.4 MG/ML / arginine 5.75 MG/ML / calcium chloride 0.004 MEQ/ML / dibasic potassium phosphate 2.61 MG/ML / glucose 200 MG/ML / glycine 5.15 MG/ML / histidine 2.4 MG/ML / isoleucine 3 MG/ML / leucine 3.65 MG/ML / lysine 2.9 MG/ML / magnesium ch... | 19131089 | Antidepressant | Depression | Drugs |
| 24 HR fluvoxamine maleate 150 MG Extended Release Oral Capsule [Luvox] | 40173467 | Antidepressant | Depression | Drugs |
| bupropion hydrobromide 348 MG Extended Release Oral Tablet | 40222100 | Antidepressant | Depression | Drugs |
| milnacipran hydrochloride 12.5 MG Oral Tablet | 19080253 | Antidepressant | Depression | Drugs |
| Amitriptyline 20 MG/ML | 589159 | Antidepressant | Depression | Drugs |
| amoxapine 25 MG Oral Tablet [Asendin] | 19029977 | Antidepressant | Depression | Drugs |
| {7 (vilazodone hydrochloride 10 MG Oral Tablet) / 23 (vilazodone hydrochloride 20 MG Oral Tablet) } Pack | 46234545 | Antidepressant | Depression | Drugs |
| maprotiline hydrochloride 50 MG Oral Tablet | 42800988 | Antidepressant | Depression | Drugs |
| 5-hydroxytryptophan 100 MG Oral Capsule | 19103431 | Antidepressant | Depression | Drugs |
| fluoxetine 25 MG / olanzapine 3 MG Oral Capsule | 755855 | Antidepressant | Depression | Drugs |
| alanine 27.6 MG/ML / arginine 19.6 MG/ML / aspartate 6 MG/ML / glutamate 10.2 MG/ML / glycine 20.6 MG/ML / histidine 11.8 MG/ML / isoleucine 10.8 MG/ML / leucine 10.8 MG/ML / lysine 13.5 MG/ML / methionine 7.6 MG/ML / phenylalanine 10 MG/ML / proline 1... | 19131138 | Antidepressant | Depression | Drugs |
| duloxetine 30 MG [Cymbalta] | 19060283 | Antidepressant | Depression | Drugs |
| 5-hydroxytryptophan 200 MG Oral Capsule | 43531880 | Antidepressant | Depression | Drugs |
| 24 HR bupropion hydrobromide 174 MG Extended Release Oral Tablet [Aplenzin] | 40221878 | Antidepressant | Depression | Drugs |
| bupropion hydrochloride 100 MG Extended Release Oral Tablet [Wellbutrin] | 40222059 | Antidepressant | Depression | Drugs |
| amitriptyline hydrochloride 10 MG / perphenazine 4 MG Oral Tablet | 40162665 | Antidepressant | Depression | Drugs |
| 5-hydroxytryptophan 50 MG Oral Capsule | 1363520 | Antidepressant | Depression | Drugs |
| amitriptyline hydrochloride 25 MG / perphenazine 2 MG Oral Tablet [Triavil] | 40162707 | Antidepressant | Depression | Drugs |
| amitriptyline hydrochloride 10 MG / perphenazine 2 MG Oral Tablet [Triavil] | 40162661 | Antidepressant | Depression | Drugs |
| 24 HR bupropion hydrobromide 522 MG Extended Release Oral Tablet | 40221881 | Antidepressant | Depression | Drugs |
| bupropion hydrochloride 200 MG Extended Release Oral Tablet [Wellbutrin] | 40222076 | Antidepressant | Depression | Drugs |
| bupropion hydrochloride 300 MG Extended Release Oral Tablet [Wellbutrin] | 40222086 | Antidepressant | Depression | Drugs |

| | | | | |
|---|---|---|---|---|
| bupropion hydrochloride 150 MG Extended Release Oral Tablet [Buproban] | 40222067 | Antidepressant | Depression | Drugs |
| maprotiline hydrochloride 75 MG Oral Tablet | 42800992 | Antidepressant | Depression | Drugs |
| PMDD fluoxetine 10 MG Oral Tablet | 740117 | Antidepressant | Depression | Drugs |
| trimipramine 100 MG Oral Capsule [Surmontil] | 19039154 | Antidepressant | Depression | Drugs |
| amitriptyline hydrochloride 75 MG Oral Tablet [Elavil] | 40162738 | Antidepressant | Depression | Drugs |
| nefazodone hydrochloride 200 MG Oral Tablet [Serzone] | 40238424 | Antidepressant | Depression | Drugs |
| isocarboxazid 10 MG Oral Tablet [Marplan] | 19025733 | Antidepressant | Depression | Drugs |
| 5-hydroxytryptophan 50 MG / magnesium oxide 50 MG / melatonin 2 MG / tryptophan 100 MG / vitamin B6 10 MG Oral Capsule [Somnicin] | 42799061 | Antidepressant | Depression | Drugs |
| maprotiline hydrochloride 25 MG Oral Tablet [Ludiomil] | 42800985 | Antidepressant | Depression | Drugs |
| alanine 4.22 MG/ML / arginine 4.32 MG/ML / aspartate 2.98 MG/ML / glucose 200 MG/ML / glutamate 3.14 MG/ML / glycine 2.12 MG/ML / histidine 1.28 MG/ML / isoleucine 2.8 MG/ML / leucine 4.25 MG/ML / lysine 4.46 MG/ML / methionine 0.73 MG/ML / phenylalani... | 19130567 | Antidepressant | Depression | Drugs |
| alanine 8.8 MG/ML / arginine 4.89 MG/ML / glucose 100 MG/ML / glycine 4.38 MG/ML / histidine 2.04 MG/ML / isoleucine 2.55 MG/ML / leucine 3.11 MG/ML / lysine 2.47 MG/ML / methionine 1.7 MG/ML / phenylalanine 2.38 MG/ML / proline 2.89 MG/ML / serine 2.1... | 19130786 | Antidepressant | Depression | Drugs |
| fluoxetine 25 MG / olanzapine 6 MG Oral Capsule [Symbyax] | 19127038 | Antidepressant | Depression | Drugs |
| duloxetine Oral Capsule | 40058318 | Antidepressant | Depression | Drugs |
| trazodone Oral Tablet [Desyrel] | 40090686 | Antidepressant | Depression | Drugs |
| bupropion Extended Release Oral Tablet [Wellbutrin] | 40102862 | Antidepressant | Depression | Drugs |
| Sprinkle duloxetine 60 MG Delayed Release Oral Capsule [Drizalma] | 37496769 | Antidepressant | Depression | Drugs |
| tryptophan 500 MG Oral Tablet | 19006223 | Antidepressant | Depression | Drugs |
| paroxetine mesylate 40 MG Oral Tablet | 35604708 | Antidepressant | Depression | Drugs |
| {7 (vilazodone hydrochloride 10 MG Oral Tablet) / 7 (vilazodone hydrochloride 20 MG Oral Tablet) / 16 (vilazodone hydrochloride 40 MG Oral Tablet) } Pack | 40235111 | Antidepressant | Depression | Drugs |
| paroxetine mesylate 30 MG Oral Tablet | 35604705 | Antidepressant | Depression | Drugs |
| 24 HR bupropion hydrobromide 174 MG Extended Release Oral Tablet | 40221877 | Antidepressant | Depression | Drugs |
| tryptophan | 19006186 | Antidepressant | Depression | Drugs |
| tryptophan 500 MG Oral Tablet [Pacitron] | 19024357 | Antidepressant | Depression | Drugs |
| amoxapine 100 MG Oral Tablet [Asendin] | 19029947 | Antidepressant | Depression | Drugs |
| amitriptyline hydrochloride 12.5 MG / chlordiazepoxide 5 MG Oral Tablet [Limbitrol] | 40162692 | Antidepressant | Depression | Drugs |
| amoxapine 50 MG Oral Tablet [Asendin] | 19029980 | Antidepressant | Depression | Drugs |
| fluoxetine 50 MG / olanzapine 6 MG Oral Capsule [Symbyax] | 19127042 | Antidepressant | Depression | Drugs |
| Sprinkle duloxetine 20 MG Delayed Release Oral Capsule [Drizalma] | 37496760 | Antidepressant | Depression | Drugs |
| trimipramine 25 MG Oral Capsule | 705796 | Antidepressant | Depression | Drugs |
| amoxapine 150 MG Oral Tablet [Asendin] | 19029974 | Antidepressant | Depression | Drugs |
| amitriptyline hydrochloride 25 MG / perphenazine 4 MG Oral Tablet | 40162711 | Antidepressant | Depression | Drugs |
| maprotiline hydrochloride 75 MG Oral Tablet [Ludiomil] | 42800993 | Antidepressant | Depression | Drugs |
| alanine 10.4 MG/ML / arginine 5.75 MG/ML / calcium chloride 0.004 MEQ/ML / dibasic potassium phosphate 2.61 MG/ML / glucose 350 MG/ML / glycine 5.15 MG/ML / histidine 2.4 MG/ML / isoleucine 3 MG/ML / leucine 3.65 MG/ML / magnesium chloride 0.01 MEQ/ML... | 19131095 | Antidepressant | Depression | Drugs |
| alanine 2.1 MG/ML / arginine 2.9 MG/ML / calcium acetate 0.003 MEQ/ML / cysteine 0.2 MG/ML / glycerin 30 MG/ML / glycine 4.2 MG/ML / histidine 0.85 MG/ML / isoleucine 2.1 MG/ML / leucine 2.7 MG/ML / lysine 3.1 MG/ML / magnesium acetate 0.008 MEQ/ML / m... | 19131215 | Antidepressant | Depression | Drugs |
| alanine 8.8 MG/ML / arginine 4.89 MG/ML / calcium chloride 0.004 MEQ/ML / dibasic potassium phosphate 2.61 MG/ML / glucose 50 MG/ML / glycine 4.38 MG/ML / histidine 2.04 MG/ML / isoleucine 2.55 MG/ML / leucine 3.11 MG/ML / lysine 2.47 MG/ML / magnesium... | 19131062 | Antidepressant | Depression | Drugs |
| alanine 8.8 MG/ML / arginine 4.89 MG/ML / glucose 50 MG/ML / glycine 4.38 MG/ML / histidine 2.04 MG/ML / isoleucine 2.55 MG/ML / leucine 3.11 MG/ML / lysine 2.47 MG/ML / methionine 1.7 MG/ML / phenylalanine 2.38 MG/ML / proline 2.89 MG/ML / serine 2.13... | 19130793 | Antidepressant | Depression | Drugs |
| imipramine hydrochloride 25 MG Oral Tablet [Pramimil] | 19000966 | Antidepressant | Depression | Drugs |
| citalopram 10 MG Oral Tablet [Cipramil] | 19017328 | Antidepressant | Depression | Drugs |
| imipramine pamoate 125 MG Oral Capsule | 778353 | Antidepressant | Depression | Drugs |
| bupropion Extended Release Oral Tablet [Buproban] | 40125347 | Antidepressant | Depression | Drugs |
| alanine 2.1 MG/ML / arginine 2.9 MG/ML / calcium acetate 0.003 MEQ/ML / cysteine 0.2 MG/ML / glycerin 30 MG/ML / glycine 4.2 MG/ML / histidine 0.85 MG/ML / isoleucine 2.1 MG/ML / leucine 2.7 MG/ML / lysine 3.1 MG/ML / magnesium acetate 0.008 MEQ/ML / m... | 19131217 | Antidepressant | Depression | Drugs |
| alanine 21.7 MG/ML / arginine 14.7 MG/ML / aspartate 4.34 MG/ML / glutamate 7.49 MG/ML / glycine 10.4 MG/ML / histidine 8.94 MG/ML / isoleucine 7.49 MG/ML / leucine 10.4 MG/ML / lysine 11.8 MG/ML / methionine 7.49 MG/ML / phenylalanine 10.4 MG/ML / pro... | 45892124 | Antidepressant | Depression | Drugs |
| citalopram 20 MG Oral Tablet [Cipramil] | 19010672 | Antidepressant | Depression | Drugs |
| alanine 10.4 MG/ML / arginine 5.75 MG/ML / calcium chloride 0.004 MEQ/ML / dibasic potassium phosphate 2.61 MG/ML / glucose 150 MG/ML / glycine 5.15 MG/ML / histidine 2.4 MG/ML / isoleucine 3 MG/ML / leucine 3.65 MG/ML / lysine 2.9 MG/ML / magnesium ch... | 19131080 | Antidepressant | Depression | Drugs |
| alanine 20.7 MG/ML / arginine 11.5 MG/ML / glycine 10.3 MG/ML / histidine 4.8 MG/ML / isoleucine 6 MG/ML / leucine 7.3 MG/ML / lysine 5.8 MG/ML / methionine 4 MG/ML / phenylalanine 5.6 MG/ML / proline 6.8 MG/ML / serine 5 MG/ML / threonine 4.2 MG/ML /... | 19131341 | Antidepressant | Depression | Drugs |

| | | | | |
|---|---|---|---|---|
| alanine 12.8 MG/ML / arginine 9.8 MG/ML / glycine 12.8 MG/ML / histidine 3 MG/ML / isoleucine 7.2 MG/ML / leucine 9.4 MG/ML / lysine 7.2 MG/ML / methionine 4 MG/ML / phenylalanine 4.4 MG/ML / proline 8.6 MG/ML / serine 4.2 MG/ML / threonine 5.2 MG/ML /... | 19133481 | Antidepressant | Depression | Drugs |
| imipramine hydrochloride 10 MG Oral Tablet [Pramimil] | 19000965 | Antidepressant | Depression | Drugs |
| alanine 10.4 MG/ML / arginine 5.75 MG/ML / calcium chloride 0.004 MEQ/ML / dibasic potassium phosphate 2.61 MG/ML / glucose 200 MG/ML / glycine 5.15 MG/ML / histidine 2.4 MG/ML / isoleucine 3 MG/ML / leucine 3.65 MG/ML / lysine 2.9 MG/ML / magnesium ch... | 19131087 | Antidepressant | Depression | Drugs |
| alanine 5.7 MG/ML / arginine 3.16 MG/ML / calcium chloride 0.004 MEQ/ML / dibasic potassium phosphate 2.61 MG/ML / glucose 50 MG/ML / glycine 2.83 MG/ML / histidine 1.32 MG/ML / isoleucine 1.65 MG/ML / leucine 2.01 MG/ML / lysine 1.59 MG/ML / magnesium... | 19131071 | Antidepressant | Depression | Drugs |
| duloxetine 20 MG Oral Capsule | 19112657 | Antidepressant | Depression | Drugs |
| vortioxetine Oral Tablet [Trintellix] | 42629568 | Antidepressant | Depression | Drugs |
| alanine 8.44 MG/ML / arginine 8.65 MG/ML / aspartate 5.95 MG/ML / glutamate 6.27 MG/ML / glycine 4.25 MG/ML / histidine 2.55 MG/ML / isoleucine 5.61 MG/ML / leucine 8.5 MG/ML / lysine 8.93 MG/ML / methionine 1.46 MG/ML / phenylalanine 2.53 MG/ML / prol... | 19130651 | Antidepressant | Depression | Drugs |
| venlafaxine 75 MG [Effexor] | 19118743 | Antidepressant | Depression | Drugs |
| amitriptyline 30 MG | 19109687 | Antidepressant | Depression | Drugs |
| clomipramine hydrochloride 50 MG | 40163069 | Antidepressant | Depression | Drugs |
| doxepin 100 MG | 19085200 | Antidepressant | Depression | Drugs |
| nortriptyline 50 MG [Pamelor] | 19026384 | Antidepressant | Depression | Drugs |
| mirtazapine 45 MG [Remeron] | 19015749 | Antidepressant | Depression | Drugs |
| trazodone hydrochloride 50 MG | 40163471 | Antidepressant | Depression | Drugs |
| trazodone hydrochloride 100 MG | 40163458 | Antidepressant | Depression | Drugs |
| fluoxetine 40 MG [Prozac] | 19080284 | Antidepressant | Depression | Drugs |
| fluoxetine 20 MG [Prozac] | 19080281 | Antidepressant | Depression | Drugs |
| doxepin 10 MG | 738240 | Antidepressant | Depression | Drugs |
| fluoxetine 60 MG | 19091121 | Antidepressant | Depression | Drugs |
| amitriptyline hydrochloride 50 MG | 40162724 | Antidepressant | Depression | Drugs |
| escitalopram 20 MG [Lexapro] | 19033450 | Antidepressant | Depression | Drugs |
| sertraline 50 MG [Zoloft] | 19043842 | Antidepressant | Depression | Drugs |
| bupropion hydrochloride 100 MG [Wellbutrin] | 40222063 | Antidepressant | Depression | Drugs |
| sertraline 100 MG | 19083100 | Antidepressant | Depression | Drugs |
| bupropion hydrochloride 200 MG [Wellbutrin] | 40222077 | Antidepressant | Depression | Drugs |
| nefazodone hydrochloride 150 MG | 40238418 | Antidepressant | Depression | Drugs |
| clomipramine hydrochloride 25 MG [Anafranil] | 40163066 | Antidepressant | Depression | Drugs |
| nortriptyline 25 MG | 19083195 | Antidepressant | Depression | Drugs |
| imipramine hydrochloride 25 MG | 778295 | Antidepressant | Depression | Drugs |
| nortriptyline 10 MG | 721761 | Antidepressant | Depression | Drugs |
| clomipramine hydrochloride 50 MG [Anafranil] | 40163073 | Antidepressant | Depression | Drugs |
| bupropion hydrochloride 150 MG | 40222064 | Antidepressant | Depression | Drugs |
| doxepin 50 MG | 738262 | Antidepressant | Depression | Drugs |
| fluvoxamine maleate 100 MG [Luvox] | 40174727 | Antidepressant | Depression | Drugs |
| mirtazapine 30 MG [Remeron] | 19015748 | Antidepressant | Depression | Drugs |
| venlafaxine 37.5 MG [Effexor] | 19021629 | Antidepressant | Depression | Drugs |
| escitalopram 10 MG [Lexapro] | 19033449 | Antidepressant | Depression | Drugs |
| bupropion hydrochloride 300 MG | 40222083 | Antidepressant | Depression | Drugs |
| bupropion hydrochloride 150 MG [Wellbutrin] | 40222072 | Antidepressant | Depression | Drugs |
| citalopram 20 MG [Celexa] | 19120296 | Antidepressant | Depression | Drugs |
| nortriptyline 30 MG | 721796 | Antidepressant | Depression | Drugs |
| sertraline 25 MG | 739282 | Antidepressant | Depression | Drugs |
| bupropion hydrochloride 75 MG [Wellbutrin] | 40222094 | Antidepressant | Depression | Drugs |
| doxepin 25 MG | 738241 | Antidepressant | Depression | Drugs |
| duloxetine 20 MG [Cymbalta] | 19060282 | Antidepressant | Depression | Drugs |
| venlafaxine 150 MG [Effexor] | 19127359 | Antidepressant | Depression | Drugs |
| amitriptyline 25 MG | 19085723 | Antidepressant | Depression | Drugs |
| citalopram 10 MG [Celexa] | 19120749 | Antidepressant | Depression | Drugs |
| sertraline 100 MG [Zoloft] | 19043841 | Antidepressant | Depression | Drugs |
| sertraline 25 MG [Zoloft] | 19043843 | Antidepressant | Depression | Drugs |
| imipramine hydrochloride 50 MG | 778298 | Antidepressant | Depression | Drugs |
| nortriptyline 75 MG | 721794 | Antidepressant | Depression | Drugs |
| imipramine hydrochloride 10 MG | 778356 | Antidepressant | Depression | Drugs |
| mirtazapine 45 MG | 725203 | Antidepressant | Depression | Drugs |
| fluoxetine 10 MG | 755762 | Antidepressant | Depression | Drugs |
| amitriptyline 75 MG | 710183 | Antidepressant | Depression | Drugs |
| amitriptyline 10 MG | 710185 | Antidepressant | Depression | Drugs |
| amitriptyline hydrochloride 25 MG [Elavil] | 40162720 | Antidepressant | Depression | Drugs |
| nefazodone hydrochloride 200 MG [Serzone] | 40238425 | Antidepressant | Depression | Drugs |
| duloxetine 60 MG [Cymbalta] | 19060284 | Antidepressant | Depression | Drugs |
| citalopram 40 MG | 797637 | Antidepressant | Depression | Drugs |
| bupropion hydrochloride 150 MG [Zyban] | 40222073 | Antidepressant | Depression | Drugs |

| | | | | |
|---|---|---|---|---|
| fluoxetine 40 MG | 19084977 | Antidepressant | Depression | Drugs |
| bupropion hydrochloride 300 MG [Wellbutrin] | 40222088 | Antidepressant | Depression | Drugs |
| citalopram 40 MG [Celexa] | 19058160 | Antidepressant | Depression | Drugs |
| trazodone hydrochloride 150 MG [Desyrel] | 40163466 | Antidepressant | Depression | Drugs |
| bupropion hydrochloride 100 MG | 40222056 | Antidepressant | Depression | Drugs |
| amitriptyline hydrochloride 100 MG | 40162682 | Antidepressant | Depression | Drugs |
| bupropion hydrochloride 200 MG | 40222074 | Antidepressant | Depression | Drugs |
| fluoxetine 20 MG | 755763 | Antidepressant | Depression | Drugs |
| mirtazapine 15 MG [Remeron] | 19015747 | Antidepressant | Depression | Drugs |
| mirtazapine 15 MG | 19095236 | Antidepressant | Depression | Drugs |
| clomipramine hydrochloride 25 MG | 40163062 | Antidepressant | Depression | Drugs |
| nefazodone hydrochloride 150 MG [Serzone] | 40238421 | Antidepressant | Depression | Drugs |
| trimipramine | 705755 | Antidepressant | Depression | Drugs |
| fluoxetine 15 MG | 19131596 | Antidepressant | Depression | Drugs |
| imipramine hydrochloride 150 MG | 19135757 | Antidepressant | Depression | Drugs |
| sertraline 200 MG | 739283 | Antidepressant | Depression | Drugs |
| nortriptyline 25 MG [Pamelor] | 19118843 | Antidepressant | Depression | Drugs |
| nortriptyline 75 MG [Pamelor] | 19118855 | Antidepressant | Depression | Drugs |
| protriptyline hydrochloride 10 MG [Vivactil] | 40175378 | Antidepressant | Depression | Drugs |
| desipramine 50 MG | 19082930 | Antidepressant | Depression | Drugs |
| bupropion hydrochloride 50 MG | 40222089 | Antidepressant | Depression | Drugs |
| fluvoxamine maleate 50 MG [Luvox] | 40174741 | Antidepressant | Depression | Drugs |
| protriptyline hydrochloride 10 MG | 40175375 | Antidepressant | Depression | Drugs |
| fluoxetine 20 MG [Sarafem] | 19090639 | Antidepressant | Depression | Drugs |
| imipramine hydrochloride 10 MG [Tofranil] | 19001275 | Antidepressant | Depression | Drugs |
| nortriptyline 50 MG | 721793 | Antidepressant | Depression | Drugs |
| bupropion hydrochloride 100 MG [Budeprion] | 40222062 | Antidepressant | Depression | Drugs |
| amitriptyline hydrochloride 10 MG [Elavil] | 40162675 | Antidepressant | Depression | Drugs |
| nefazodone hydrochloride 100 MG [Serzone] | 40238417 | Antidepressant | Depression | Drugs |
| imipramine hydrochloride 75 MG | 19135759 | Antidepressant | Depression | Drugs |
| duloxetine 30 MG | 715296 | Antidepressant | Depression | Drugs |
| amitriptyline hydrochloride 150 MG | 40162694 | Antidepressant | Depression | Drugs |
| fluvoxamine maleate 150 MG | 40174728 | Antidepressant | Depression | Drugs |
| clomipramine hydrochloride 20 MG | 40169581 | Antidepressant | Depression | Drugs |
| fluoxetine 50 MG | 19103071 | Antidepressant | Depression | Drugs |
| desipramine 25 MG | 19082929 | Antidepressant | Depression | Drugs |
| trazodone hydrochloride 50 MG [Desyrel] | 40163475 | Antidepressant | Depression | Drugs |
| trazodone 75 MG | 703615 | Antidepressant | Depression | Drugs |
| trazodone hydrochloride 150 MG | 40163463 | Antidepressant | Depression | Drugs |
| fluvoxamine 50 MG | 751419 | Antidepressant | Depression | Drugs |
| trazodone 25 MG | 19109758 | Antidepressant | Depression | Drugs |
| fluoxetine 10 MG [Prozac] | 19080283 | Antidepressant | Depression | Drugs |
| fluoxetine 60 MG [Prozac] | 19116858 | Antidepressant | Depression | Drugs |
| bupropion hydrochloride 150 MG [Budeprion] | 40222070 | Antidepressant | Depression | Drugs |
| escitalopram 5 MG [Lexapro] | 19121118 | Antidepressant | Depression | Drugs |
| mirtazapine 30 MG | 19095237 | Antidepressant | Depression | Drugs |
| nortriptyline 20 MG | 721797 | Antidepressant | Depression | Drugs |
| citalopram 20 MG | 797636 | Antidepressant | Depression | Drugs |
| doxepin 75 MG | 738263 | Antidepressant | Depression | Drugs |
| citalopram 10 MG | 797639 | Antidepressant | Depression | Drugs |
| fluvoxamine maleate 150 MG [Luvox] | 40174733 | Antidepressant | Depression | Drugs |
| sertraline 50 MG | 739211 | Antidepressant | Depression | Drugs |
| venlafaxine 150 MG | 743759 | Antidepressant | Depression | Drugs |
| fluvoxamine maleate 25 MG | 40174734 | Antidepressant | Depression | Drugs |
| sertraline 150 MG | 739284 | Antidepressant | Depression | Drugs |
| fluoxetine 4 MG/ML [Prozac] | 19080282 | Antidepressant | Depression | Drugs |
| nefazodone hydrochloride 250 MG [Serzone] | 40238431 | Antidepressant | Depression | Drugs |
| venlafaxine 225 MG | 19131402 | Antidepressant | Depression | Drugs |
| clomipramine hydrochloride 75 MG [Anafranil] | 40163078 | Antidepressant | Depression | Drugs |
| amitriptyline hydrochloride 50 MG [Elavil] | 40162733 | Antidepressant | Depression | Drugs |
| sertraline 20 MG/ML [Zoloft] | 40165284 | Antidepressant | Depression | Drugs |
| venlafaxine 37.5 MG | 743755 | Antidepressant | Depression | Drugs |
| tranylcypromine 10 MG [Parnate] | 19115965 | Antidepressant | Depression | Drugs |
| fluvoxamine 100 MG | 751420 | Antidepressant | Depression | Drugs |
| imipramine hydrochloride 25 MG [Tofranil] | 19001272 | Antidepressant | Depression | Drugs |
| venlafaxine 75 MG | 19083160 | Antidepressant | Depression | Drugs |
| nefazodone hydrochloride 300 MG | 40238432 | Antidepressant | Depression | Drugs |
| protriptyline hydrochloride 5 MG | 40175379 | Antidepressant | Depression | Drugs |
| desipramine hydrochloride 100 MG | 40237421 | Antidepressant | Depression | Drugs |
| nortriptyline 10 MG [Pamelor] | 19026382 | Antidepressant | Depression | Drugs |
| desipramine 40 MG | 717063 | Antidepressant | Depression | Drugs |
| trazodone hydrochloride 300 MG [Desyrel] | 40163470 | Antidepressant | Depression | Drugs |

| | | | | |
|---|---|---|---|---|
| desipramine dibudinate 30 MG | 40237415 | Antidepressant | Depression | Drugs |
| desipramine hydrochloride 10 MG | 40237417 | Antidepressant | Depression | Drugs |
| fluoxetine 90 MG | 755766 | Antidepressant | Depression | Drugs |
| bupropion hydrochloride 450 MG | 42705084 | Antidepressant | Depression | Drugs |
| sertraline Oral Tablet | 40081943 | Antidepressant | Depression | Drugs |
| duloxetine 20 MG | 715293 | Antidepressant | Depression | Drugs |
| nefazodone hydrochloride 50 MG [Serzone] | 40238437 | Antidepressant | Depression | Drugs |
| trazodone hydrochloride 300 MG | 40163467 | Antidepressant | Depression | Drugs |
| fluoxetine 10 MG [Sarafem] | 19090638 | Antidepressant | Depression | Drugs |
| trazodone 10 MG/ML | 703614 | Antidepressant | Depression | Drugs |
| nortriptyline 40 MG | 721798 | Antidepressant | Depression | Drugs |
| isocarboxazid | 781705 | Antidepressant | Depression | Drugs |
| doxepin 40 MG/ML | 19109670 | Antidepressant | Depression | Drugs |
| venlafaxine 25 MG [Effexor] | 19021628 | Antidepressant | Depression | Drugs |
| venlafaxine 50 MG [Effexor] | 19118741 | Antidepressant | Depression | Drugs |
| fluoxetine 90 MG [Prozac] | 19113117 | Antidepressant | Depression | Drugs |
| amitriptyline hydrochloride 5 MG/ML | 40162722 | Antidepressant | Depression | Drugs |
| fluvoxamine maleate 100 MG | 40174722 | Antidepressant | Depression | Drugs |
| imipramine pamoate 100 MG | 19134044 | Antidepressant | Depression | Drugs |
| venlafaxine 100 MG [Effexor] | 19021627 | Antidepressant | Depression | Drugs |
| amitriptyline hydrochloride 12.5 MG | 40162688 | Antidepressant | Depression | Drugs |
| protriptyline hydrochloride 5 MG [Vivactil] | 40175382 | Antidepressant | Depression | Drugs |
| fluoxetine 25 MG / olanzapine 12 MG Oral Capsule | 19102553 | Antidepressant | Depression | Drugs |
| nefazodone hydrochloride 200 MG | 40238422 | Antidepressant | Depression | Drugs |
| citalopram 2 MG/ML [Celexa] | 19058161 | Antidepressant | Depression | Drugs |
| imipramine pamoate 125 MG | 778352 | Antidepressant | Depression | Drugs |
| phenelzine 15 MG [Nardil] | 19048255 | Antidepressant | Depression | Drugs |
| clomipramine 75 MG | 19085292 | Antidepressant | Depression | Drugs |
| doxepin hydrochloride 150 MG | 40224890 | Antidepressant | Depression | Drugs |
| desipramine hydrochloride 150 MG | 40237427 | Antidepressant | Depression | Drugs |
| amitriptyline 15 MG | 710263 | Antidepressant | Depression | Drugs |
| fluvoxamine maleate 150 MG Extended Release Oral Capsule | 40174729 | Antidepressant | Depression | Drugs |
| trazodone hydrochloride 100 MG [Desyrel] | 40163462 | Antidepressant | Depression | Drugs |
| fluoxetine 50 MG / olanzapine 12 MG Oral Capsule [Symbyax] | 19127040 | Antidepressant | Depression | Drugs |
| trazodone Oral Solution | 40090682 | Antidepressant | Depression | Drugs |
| Sprinkle duloxetine 30 MG Delayed Release Oral Capsule [Drizalma] | 37496763 | Antidepressant | Depression | Drugs |
| alanine 9.93 MG/ML / arginine 10.2 MG/ML / aspartate 7 MG/ML / glutamate 7.38 MG/ML / glycine 5 MG/ML / histidine 3 MG/ML / isoleucine 6.6 MG/ML / leucine 10 MG/ML / lysine 10.5 MG/ML / methionine 1.72 MG/ML / phenylalanine 2.98 MG/ML / proline 7.22 MG... | 19130670 | Antidepressant | Depression | Drugs |
| maprotiline hydrochloride 25 MG | 42800983 | Antidepressant | Depression | Drugs |
| bupropion hydrochloride 75 MG | 40222091 | Antidepressant | Depression | Drugs |
| escitalopram 10 MG | 19098080 | Antidepressant | Depression | Drugs |
| esketamine 140 MG/ML Nasal Spray | 1366618 | Antidepressant | Depression | Drugs |
| escitalopram 20 MG | 715968 | Antidepressant | Depression | Drugs |
| mirtazapine 7.5 MG | 19112587 | Antidepressant | Depression | Drugs |
| duloxetine 60 MG | 715299 | Antidepressant | Depression | Drugs |
| desipramine hydrochloride 75 MG | 40237444 | Antidepressant | Depression | Drugs |
| amitriptyline 75 MG [Elavil] | 19117641 | Antidepressant | Depression | Drugs |
| paroxetine Oral Tablet | 40071301 | Antidepressant | Depression | Drugs |
| doxepin hydrochloride 25 MG [Sinequan] | 40224901 | Antidepressant | Depression | Drugs |
| amitriptyline hydrochloride 10 MG Oral Tablet [Endep] | 40162674 | Antidepressant | Depression | Drugs |
| Sprinkle duloxetine 40 MG Delayed Release Oral Capsule [Drizalma] | 37496766 | Antidepressant | Depression | Drugs |
| bupropion hydrochloride 300 MG [Budeprion] | 40222087 | Antidepressant | Depression | Drugs |
| amitriptyline 60 MG | 19110100 | Antidepressant | Depression | Drugs |
| fluvoxamine maleate 25 MG [Luvox] | 40174737 | Antidepressant | Depression | Drugs |
| venlafaxine 100 MG | 743754 | Antidepressant | Depression | Drugs |
| citalopram 2 MG/ML | 19084879 | Antidepressant | Depression | Drugs |
| venlafaxine 25 MG | 19086246 | Antidepressant | Depression | Drugs |
| maprotiline hydrochloride 50 MG [Ludiomil] | 42800990 | Antidepressant | Depression | Drugs |
| amitriptyline 90 MG | 19109688 | Antidepressant | Depression | Drugs |
| maprotiline hydrochloride 75 MG | 42800991 | Antidepressant | Depression | Drugs |
| nortriptyline 10 MG Oral Tablet | 721728 | Antidepressant | Depression | Drugs |
| alanine 9.93 MG/ML / arginine 10.2 MG/ML / aspartate 7 MG/ML / glutamate 7.38 MG/ML / glycine 5 MG/ML / histidine 3 MG/ML / isoleucine 6.6 MG/ML / leucine 10 MG/ML / lysine 10.5 MG/ML / methionine 1.72 MG/ML / phenylalanine 2.98 MG/ML / proline 7.22 MG... | 19130668 | Antidepressant | Depression | Drugs |
| tryptophan Oral Tablet | 40088846 | Antidepressant | Depression | Drugs |
| nefazodone hydrochloride 100 MG | 40238414 | Antidepressant | Depression | Drugs |
| amitriptyline 2 MG/ML | 19087006 | Antidepressant | Depression | Drugs |
| alanine 21.7 MG/ML / arginine 14.7 MG/ML / aspartate 4.34 MG/ML / glutamate 7.49 MG/ML / glycine 10.4 MG/ML / histidine 8.94 MG/ML / isoleucine 7.49 MG/ML / leucine 10.4 MG/ML / lysine 11.8 MG/ML / methionine 7.49 MG/ML / phenylalanine 10.4 MG/ML / pro... | 19130881 | Antidepressant | Depression | Drugs |

| | | | | | |
|---|---|---|---|---|---|
| nortriptyline 25 MG [Aventyl] | 19115958 | Antidepressant | Depression | Drugs | |
| fluoxetine 6 MG | 755770 | Antidepressant | Depression | Drugs | |
| nefazodone hydrochloride 25 MG | 40238426 | Antidepressant | Depression | Drugs | |
| fluoxetine 25 MG | 19103068 | Antidepressant | Depression | Drugs | |
| fluoxetine hydrochloride 16 MG | 40174701 | Antidepressant | Depression | Drugs | |
| fluoxetine 25 MG / olanzapine 3 MG Oral Capsule [Symbyax] | 19127046 | Antidepressant | Depression | Drugs | |
| desvenlafaxine 100 MG Extended Release Oral Tablet [Pristiq] | 19134912 | Antidepressant | Depression | Drugs | |
| alanine 5.4 MG/ML / arginine 12 MG/ML / aspartate 3.2 MG/ML / cysteine 0.24 MG/ML / glutamate 5 MG/ML / glycine 3.6 MG/ML / histidine 4.8 MG/ML / isoleucine 8.2 MG/ML / leucine 1.4 MG/ML / lysine 1.2 MG/ML / methionine 3.4 MG/ML / phenylalanine 4.8 MG/... | 19130902 | Antidepressant | Depression | Drugs | |
| doxepin 5 MG | 19109695 | Antidepressant | Depression | Drugs | |
| clomipramine hydrochloride 10 MG | 40163057 | Antidepressant | Depression | Drugs | |
| nefazodone hydrochloride 50 MG | 40238434 | Antidepressant | Depression | Drugs | |
| venlafaxine 50 MG | 19083159 | Antidepressant | Depression | Drugs | |
| amitriptyline 120 MG | 19109686 | Antidepressant | Depression | Drugs | |
| bupropion | 750982 | Antidepressant | Depression | Drugs | |
| trazodone | 703547 | Antidepressant | Depression | Drugs | |
| citalopram | 797617 | Antidepressant | Depression | Drugs | |
| duloxetine | 715259 | Antidepressant | Depression | Drugs | |
| fluoxetine | 755695 | Antidepressant | Depression | Drugs | |
| paroxetine | 722031 | Antidepressant | Depression | Drugs | |
| sertraline | 739138 | Antidepressant | Depression | Drugs | |
| mirtazapine | 725131 | Antidepressant | Depression | Drugs | |
| venlafaxine | 743670 | Antidepressant | Depression | Drugs | |
| escitalopram | 715939 | Antidepressant | Depression | Drugs | |
| amitriptyline | 710062 | Antidepressant | Depression | Drugs | |
| nortriptyline | 721724 | Antidepressant | Depression | Drugs | |
| citalopram 10 MG Oral Tablet | 797632 | Antidepressant | Depression | Drugs | |
| citalopram 20 MG Oral Tablet | 19023636 | Antidepressant | Depression | Drugs | |
| citalopram 40 MG Oral Tablet | 19075394 | Antidepressant | Depression | Drugs | |
| sertraline 25 MG Oral Tablet | 19079497 | Antidepressant | Depression | Drugs | |
| sertraline 50 MG Oral Tablet | 739209 | Antidepressant | Depression | Drugs | |
| fluoxetine 10 MG Oral Capsule | 19077462 | Antidepressant | Depression | Drugs | |
| fluoxetine 20 MG Oral Capsule | 19077463 | Antidepressant | Depression | Drugs | |
| fluoxetine 40 MG Oral Capsule | 755739 | Antidepressant | Depression | Drugs | |
| mirtazapine 15 MG Oral Tablet | 725178 | Antidepressant | Depression | Drugs | |
| mirtazapine 30 MG Oral Tablet | 725180 | Antidepressant | Depression | Drugs | |
| sertraline 100 MG Oral Tablet | 739207 | Antidepressant | Depression | Drugs | |
| escitalopram 10 MG Oral Tablet | 715940 | Antidepressant | Depression | Drugs | |
| escitalopram 20 MG Oral Tablet | 19098166 | Antidepressant | Depression | Drugs | |
| nortriptyline 10 MG Oral Capsule | 19019412 | Antidepressant | Depression | Drugs | |
| nortriptyline 25 MG Oral Capsule | 721760 | Antidepressant | Depression | Drugs | |
| sertraline 50 MG Oral Tablet [Zoloft] | 19037684 | Antidepressant | Depression | Drugs | |
| sertraline 100 MG Oral Tablet [Zoloft] | 19037642 | Antidepressant | Depression | Drugs | |
| trazodone hydrochloride 50 MG Oral Tablet | 40163473 | Antidepressant | Depression | Drugs | |
| paroxetine hydrochloride 20 MG Oral Tablet | 35604576 | Antidepressant | Depression | Drugs | |
| paroxetine hydrochloride 40 MG Oral Tablet | 35604586 | Antidepressant | Depression | Drugs | |
| trazodone hydrochloride 100 MG Oral Tablet | 40163460 | Antidepressant | Depression | Drugs | |
| trazodone hydrochloride 150 MG Oral Tablet | 40163464 | Antidepressant | Depression | Drugs | |
| amitriptyline hydrochloride 10 MG Oral Tablet | 40162672 | Antidepressant | Depression | Drugs | |
| amitriptyline hydrochloride 25 MG Oral Tablet | 40162717 | Antidepressant | Depression | Drugs | |
| amitriptyline hydrochloride 50 MG Oral Tablet | 40162729 | Antidepressant | Depression | Drugs | |
| duloxetine 20 MG Delayed Release Oral Capsule | 715294 | Antidepressant | Depression | Drugs | |
| duloxetine 30 MG Delayed Release Oral Capsule | 19122148 | Antidepressant | Depression | Drugs | |
| duloxetine 60 MG Delayed Release Oral Capsule | 715300 | Antidepressant | Depression | Drugs | |
| 24 HR venlafaxine 75 MG Extended Release Oral Capsule | 743721 | Antidepressant | Depression | Drugs | |
| 24 HR venlafaxine 150 MG Extended Release Oral Capsule | 743717 | Antidepressant | Depression | Drugs | |
| 24 HR venlafaxine 37.5 MG Extended Release Oral Capsule | 743719 | Antidepressant | Depression | Drugs | |
| duloxetine 60 MG Delayed Release Oral Capsule [Cymbalta] | 715301 | Antidepressant | Depression | Drugs | |
| 12 HR bupropion hydrochloride 100 MG Extended Release Oral Tablet | 40221856 | Antidepressant | Depression | Drugs | |
| 12 HR bupropion hydrochloride 150 MG Extended Release Oral Tablet | 40221859 | Antidepressant | Depression | Drugs | |
| 24 HR bupropion hydrochloride 150 MG Extended Release Oral Tablet | 40221871 | Antidepressant | Depression | Drugs | |
| 24 HR bupropion hydrochloride 300 MG Extended Release Oral Tablet | 40221874 | Antidepressant | Depression | Drugs | |
| Recurrent major depressive episodes, moderate | 432883 | Depression | Depression | Conditions | Excluded |
| Recurrent major depressive episodes | 432285 | Depression | Depression | Conditions | Excluded |
| Recurrent major depression in partial remission | 4141454 | Depression | Depression | Conditions | Excluded |
| Depressive disorder | 440383 | Depression | Depression | Conditions | |
| Recurrent major depression in remission | 433991 | Depression | Depression | Conditions | Excluded |
| Bipolar affective disorder, current episode depression | 439254 | Depression | Depression | Conditions | Excluded |
| Major depression, single episode | 4282096 | Depression | Depression | Conditions | Excluded |
| Bipolar affective disorder, currently depressed, in full remission | 439251 | Depression | Depression | Conditions | Excluded |

| | | | | | |
|---|---|---|---|---|---|
| Bipolar affective disorder, currently depressed, moderate | 437528 | Depression | Depression | Conditions | |
| Schizoaffective disorder, depressive type | 4224940 | Depression | Depression | Conditions | Excluded |
| Dysthymia | 433440 | Depression | Depression | Conditions | |
| Recurrent major depression in full remission | 4263748 | Depression | Depression | Conditions | Excluded |
| Single episode of major depression in full remission | 4025677 | Depression | Depression | Conditions | Excluded |
| Recurrent major depressive episodes, severe, with psychosis | 434911 | Depression | Depression | Conditions | Excluded |
| Chronic depression | 4103574 | Depression | Depression | Conditions | |
| Severe depression | 4149321 | Depression | Depression | Conditions | |
| Premenstrual dysphoric disorder | 4242733 | Depression | Depression | Conditions | Excluded |
| Recurrent depression | 4098302 | Depression | Depression | Conditions | |
| Moderate major depression, single episode | 4049623 | Depression | Depression | Conditions | Excluded |
| Mild major depression, single episode | 4195572 | Depression | Depression | Conditions | Excluded |
| Mild recurrent major depression | 4228802 | Depression | Depression | Conditions | |
| Recurrent major depression | 4282316 | Depression | Depression | Conditions | |
| Positive screening for depression on PHQ-9 (Patient Health Questionnaire 9) | 43021839 | Depression | Depression | Conditions | |
| Recurrent major depressive episodes, mild | 438998 | Depression | Depression | Conditions | Excluded |
| Severe major depression, single episode, with psychotic features | 438406 | Depression | Depression | Conditions | Excluded |
| Severe major depression, single episode, without psychotic features | 441534 | Depression | Depression | Conditions | Excluded |
| Moderate recurrent major depression | 4077577 | Depression | Depression | Conditions | |
| Bipolar affective disorder, currently depressed, mild | 439253 | Depression | Depression | Conditions | |
| Psychosis and severe depression co-occurrent and due to bipolar affective disorder | 35622934 | Depression | Depression | Conditions | |
| Severe recurrent major depression without psychotic features | 435220 | Depression | Depression | Conditions | |
| Mixed anxiety and depressive disorder | 4338031 | Depression | Depression | Conditions | |
| Reactive depression (situational) | 4314692 | Depression | Depression | Conditions | Excluded |
| Major depression single episode, in partial remission | 4323418 | Depression | Depression | Conditions | Excluded |
| Major depressive disorder | 4152280 | Depression | Depression | Conditions | |
| Severe recurrent major depression with psychotic features | 4154309 | Depression | Depression | Conditions | |
| Major depression in partial remission | 4148630 | Depression | Depression | Conditions | Excluded |
| Major depression in remission | 4176002 | Depression | Depression | Conditions | Excluded |
| Severe recurrent major depression | 43531624 | Depression | Depression | Conditions | |
| Single major depressive episode, severe, with psychosis | 439259 | Depression | Depression | Conditions | Excluded |
| Atypical depressive disorder | 438727 | Depression | Depression | Conditions | Excluded |
| Major depression with psychotic features | 37111697 | Depression | Depression | Conditions | |
| Mild major depression | 4336957 | Depression | Depression | Conditions | |
| Reactive depressive psychosis | 435520 | Depression | Depression | Conditions | Excluded |
| Major depression in full remission | 4269493 | Depression | Depression | Conditions | Excluded |
| Depressive disorder in remission | 44782943 | Depression | Depression | Conditions | Excluded |
| Moderate major depression | 4307111 | Depression | Depression | Conditions | |
| Menopausal depression | 4223090 | Depression | Depression | Conditions | |
| Severe major depression, single episode | 42872411 | Depression | Depression | Conditions | Excluded |
| Seasonal affective disorder | 4092239 | Depression | Depression | Conditions | Excluded |
| Recurrent major depressive disorder with postpartum onset | 4324959 | Depression | Depression | Conditions | |
| Severe major depression with psychotic features, mood-congruent | 4144233 | Depression | Depression | Conditions | |
| Multi-infarct dementia with depression | 443864 | Depression | Depression | Conditions | |
| Chronic depressive personality disorder | 40481798 | Depression | Depression | Conditions | |
| Moderately severe recurrent major depression | 36714998 | Depression | Depression | Conditions | |
| Depression screening positive | 762504 | Depression | Depression | Conditions | |
| Severe major depression with psychotic features | 4250023 | Depression | Depression | Conditions | |
| Endogenous depression | 4114950 | Depression | Depression | Conditions | |
| Mild depression | 4149320 | Depression | Depression | Conditions | |
| Acute depression | 37016718 | Depression | Depression | Conditions | |
| Severe postnatal depression | 4129184 | Depression | Depression | Conditions | |
| Moderately severe major depression | 36714389 | Depression | Depression | Conditions | |
| Severe major depression | 42872722 | Depression | Depression | Conditions | |
| Recurrent major depressive disorder with melancholic features | 4205471 | Depression | Depression | Conditions | |
| Major depression, melancholic type | 4154391 | Depression | Depression | Conditions | |
| Severe major depression without psychotic features | 4327337 | Depression | Depression | Conditions | |
| Moderate depression | 4151170 | Depression | Depression | Conditions | |
| Maternity blues | 4133073 | Depression | Depression | Conditions | |
| Drug-induced depressive state | 4103126 | Depression | Depression | Conditions | |
| Chronic recurrent major depressive disorder | 4094358 | Depression | Depression | Conditions | |
| Minimal major depression | 36715000 | Depression | Depression | Conditions | |
| O/E - depressed | 4038252 | Depression | Depression | Conditions | |
| Recurrent severe major depressive disorder co-occurrent with anxiety | 35615152 | Depression | Depression | Conditions | |
| Moderately severe depression | 36717092 | Depression | Depression | Conditions | |
| Recurrent moderate major depressive disorder co-occurrent with anxiety | 35615153 | Depression | Depression | Conditions | |
| Chronic major depressive disorder, single episode | 4031328 | Depression | Depression | Conditions | Excluded |
| Minimal depression | 36713698 | Depression | Depression | Conditions | |
| Depressive disorder in mother complicating pregnancy | 37018656 | Depression | Depression | Conditions | |
| Secondary dysthymia | 4224639 | Depression | Depression | Conditions | |
| Recurrent major depressive disorder in partial remission co-occurrent with anxiety | 35615155 | Depression | Depression | Conditions | Excluded |
| Postpartum depression | 4239471 | Postpartum depression | Depression | Conditions | |
